# Projected population level impact and cost-effectiveness of clinic and community-based tuberculosis screening approaches

**DOI:** 10.64898/2026.06.10.26355416

**Authors:** Nicky McCreesh, Palwasha Y Khan, Ian Yoon, Indira Govender, Mareca Sithole, Rein MGJ Houben, Richard G White, Alison D Grant, Sedona Sweeney

## Abstract

The South Africa National Department of Health have set ambitious targets to scale up TB testing, focusing primarily on clinic attendees. In the context of declining funding for TB care and prevention, the most cost-effective approaches for targeting testing should be identified.

We developed a mathematical model of TB in South Africa, explicitly incorporating clinic attendance by sex and HIV/ART status. We simulated six screening approaches over 2026–2035 (individually and in combination): three clinic-based (symptom screening, intensified targeted universal TB testing [TUTT, symptom-agnostic sputum testing of clinic attendees in key risk groups], and intensified TUTT allowing saliva samples) and three targeted community-based (community radiographic screening, symptom screening, and universal Xpert Ultra testing), each implemented at a range of coverage levels. Model outputs were combined with a mechanistic cost function to estimate potential impact and cost-effectiveness from a societal perspective.

The most cost-effective standalone approach was community radiographic screening at 10% annual population coverage, with an incremental cost-effectiveness ratio (ICER) of $421 per disability-adjusted life year (DALY) averted. 10/11 scenarios along the expansion path included community radiographic screening at progressively higher coverage, combined with a clinic-based approach.

Combining complementary approaches to reach both groups at increased risk of TB (e.g. clinic-based screening) and groups with lower screening coverage (e.g. community-based screening) may increase cost-effectiveness of TB screening, compared to standalone approaches. When designing TB screening strategies, both population risk and existing screening coverage should be considered.

## Introduction

Tuberculosis (TB) mortality in South Africa has fallen by only 17% over the past decade, well below the 75% reduction required to meet the End TB Strategy 2025 milestone(1). TB remains a leading cause of death in South Africa and an estimated 26% of people who develop TB are never diagnosed.

The South African National Department of Health’s End TB Campaign 2025/26(2) aims to test five million people for TB in 2025/26, primarily focussing on clinic-based testing, and with an aim to reduce transmission and improve treatment outcomes. The clinic strategy includes (1) Targeted Universal TB Testing (TUTT), which consists of testing clinic attendees in predefined risk groups (people living with HIV [PLHIV], people with recent TB, and recent close contacts) regardless of symptoms;(3) and (2) symptom screening all clinic attendees using the WHO four symptom screen.(4)

This drive to increase TB screening is occurring alongside large-scale cuts to the funding available for TB prevention and care worldwide. Following the abrupt dismantling of USAID and reduction of Global Fund contributions, estimates by the South African National Department of Health showed a net loss to TB funding of US$34 million, predicted to lead to 580,000 fewer people tested for TB in 2025.(5) Evidence is therefore needed to guide approaches to selecting populations for testing, and the choice of tests used, to maximise the health gains within a constrained budget.

While systematic screening for TB is recommended by WHO in high-burden settings, the certainty of evidence is low and there is no clear guidance on the choice of approach.(6) Existing evidence indicates that that active case finding (ACF) programmes tend to offer good value for money, but costs and cost-effectiveness are heterogeneous, context-specific, and vary considerably according to study methodology.(7–9)

In this paper, we use mathematical and economic modelling to identify the most cost-effective ACF approaches from a societal perspective, comparing a range of stand-alone and combined approaches to conducting screening in the community and among clinic attendees. We characterise the likely best investments depending on willingness and ability to pay for improved health, and describe potential routes for expansion as funding for TB increases.

## Methods

### Mathematical model

We developed an individual-based model of TB infection transmission, and TB mortality, diagnosis, and treatment, calibrated to national prevalence survey data and WHO estimates for South Africa.(10, 11) The model was stratified by sex and HIV/ART status. Infectious TB was divided into bacteriologically positive TB without and with any TB symptoms caused by TB (defined as one or more of cough (persistent of any duration), drenching night sweats, unexplained weight loss, and unexplained fever for at least two weeks; in line with the South Africa TB prevalence survey^10^). Self-cure occurred only in those without symptoms caused by TB; TB mortality occurred only in those with symptoms caused by TB. We explicitly modelled symptoms typical of TB which were caused by other conditions, and care seeking and symptom reporting could therefore occur from either TB state, as well as in people without TB. Clinic-visiting rates, seeking care for TB symptoms or for other reasons, varied by sex, HIV/ART status, and between individuals, informed using empirical data from South African.

The model was calibrated using history matching and model emulation (R package hmer(12)), an approach that aims to comprehensively capture uncertainty in both model parameters and calibration targets. Broad ranges were used for model parameters related to TB symptoms, reflecting the substantial uncertainty that exists in TB natural history. Variation infectiousness by HIV/ART and TB symptoms was explicitly simulated, and the model calibrated to estimates of the prevalence of IGRA positivity in child household contacts by index patient HIV/ART status and reported symptoms(13). Full model structure and calibration are described in the Supplementary material Section 1.1.

### Baseline and Intervention scenarios

We developed conceptual models through iterative consultation with country experts and the AHCOS study team to represent six screening scenarios (Supplementary material Section 1.1.13), with assumptions reflecting routine practice in South Africa. Intervention approaches were implemented from 2026 to 2035.

#### Baseline

In the baseline, PLHIV not on ART and HIV-negative individuals were tested only when seeking care for symptoms that would lead a clinician to consider TB. Clinic attendees on ART received limited ACF, consisting of a probability of annual symptom-agnostic testing with Xpert Ultra, and a 50-70% probability of symptom-screening with sputum collection if >6 months since prior ACF Xpert testing. Symptomatic people unable to produce sputum had a probability of further testing using other test modalities.

#### Community screening scenarios

For all community screening scenarios, we modelled a range of annual population coverages, assuming a higher prevalence in areas targeted when coverage was lower.

##### Community symptom screening

We assumed WBOTs conducted home-based symptom screening (one or more of persistent cough, night sweats, weight loss, fever), with sputum collection attempted for all symptomatic individuals. We assumed that 20% of symptomatic people unable to produce sputum underwent chest radiography at a district hospital.

##### Community radiographic screening

We simulated population screening with mobile vans fitted with chest radiography equipment interpreted by radiographers, with sputum collection attempted if the results showed any abnormality. A proportion of symptomatic individuals unable to provide sputum were referred for additional testing.

##### Community universal Xpert

We assumed Ward Based Outreach Teams (WBOTs) conducted home-based sputum collection, regardless of reported symptoms. No further testing would be conducted in people unable to produce sputum.

#### Clinic screening scenarios

##### Intensified targeted universal testing and treatment (TUTT)

We assumed all clinic attendees were screened for TUTT eligibility (selfZlreported HIVZlpositive status, recent TB treatment, recent close contact of someone with TB). Eligible individuals without an ACF Xpert within the past six months were asked for sputum, and those unable to produce sputum had a probability of further testing if symptomatic.

##### Intensified TUTT allowing saliva samples

As above, except that saliva samples were used if individuals were unable to produce sputum.

##### Universal clinic symptom screening

We assumed pragmatically that 75% of adults received symptom screening (one or more of persistent cough, night sweats, weight loss, fever), regardless of HIV/ART status, on arrival to the health facility. Symptomatic individuals without an ACF Xpert in the last 6 months were asked for sputum; those unable to produce sputum had a probability of further testing.

### Costing Approach

We estimated provider costs using primary data collected Feb 2024 at six public facilities in South Africa. We supplemented primary data collection with secondary cost estimates, with a prioritization on the study having taken place within the last 5 years and the study having taken place in South Africa. Costing methods are detailed further in the Supplementary material Section 1.2.

Costs were parameterized using a mechanistic cost function, which accounted for variations in unit costs at different scales of implementation. We disaggregated costs into fixed programme costs, fixed facility- and lab-level, and variable costs.(14) Fixed costs assumed no variation with scale and were set according to the national number of health facilities, labs, and WBOTs.(15) Variable costs represented inputs with constant returns to scale and were set according to the number of visits and tests in the model outputs.

Patient-incurred costs in the model were parameterised as costs of seeking care for TB symptoms and costs of TB treatment. We collected costs incurred by people prior to starting TB treatment in primary healthcare facilities in KwaZulu-Natal, in a cross-sectional survey of 99 people.(13) From 1 February 2022 to 29 February 2024, the study enrolled eligible index PWTB at the point of starting treatment at 13 primary healthcare (PHC) clinics within Umkhanyakude district, KwaZulu-Natal, South Africa. Enrolled participants >18 years gave written consent or witnessed verbal consent for those who could not read or write. We used a standardized questionnaire to estimate direct (including payment for accommodation, consultant fees, medicines, food, diagnostic fees for scans, tests and x-rays, procedures, travel and all other medical costs) and indirect costs (estimated using a human capital approach).

We assumed costs of seeking care were incurred by patients if they were diagnosed after seeking care for their TB symptoms, or if they died due to TB. We assumed those who were diagnosed through ACF before they sought care for their symptoms did not incur care-seeking costs. Costs incurred during TB and/or HIV treatment were sourced from the literature.(16) Patient-incurred costs were parameterized by HIV status, accounting for economies of scope where treatment was integrated.

All costs are presented in 2023 USD; methods for conversion are detailed in the Supplementary material. All cost estimates were applied using a gamma distribution across model runs. All costs were discounted at 5% in the base case.(17)

### Outcomes

We used disability-adjusted life years (DALYs) as our primary outcome measure; DALYs were estimated as the sum of years of life lost (YLLs) and years of life lived with disability (YLDs), and were parameterized based on time spent in different health states in the model (Ses Supplementary material Section 1.3 for details). All outcomes were discounted at 5% in the base case.(17)

### Cost-effectiveness and expansion paths

We estimated the proportion of parameter sets in which each of our six scenarios was cost-effective from a societal perspective. We used three potential cost-effectiveness thresholds reflecting the health spending elasticity of mortality and revealed willingness to pay for health in South Africa.(18, 19)

We then combined clinic-based and community-based scenarios to understand the potential expansion paths that South Africa might follow as available funding for TB increases. The total list of combined scenarios included in the expansion path analysis (including varying coverage for community-based from 10-80% of the population) is in Table S11. We mapped all approaches on the cost-effectiveness plane, identified the cost-effectiveness frontier and expansion path, and calculated incremental cost-effectiveness ratios (ICERs) between adjacent frontier points.

As this is an exploratory modelling exercise, no explicit prior health economics plan was developed. The study was discussed and approved by the AHRI community advisory board, and was approved by the district and provincial depts of health. Engagement with stakeholders did not change the approach or findings of the study. We complied with the Consolidated Health Economic Evaluation Reporting Standards (CHEERS 2022) reporting guidelines.(20)

### Sensitivity analysis

We conducted one-way sensitivity analyses to identify the impact of parameter and structural uncertainty on model outputs. We varied the discount rate (1-10%), time horizon (2-year and lifetime), Xpert Ultra pricing (reduced to mirror MiniDock MTB test pricing(21)), annual throughput of mobile radiography units (halved), costs and DALYs associated with false-positive diagnoses (removed) and the addition of CAD4TB interpretation software.

## Results

### Model fit to data

We generated 1041 parameter sets that were within the target ranges for all 42 calibration targets. Figure 1 shows the fit to data for 30 key calibration targets. The fit to the remaining 12 targets, and the values of the input parameters in the fitting parameter sets are shown in Tables S5 and S3 respectively.

**Fig 1:**
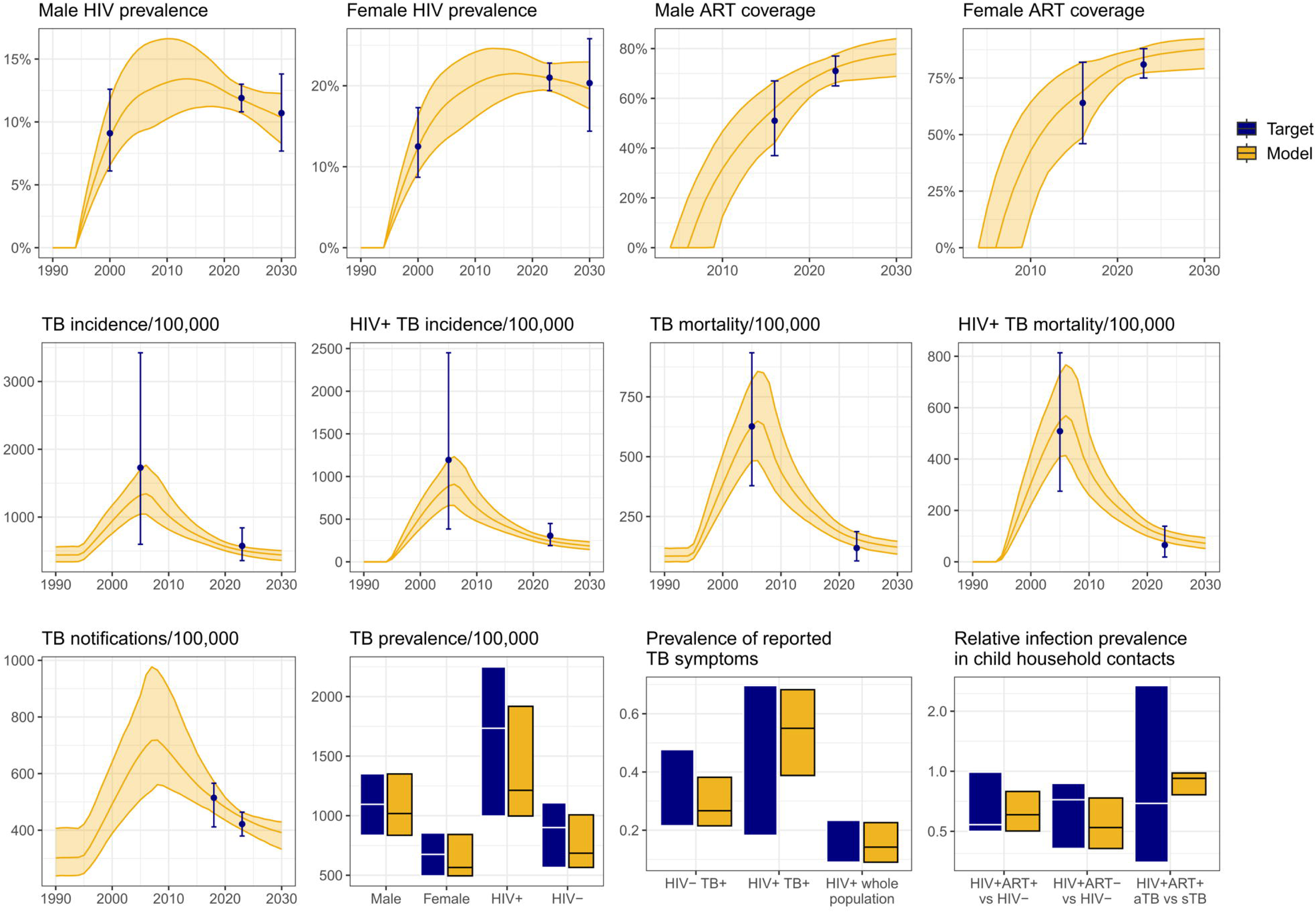
Model fit to data, for selected calibration targets.

### Intervention impact

Table 1 shows the mean reduction in TB incidence as compared with baseline in 2026 – 2035. Clinic symptom screening was the most effective at reducing incidence, with a mean reduction of 25% between 2026 – 2035 (95% UI 19 – 31%). This reflects the higher population-level coverage of Xpert testing compared with other approaches (median coverage of 9.7% in 2026, compared to 3.6% - 8.1%, Figure S5), and an increased probability of diagnosis per clinic visit for people with TB seeking care for TB symptoms, reducing diagnostic delays.

**Table 1:** Mean total and incremental costs and DALYs.

| Variable | Baseline | 1. Community symptom screening | 2. Community radiographic screening | 3. Community universal Xpert | 4. Intensified TUTT | 5. Intensified TUTT allowing saliva samples | 6. Clinic symptom screening |
| --- | --- | --- | --- | --- | --- | --- | --- |
| Mean annual incidence of infectious (bacteriologically-positive) TB in 2026-2035 | 438 | 423 | 391 | 389 | 376 | 385 | 328 |
| Mean reduction in the incidence of infectious TB in 2026-2035, relative to baseline |  | 3.4% | 10.7% | 11.2% | 14.2% | 12.1% | 25.1% |
| Total cost (2023 USD) | \$16.2B | \$16.9B | \$16.6B | \$17.3B | \$17.1B | \$17.2B | \$17.6B |
| Total provider- incurred cost | \$14.5B | \$15.1B | \$14.9B | \$15.6B | \$15.3B | \$15.5B | \$15.8B |
| Total patient- incurred cost | 1732.2M | 1735.3M | 1730.7M | 1736.4M | 1737.7M | 1740.8M | 1747.7M |
| Total DALYs | 137.6M | 137.3M | 136.7M | 136.7M | 136.3M | 136.3M | 135.3M |
| Years of life lost | 152.3M | 151.9M | 151.1M | 151.1M | 150.6M | 150.7M | 149.3M |
| Years of life lived with disability | 19.1M | 19.1M | 19.1M | 19.1M | 19.1M | 19.1M | 19.1M |
| Mean incremental cost (2023 USD) |  | 622.3M | 382.4M | 1090.8M | 841.3M | 965.8M | 1350.9M |
| Mean incremental DALYs averted |  | 0.3M | 0.9M | 0.9M | 1.3M | 1.3M | 2.4M |
| Mean ICER (2023 USD) | | \$1,900 | \$418 | \$1,184 | \$656 | \$779 | \$589 |

*M millions; B billions; TUTT targeted universal testing and treatment; DALYs disability-adjusted life years; ICER incremental cost-effectiveness ratio.*

The immediate effects of the approaches varied by HIV/ART status. Community approaches had similar Xpert coverage for all groups, but improved 2026 notifications most in HIVlZlnegative people due to longer disease duration and thus greater capture by screening (Supplementary figures S5 – S7). Clinic symptom screening resulted in the greatest increases in Xpert coverage and notifications among PLWHIV not on ART and HIV-negative people (Figures S5 and S6). It had little impact on screening coverage or notification rates in people on ART however, as we assumed that clinic symptom-screening coverage was already relatively high in the baseline scenario. Finally, the two intensified TUTT scenarios were the only scenarios that resulted in substantial increases in testing coverage (intensified TUTT and intensified TUTT allowing saliva samples) or notification rates (intensified TUTT only) among people on ART, as they allowed for more frequent testing among asymptomatic clinic attendees on ART.

### Costs and cost-effectiveness

The total and incremental costs and DALYs incurred over the 10-year time horizon are shown in Table 1 and Figure S10. Baseline was the least costly and least effective approach. Clinic symptom screening was the most costly and most effective. Incremental costs were largely driven by an increased quantity of Xpert Ultra tests, particularly in clinic-based scenarios. Incremental DALYs averted were driven by reductions in years of life lost (YLLs).

Figure 2 shows incremental cost and effectiveness estimates for all model runs, and Table S8 shows the percentage of model runs falling under each cost-effectiveness threshold. Community radiographic screening was the most cost-effective approach, with 98% of model runs falling under the lowest cost-effectiveness threshold ($618/DALY averted). At a threshold of $918/DALY averted, intensified TUTT (97%) and clinic symptom screening (99%) were also costlZleffective; at the highest threshold, all approaches were highly likely to be costlZleffective. Figure 3 shows the expansion path along the cost-effectiveness frontier, using all base and combined approaches. Mean cost, effect, and ICER estimates for all approaches are detailed in Table S9. Combined community and clinic intervention approach scenarios dominate stand-alone intervention scenarios. Ten of the eleven scenarios on the expansion path included community radiographic screening, advancing through progressively higher coverage levels, combined with a clinic-based approach (clinic symptom screen or intensified TUTT). Cost-effectiveness acceptability curves for approaches along the expansion path are shown in Figure S11. The likely best approach for adoption at a threshold of $619 ($985) per DALY averted was community radiographic screening covering 10% (20%) of the population, combined with intensified TUTT.

**Fig 2:**
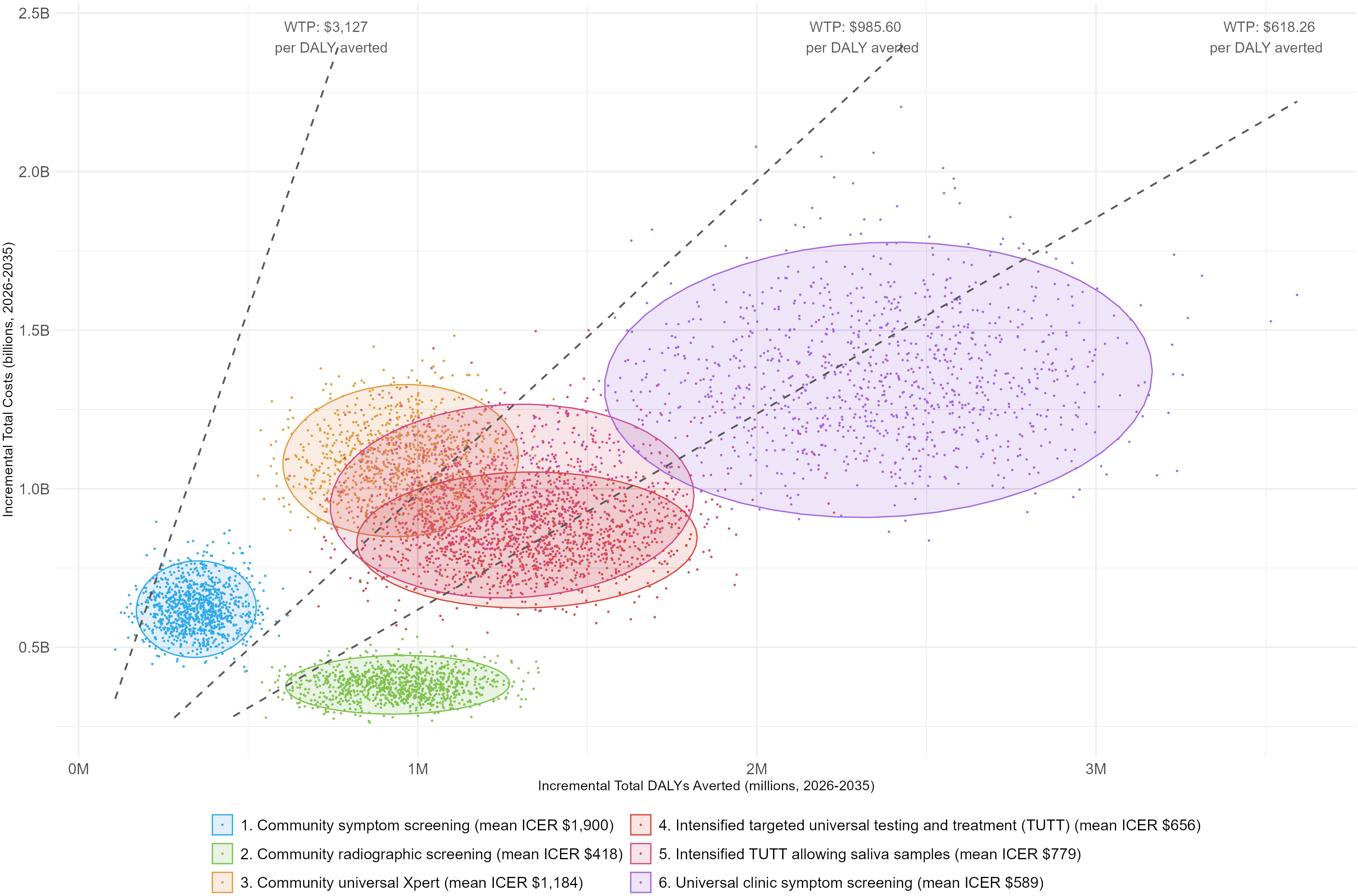
Cost-effectiveness plane

**Fig 3:**
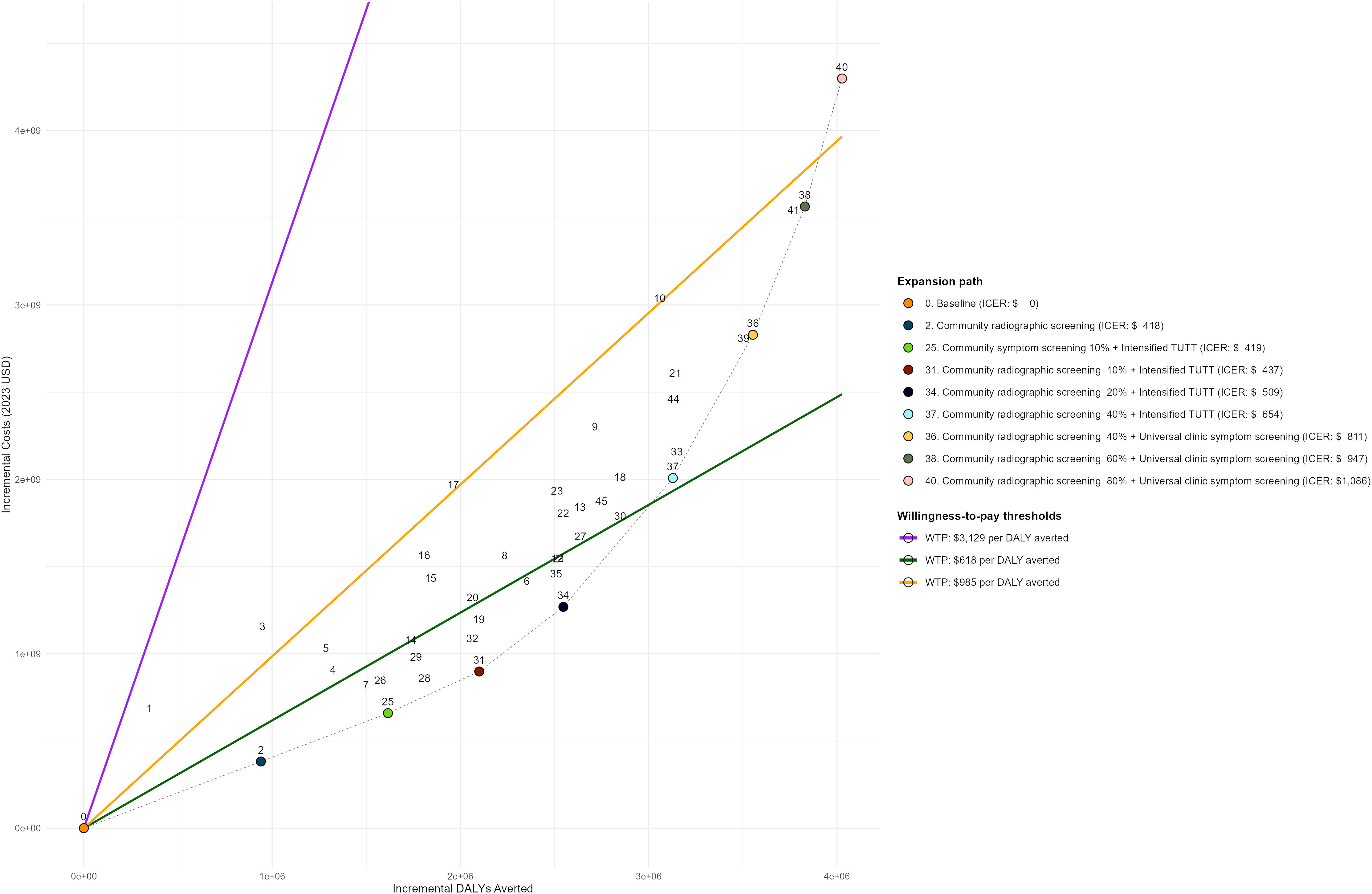
Expansion path

Each numbered point represents a stand-alone or combined screening scenario. Total estimates for all points are detailed in Table S9. ICER incremental cost-effectiveness ratio; DALYs disability-adjusted life years; USD United States Dollar; WTP willingness-to-pay.

Results from the one-way sensitivity analysis are shown in Figures S12 and S13. The choice of discount rate and time horizon had the largest impact on ICERs. A lifetime horizon and 1% discount increased most ICERs as compared with the base case, pushing community symptom screening and community universal Xpert above $3,127/DALY. Changing the price of Xpert Ultra to match that of MiniDock MTB, adding costs of CAD4TB software, and removing costs and impacts for false positive diagnoses, had minimal impact on all scenarios. Halving annual throughput for mobile chest radiography increased the mean ICER to $585, but did not change the order of approaches on the expansion path. Annual throughput as low as 2500 was likely to be cost-effective at a threshold of $985/DALY.

## Discussion

In our model-based cost-effectiveness analysis, community radiographic screening was the most cost-effective stand-alone approach to find people with TB in South Africa. Combining community and clinic-based approaches was consistently more cost-effective than stand-alone approaches. Our results suggest that a two-pronged approach, incorporating both clinic-level case finding and community-level chest radiography, is likely the best use of limited funds.

At the clinic level, we evaluated the relative cost-effectiveness of focusing on higher-risk groups (intensified TUTT) vs. symptom screening all clinic attendees regardless of risk. Our results highlight the importance of considering a group’s TB disease risk, their facility attendance, and existing screening coverage when designing a case finding programme. Although PLWHIV on ART are at elevated TB risk, they already attend facilities frequently for ART care, where some TB screening occurs. In contrast, HIV-negative individuals, particularly men, have far lower rates of clinic attendance,(22, 23) and do not receive the TB screening that can occur during visits for ART care. When considered in isolation, universal clinic symptom screening (which primarily increased testing coverage among people not on ART) was more cost-effective than intensified TUTT (which concentrated coverage gains among PLWHIV on ART); when combined with community screening, intensified TUTT was more cost-effective than universal clinic symptom screening. Because community screening independently increased coverage among people not on ART, the added value of clinic symptom screening diminished.

The ideal coverage of community radiographic screening depends on available funds and willingness-to-pay for improved health. There is currently no formal budget impact threshold to guide decision-making in South Africa.(24) We used three potential thresholds to give an indication of the potential cost-effectiveness (and affordability) of intervention approaches. We found that a population coverage of 10% is most likely to be cost-effective at our lowest threshold ($618/DALY averted), increasing to 20% at a threshold of $985/DALY averted. These may overstate willingness to pay for TB, particularly in the current global environment, however they provide a benchmark to assist policy makers in understanding the relative value of different approaches.

Diagnostic technology for TB is rapidly evolving, and new tests could offer wider reach and improved cost-effectiveness. In our univariable sensitivity analysis, we evaluated the potential impact of substantially reducing Xpert Ultra test costs, to match recently agreed costs for MiniDock MTB.(21) Reducing these costs reduced total costs in both baseline and intervention scenarios, resulting in limited effect on the incremental cost-effectiveness of our scenarios. In our univariable scenario adding fixed CAD4TB costs the expansion path was slightly different than in other scenarios. Our analyses suggest that the relative cost-effectiveness of screening approaches is driven primarily by overall design (who is screened, and how) rather than by the absolute cost of the diagnostic technology used. Community radiographic screening may also be more cost-effective if empirical treatment of people with high CAD scores and bacteriologically negative TB occurs.

A key strength of this work is that the individual-based modelling approach allowed us to simulate clinic visiting behaviour in detail, incorporating variation not only by HIV/ART status and sex, but also between individuals. This allowed us to represent in the model how frequently individuals could be reached in clinics and impose constraints on how frequently they could be tested through ACF, leading to more reliable estimates of the impact of the clinic-based approaches.

Another strength is our use of a mechanistic cost function allowing for distinction between fixed and variable costs. Costing analyses alongside transmission modelling often assumes constant marginal costs when services scale up, running counter to economic theory and ignoring important economies of scale and scope. Our approach allows for consideration of both economies of scale and scope and therefore provides a more robust estimate of national-level costs and cost-effectiveness.

A limitation of this work is that we were unable to account for health system constraints which limit the number of people tested for TB in model outputs. Several of our scenarios include high volumes of lab tests, which may not be operationally feasible. GeneXpert Ultra throughput may be lower in the real world due to staff shortages, equipment failures, and cartridge stockouts.(25) Our analysis supports existing evidence that a radiographic triage-based approach can reduce Xpert testing requirements by 31–60% with minimal reduction in yield.(26)

Community case finding is likely to be targeted at areas of the country where it is thought that there is a higher prevalence of TB. To model the relationship between intervention coverage and the prevalence among those screened, we used district-level variation in notification rates as a proxy—recognising that this is an imperfect measure. We may therefore be under- or over-estimating how the impact and cost-effectiveness of the community approaches scales with their coverage level. We also did not include the additional programme costs of covering larger population shares, which may require extra mobilisation, logistics, and governance resources, potentially raising ICERs at higher coverage. Given the geographic clustering of TB, simple geographic targeting would capture much of the burden, and subnational TB managers are often already aware where the highest prevalence lies.

Finally, there is substantial uncertainty in many aspects of TB natural history, including with respect to symptoms. TB symptoms can have alternative causes, even in people with TB, and asymptomatic and symptomatic TB may not represent distinct and stable states.(27) This is captured in the model by defining infectious TB states based on the presence or absence of TB symptoms caused by TB, and using wide ranges for input parameters related to TB symptoms, including values that allow for high rates of movement between the states. Additional discussion of assumptions made in the model structure and parameterisation is given in Supplementary material section 3.

Overall, our analysis demonstrates how combining behavioural, epidemiological, and economic data in a unified modelling framework can inform national screening strategies. We show that aligning programmes with both sub-population risk and real-world service use will be essential to maximise the health benefits of limited TB resources.

## Data Availability

The mathematical model code is publicly available on GitHub https://github.com/NickyMcC/AHCoS_model. Data from the AHCoS household contact study will be made available on the AHRI data repository at the time of publication https://data.ahri.org/index.php/home. Deidentified provider and patient-perspective cost data will be made available on the LSHTM Data Compass platform within 6 months of publication.

https://data.ahri.org/index.php/home

## Author contributions

Conceptualisation: NM, SS, ADG PYK; Funding acquisition: ADG, NM, RGW, RMGJH; Writing SS, NM; Methodology: SS, NM; Formal analysis: NM, SS, PYK; Data collection: SS, PYK, IG, IY. All authors reviewed the final draft.

## Acknowledgements

This study was funded by the National Institute of Allergy and Infectious Diseases (grant no R01A1147321).

The authors thank the study participants, community, members of the community advisory board, staff in the health facilities where the AHCoS household contact study was conducted, and the many people at AHRI and elsewhere who contributed to this work, including Mzamo Buthelezi, Nqubelo Dladla, Mlungisi Dube, Nomthandazo Mbatha, Nokuthula Jiyane, Nonkululeko Magagula, Ginger Mahlake, Sphephelo Masondo, Pinkie Mbotho, Sanele Mdletshe, Edwin Mkhwanazi, Mazani Mkhwanazi, Sanele Mngadi, Silindile Mthembu, Sizwe Ndlela, Nokuthula P. Ndlovu, Mbuso Ngema, Nomthunzi N. Ngema, Nothando Nkosi, Mpepho S. Ntombela, Simphiwe B. Ntshangase, Siyabonga Nxumalo, Thuthukani Nyawo, Nolwazi Nyuswa, Bongekile Nzuza, Bathandekile M. Phahlamohlaka, Zizile E. Sikhosana, Sibongokuhle M. Zulu and Professor Rodney Ehrlich, Dr Hobe, Sr Sindi Zikhali, Thabile Maphumulo, Nokuthula Nxumalo, Dr Hervey Vaughn-Williams, Zamani Dlamini, Siyabonga Gwala, Bonokwakhe Mkhwanazi, Sr Winile Mkhaliphi, Mr Philani Hadebe, Dr Salome Charalambous and Dr Limakatso Lebina

For the purposes of open access, the author has applied a Creative Commons Attribution (CC BY) licence to any Accepted Author Manuscript version arising from this submission.

## Data sharing

The mathematical model code is publicly available on GitHub https://github.com/NickyMcC/AHCoS_model. Data from the AHCoS household contact study will be made available on the AHRI data repository at the time of publication https://data.ahri.org/index.php/home. Deidentified provider and patient-perspective cost data will be made available on the LSHTM DataCompass platform within 6 months of publication.

## Supporting information captions

Supporting information 1. Additional methods, results, and discussion

Supporting information 2. Baseline and intervention approach flowcharts

