## Supporting info 1 for "Projected population level impact and cost-effectiveness of clinic and community-based tuberculosis screening approaches"

### Supporting information 1. Additional methods, results, and discussion

### 1 Supplementary Methods

#### 1.1 Description of the mathematical model

##### 1.1.1 Key

Model parameter names are written in italics, with colour indicating whether the parameter is an input parameter, a parameter with a global model-wide value, calculated from input parameter(s) or other values, or an individual-level parameter, which can take a different value for each simulated person.

In variable names, the terms HIV0, HIV1, and HIV2, are used to refer to people who are HIV-, HIV+ART-, and HIV+ART+ respectively.

*m/f* is used in parameter names to indicate that the parameter value varies by sex. *hiv0/1/2* and *hiv01/2* are used in parameter names to indicate that the parameter value varies by HIV and ART status and ART status only respectively.

##### 1.1.2 Use of expert opinion to inform model parameter ranges

It is a common misconception that adding additional complexity to mathematical models inevitably involves making additional assumptions, particularly if the data available to inform parameterisation are limited or of poor quality. This is usually not the case – instead, adding complexity makes implicit assumptions explicit, and allows the impact of the assumptions on the results to be explored. To give a simple example, if we choose to explicitly include sex in our model, we will make a range of explicit assumptions around how sex influences factors such as rates of developing disease, treatment rates, etc. If we do not explicitly include sex we still make assumptions, however we will be obliged to make the implicit assumptions that sex does not affect rates of developing disease, treatment rates, etc, and will not be able to explore the sensitivity of the results to the assumptions we made.

With that in mind, there are a numbers of areas in the model structure (particularly related to the impact on TB natural history of conditions other than TB that can cause TB symptoms) where we considered that it was important to include complexity, despite a lack of data to inform model parameterisation. For these areas, we relied on expert opinion to determine plausible ranges for the model parameters, favouring broad and inclusive ranges over more precise estimates. Ranges were obtained by consensus between three of the publication’s authors, Alison Grant, Palwasha Khan, and Indira Govender, all of who are clinicians and TB epidemiologists with extensive experience of primary healthcare in South Africa, along with advice from other South Africa experts.

##### 1.1.3 People

The main state variables assigned to people in the model were:

- Unique ID – *person_ID*
- Sex – *sex* (male, female)
- TB status – *TB_status* (uninfected, latent, subclinical disease, clinical disease, on treatment)
- Individual-level TB infectiousness – *infectiousness* (numeric, see Section 1.1.12 'Individual-level variation in infectiousness)
- HIV/ART status – *HIV_status* (HIV-, HIV+ART-, HIV+ART+)
- Whether they have a chronic health condition (other than or in addition to TB) that causes TB symptoms – *chronic_status*
- Their baseline clinic visiting rate – *my_baseline_clinic_rate* – and an adjusted rate that accounts for any higher clinic visiting rate in people with symptoms, and for higher clinic visiting rates for people on ART – *my_adjusted_clinic_rate*

Other state variables were used to track individuals' histories in the model, for the purpose of creating model output.

##### 1.1.4 Model initialisation

To initialise the model, 20000 people were created. The newly created people were each assigned a *sex*, with a probability of *prop_male* of being male and 1 - *prop_male* of being female. They were then assigned a time to (non-HIV and non-TB) death, set equal to:

minimum (*max_age* – *min_age*, (*min_background_mort_age_m/f* – *min_age* – ln (1 - random-float 1) / *background_mort_m/f*)

A random *infection_seed_proportion* were seeded with latent infection, with no risk of progression without reinfection. A random *tb_seed_proportion* were seeded with subclinical TB disease.

The model was run from 1850 to allow the population age distribution and TB incidence and mortality to reach equilibrium, before the introduction of HIV in *hiv_intro_year*.

##### 1.1.5 Model scheduling

The majority of events in the model were simulated using continuous time.

The exceptions to this were the *Mtb* transmission process, which used a monthly time step, and the production of model output, which was done annually.

##### 1.1.6 Demography

Individuals were introduced into the model at age 15 (*min_age*). People aged <15 were not modelled, as the risk of *Mtb* transmission from children is low[1], and the simulated intervention approaches considered screening in adults only.

A constant population size was simulated, with a new person created whenever someone died in the model.

There were four types of mortality in the model

- HIV mortality
- TB mortality
- Background mortality
- All individuals die upon reaching the age of 80 years (*max_age*)

The background mortality started at age *min_background_mort_age_m/f*, and had a rate of *background_mort_m/f* per year. The values of *min_background_mort_age_m/f* and *background_mort_m/f* were fixed, and chosen to give an approximate fit to the age distribution by sex in South Africa in 2021[2].

TB and HIV mortality are described in the sections on TB and HIV.

##### 1.1.7 TB symptoms

We define TB symptoms in line with the 2018 South Africa TB prevalence survey: reporting one or more of cough (persistent of any duration), drenching night sweats, unexplained weight loss, and unexplained fever for at least two weeks[3]. Approximately 15% HIV-negative people and 23% of HIV-positive people without TB were symptom-screen positive in the 2018 South African TB prevalence survey[4], demonstrating that TB symptoms are relatively common even in people without TB.

Mathematical models of tuberculosis that explicitly model TB symptoms have typically divided the bacteriologically-positive TB state into two compartments, symptom-screen negative and symptom-screen positive (often referring to the compartments as subclinical or asymptomatic and clinical or symptomatic respectively). With this model structure, people enter the symptom-screen negative compartment when first developing bacteriologically-positive TB. This results in a model structure where the probability of reporting symptoms falls from frequently >10% in people without bacteriologically positive TB to zero in people after first they first develop bacteriologically positive TB. We therefore explicitly take into account in our model structure the presence of people with bacteriologically-positive TB and TB symptoms whose symptoms are not caused by TB.

Recent WHO guidance[5] defines asymptomatic TB as "*A person with TB disease who did not report symptoms suggestive of TB during screening*" – i.e. a pragmatic division into asymptomatic and symptomatic TB based on whether someone reports TB symptoms, regardless of the underlying cause of the symptoms. The slightly earlier International Consensus for Early TB framework (ICE-TB)[6] took a more conceptual approach to classifying early TB states, saying that "*we agreed that each conceptual state should reflect pathophysiological processes, rather than be bound solely by practical considerations, such as the ability to identify states with existing diagnostic tools*", and describing symptoms as "*caused by the host response to M. tuberculosis".* ICE-TB also favoured the term subclinical over asymptomatic to refer to people with TB who do not recognise and report symptoms. For the purposes of this manuscript we retain this distinction, using the terms **asymptomatic** and **symptomatic TB** to refer to people with TB who do not or do report symptoms during screening (in line with WHO guidance), and the terms **subclinical** and **clinical TB** to refer to people without and with reported symptoms that are *caused by TB* respectively. People with subclinical TB can therefore be symptomatic or asymptomatic, based on the presence of absence of TB symptoms caused by conditions other than TB.

There are a wide range of conditions that can cause TB symptoms that are not TB, ranging from minor transient conditions such as the common cold, to chronic conditions such as chronic obstructive pulmonary disease. Transient conditions are likely to have little effect on TB model dynamics, beyond temporarily making some people symptom-screen positive. Chronic conditions may have a greater effect on model dynamics – for instance potentially increasing TB mortality rates in people with some chronic conditions, but also potentially making them detectable through passive case finding from as soon as they develop bacteriologically-positive TB.

In the model, we simplify conditions that cause TB symptoms that are not TB into two categories: chronic and transient. We assume that the presence or absence of chronic conditions is a permanent characteristic of simulated individuals that can increase the rate of developing disease and alter disease progression. We assume that people with chronic conditions always report symptoms when asked. Transient conditions are not modelled as a characteristic of people. Instead, all people in the model have a small, non-zero probability of reporting TB symptoms due to transient conditions each time they are symptom screened.

###### *1.1.7.1 Development and loss of TB symptoms caused by TB*

Few data are available to inform the rate at which people with subclinical TB develop clinical TB, or the rate at which people with clinical TB regress to subclinical TB. A recent review estimated that, across national prevalence surveys and adjusted for under-diagnosis, 17.2% of people with prevalent bacteriological TB reported a persistent cough (duration ≥2 weeks), and 37.5% reported a cough of any duration (with figures of 10.3% and 16.8% respectively for South Africa)[7]. From this, we can estimate that an average of 54% of people with bacteriological TB who reported a cough reported that they had had their cough for less than 2 weeks (39% for South Africa). In a community prevalence survey conducted in KwaZulu-Natal, South Africa, there was a mean reported symptom duration in people with bacteriologically positive TB of 2.8 weeks, and a median duration of 2 weeks[8].

These estimates suggest that there may be a high rate of movement between the symptomatic and asymptomatic states. However reported symptom duration may be affected by inaccurate recall, and the national estimates are based on cough only (as opposed to the four-symptom screen). The data are also for reported symptoms regardless of cause, not symptoms caused by TB, and rates of loss of symptoms may by higher for symptoms caused by other things. For these reasons, we used a wide range for the rates of movement from clinical to subclinical TB in HIV negative people (i.e. loss of TB symptoms caused by TB), ranging from no regression to regression at a mean rate of 1 per month. We assumed that the rate of regression must be lower in people living with HIV than in HIV-negative people, and lower in PLWHIV who are not on ART than in PLWHIV who are on ART.

Calibrating the model to data from the South Africa National prevalence survey and to WHO data and estimates, most notably estimates of the proportion of people with prevalent TB who are symptom-screen negative, restricts the values of the rates of movement from subclinical to clinical TB relative to the rates of movement from clinical to subclinical TB. For this reason, we used wide uninformative ranges around the parameter controlling the rate of movement from subclinical to clinical TB. We assumed that the rate of progression must be higher in people living with HIV than in HIV negative people, and higher in PLWHIV who are not on ART than in PLWHIV who are on ART.

We assumed that having a chronic condition that causes TB symptoms would decrease the rate of regression from clinical to subclinical TB, and increase the rate of progression from subclinical to clinical TB (see Section 1.1.7.2 for details).

###### *1.1.7.2 Chronic conditions that cause TB symptoms*

We assumed that chronic conditions that cause TB symptoms can affect TB natural history and care seeking in a number of ways:

- Alter the rate of developing TB following infection
- Alter the rate at which people move between subclinical TB and clinical TB
- Alter the rate of self-cure for people with subclinical TB

- Alter the TB mortality rate for people with clinical TB
- Lead to passive treatment seeking and being tested for TB in people with subclinical TB
- Alter the level of infectiousness per unit time

Few empirical data are available to inform these rates, due to the complex array of conditions that can cause TB symptoms, and the difficulties in distinguishing whether symptoms are caused by TB or other conditions in people with TB. We therefore relied on expert opinion (see Section 1.1.2) to obtain broad plausible ranges for parameters related to the above factors, as well as for the proportion of prevalent TB symptoms in people without bacteriologically positive TB that are due to chronic conditions.

**Proportion of prevalent TB symptoms in people without bacteriologically positive TB in South Africa that are due to chronic conditions**

It was considered plausible that 30% - 70% of prevalent TB symptoms in HIV- people without bacteriologically positive TB are due to chronic conditions (*proportion_symp_chronic_hiv0*), with the remaining 70% - 30% due to transient conditions. It was considered that the proportion due to chronic conditions in HIV+ people was likely to be higher by an absolute value of 10% (*abs_increased_prop_symp_chronic_hiv*):

*proportion_symp_chronic_hiv12* = *proportion_symp_chronic_hiv0* + *abs_increased_prop_symp_chronic_hiv*

**Rate of developing TB following infection, and TB progression**

It was considered plausible that having a chronic condition that causes TB symptoms could increase the rate of developing TB following infection (*increased_TB_development_chronic*), and the rate of progression from subclinical to clinical TB, by a ratio of 1.3 to 1.9, relative to people with no chronic conditions (*relative_rate_sub_to_clin_chronic*).

It was considered plausible that having a chronic condition that causes TB symptoms could decrease the rate of self-cure from subclinical TB (*relative_self_cure_rate_chronic*), and the rate of regression from clinical to subclinical TB (*relative_rate_clin_to_sub_chronic*), by a ratio of 1/1.3 to 1/1.9, relative to people with no chronic conditions.

Overall, it was considered plausible that having a chronic condition that causes TB symptoms could increase the overall rate of TB mortality by a ratio of 1.7 to 3, relative to people with no chronic conditions. As we modelled TB mortality occurring from clinical TB only, and the proportion of prevalent bacteriological TB that was clinical was higher for people with chronic conditions in the model, we modelled this through assuming that the rate of TB mortality in the clinical state was 1.3 – 1.9 times higher for people with chronic conditions than for people without (*relative_TB_mortality_rate_chronic*). After the model was calibrated, we checked in the fitting parameter sets that this gave an overall rate of TB mortality for people with chronic conditions compared to those without of approximately 1.7 to 3 (see Section 2.1.2).

**Passive diagnosis**

It was considered plausible that the rate of seeking care for TB symptoms caused by chronic conditions only could be between 0.5 and 2 times that of the rate of seeking care for TB symptoms caused by TB (*clinic_TBsymp_adjust_chronic*).

**Alter the level of infectiousness per unit time**

We assume that the probability of transmission per contact for people with subclinical TB and a chronic condition is dependent of whether they have a cough. If they do not, we assume that the probability of transmission per contact is the same as for people with subclinical TB and no chronic condition. If they do have a cough, we consider it plausible that the probability of transmission per contact could be between 10% and 50% higher than for people with subclinical TB and no chronic condition. As the type of TB symptom (cough vs not) was not explicitly simulated, this was simulated as a single transmission multiplier (relative to clinical TB) applied to all people with subclinical TB and a chronic condition:

*transmission_multiplier_chronic * 0.587 + reduced_transmission_sub * (1 – 0.587)*

Where 0.587 is the proportion of all people who reported TB symptoms in the South Africa TB prevalence survey who reported a cough[9].

##### 1.1.8 TB infectiousness

The parameter in our model *reduced_transmission_sub* controls the relative infectiousness per unit time of people with subclinical TB and no chronic condition, relative to people with clinical TB, encompassing both differences in contact behaviour and in the probability of transmission per contact. From a physiological perspective, we consider it unlikely subclinical TB is more infectious per unit time than clinical TB. Having TB symptoms may reduce the amount of contact that someone has with other people however. We therefore set a range for *reduced_transmission_sub* of 0.2 to 1.3. Infectiousness in people with subclinical TB and chronic conditions is described in Section 1.1.7.2

##### 1.1.9 Clinic visiting and diagnosis

###### *1.1.9.1 Overview*

We model two separate types of clinic visit: visits made seeking care for TB symptoms, and visits made for any other reason (not seeking care for TB symptoms). These types of clinic visit vary in the rates at which they occur, and in the probability of TB screening and testing occurring during the clinic visit. Clinic visiting by people with TB is explicitly modelled throughout the whole model run. Clinic visiting by people without TB is modelled from 1990 only, as it greatly increases the model runtime, and has negligible impact on TB dynamics in the baseline model.

Empirical data from clinic visit surveys conducted in South Africa suggest that even in people reporting TB symptoms, seeking care for those symptoms is the reason for the clinic visit in only 9%[10] or 35%[11] of visits. Therefore, we model a rate of clinic visiting not seeking care for TB symptoms in all simulated individuals, including those who do have TB symptoms (see Section 1.1.9.2). On these visits, we assume that there is no probability of diagnosis for people not on ART (in the absence of clinic-based interventions), and that diagnosis can occur through an active case finding pathway for people on ART.

In addition, simulated individuals with TB symptoms caused by chronic conditions and/or TB have a rate of seeking care for those symptoms in the model, and we simulate a rate of seeking care for TB symptoms caused by transient conditions for all individuals without TB symptoms caused by TB or chronic conditions. On each visit seeking care for TB symptoms, these individuals have a probability of diagnosis through active case finding (if on ART or in the clinic-based intervention scenarios), and also have an additional probability of diagnosis through passive case finding.

###### *1.1.9.2 Rate of clinic visits made for any other reason (not seeking care for TB symptoms)*

###### 1.1.9.2.1 Individual-level baseline clinic visiting rates

###### 1.1.9.2.1.1 Data

Data used to determine how rates of clinic visiting vary by sex and ART status, and between individuals, were taken from the Africa Health Research Institute (AHRI) demographic surveillance area (DSA) in KwaZulu-Natal, South Africa, from 1st February 2017 to 29th February 2020. The data sources are described in Randera-Rees *et al* 2021[12]. The data used were limited to people who were aged 15 years and over. It was not possible to distinguish in the data between people who did not visit a clinic at all, and people who visited a clinic but did not consent to their data being recorded. To reduce underestimation of clinic visiting rates, we therefore limited the analysis to people who had consented to have at least one HIV test conducted as part of a DSA serosurvey – people who we considered were also likely to consent to their clinic visits being recorded.

ART interruptions were not explicitly simulated in the model, and periods of treatment interruption contributed only 8.3% of all person time for people who were or had previously been on ART in the dataset. In the analysis, we therefore considered individuals to be on ART from the first date at which a clinic visit for ART-related reasons was recorded, including during interruptions.

HIV status was unknown for 53% of person-time for people not on ART, and there was little difference in the mean rate of clinic visiting between people who were HIV positive and not on ART and of unknown HIV status (0.70 and 0.76 visits per year respectively). Visits for people who were known to be HIV negative were higher (1.4 per year), however it is likely that this is an overestimate – only people who had a recent negative HIV test at a clinic or in a DSA sero-survey were considered to be HIV negative. Limiting the analysis to people who were recorded as visiting a clinic at least once during the data collection period greatly reduced the difference between HIV negative people and people with unknown HIV status (to 2.7 and 2.2 visits per year respectively). Patterns of clinic visiting were therefore assumed to be the same in all people who were not considered to be on ART.

For each sex and ART category, a gamma distribution was fitted to the data on the annual rate of clinic visiting per person (Figure S1).

Figure S1. Distribution of the annual rate of clinic visiting in the data and fitted gamma distribution, by sex and ART. The x-axis shows the cumulative proportion of who visit clinics at at least the given rate.

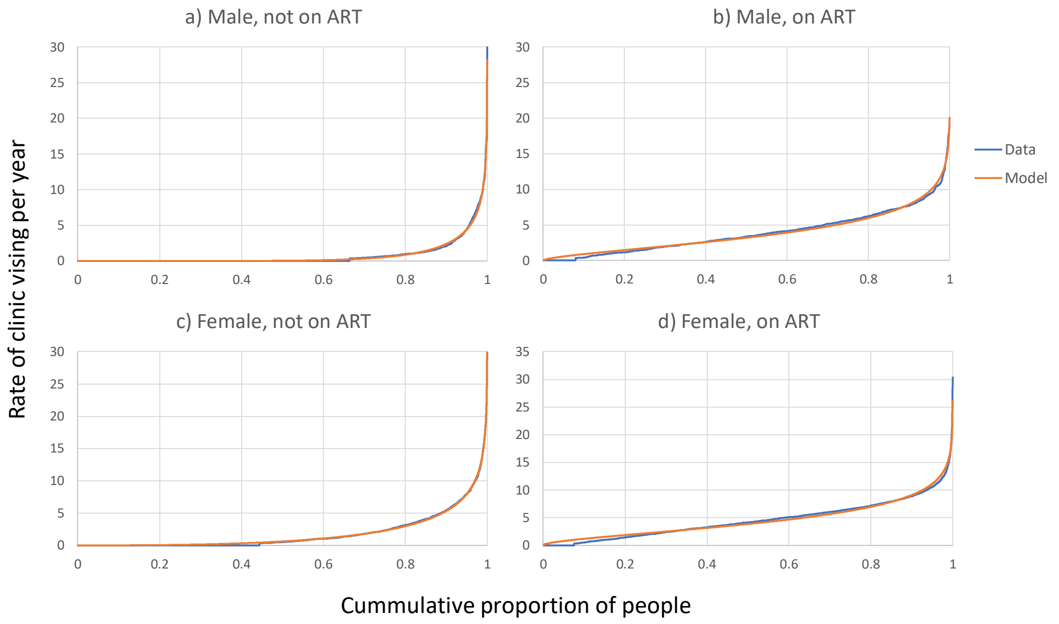

###### 1.1.9.2.1.2 Model

In the model, an initial baseline rate at which a person visited clinics for reasons other than seeking care for TB symptoms was sampled at their creation at random from a gamma distribution, with the parameter values varying by sex (Figure S1). The baseline rate is then multiplied by *clinic_visit_adjust*, a parameter that allows the models to be calibrated to National data on clinic visit rates (see Section 1.1.9.5). This gives an individual their final baseline rate of visiting clinics, *my_baseline_clinic_rate*. Upon starting ART, their *my_baseline_clinic_rate* and time of next clinic visit were resampled.

###### 1.1.9.2.2 Adjustments to clinic visit rates in people with TB symptoms and/or TB

Empirical data suggest that people who report TB symptoms may have higher rates of visiting clinics, and that this may not be entirely explained by them seeking care for those symptoms. For instance, in one clinic exit survey in South Africa, 50% of respondents reported TB symptoms, with those symptoms the main reason for attending the clinic for only 35% of them[11]. It is plausible that the relative increase in visiting rates may be lower for people on ART compared to people who are HIV- or HIV+ART-, due to their high rates of clinic visiting for routine HIV care. We assume that having TB symptoms caused by chronic conditions and/or TB increases the rate of clinic visiting by a factor of:

- *clinic_other_symp_hiv01* in people on ART
- *clinic_other_symp_hiv2* = 1 + (*clinic_other_symp_hiv01* – 1) * *clinic_other _hiv2_vs_hiv01* in people who are not on ART

We further assume that having bacteriologically positive TB (regardless of symptoms) may be associated with an increased rate of clinic visiting, due to the existence of comorbidities (e.g. diabetes) that both increase clinic visiting rates, and increase the risk of developing TB. This is modelled by increasing the clinic visiting rate in people with TB by a factor of:

- *clinic_other_tb_hiv01* in people on ART
- *clinic_other_tb_hiv2* = 1 + (*clinic_other_tb_hiv01* – 1) * *clinic_other _hiv2_vs_hiv01* in people who are not on ART

Transient conditions causing TB symptoms are simulated as a probability of reporting symptoms when asked in people without TB symptoms caused by TB or chronic conditions, and not as an individual-level characteristic. They are therefore simulated in clinic attendees as an increased probability of reporting symptoms in clinic attendees compared to the general population, rather than as an increased rate of attending clinics in people with TB symptoms. The probability of a clinic attendee reporting symptoms due to transient conditions is calculated as:

prob_transient_clinic_hiv0/1/2 = (prob_transient_comm_hiv0/1/2 * clinic_other_symp_hiv01/2 / (prob_transient_comm_hiv0/1/2 * clinic_other_symp_hiv01/2 + (1 - prob_transient_comm_hiv0/1/2)))

where:

- *prob_transient_comm_hiv0* = (*prop_with_symp_hiv0* - *prop_with_symp_hiv0* * *proportion_symp_chronic_hiv0*)/(1 - *prop_with_symp_hiv0* * *proportion_symp_chronic*)
- *prob_transient_comm_hiv1/2* = (*prop_with_symp_hiv1/2* - *prop_with_symp_hiv1/2* * *proportion_symp_chronic_hiv12*)/(1 - *prop_with_symp_hiv1/2* * *proportion_symp_chronic_hiv12*)

and:

- *prop_with_symp_hiv1* = *prop_with_symp_hiv0* + *prop_with_symp_input_hiv1* * (1 - *prop_with_symp_hiv0*)
- *prop_with_symp_hiv2* = *prop_with_symp_hiv0* + *prop_with_symp_input_hiv1* * *prop_with_symp_input_hiv2* * (1 - *prop_with_symp_hiv0*)

###### 1.1.9.3 Rate of clinic visiting seeking care for TB symptoms

We model a rate of seeking care for TB symptoms caused by TB that varies by HIV and ART status, equal to *clinic_TBsymp_HIV0/1/2*. The rate of seeking care for TB symptoms could be higher or lower in people whose symptoms are caused by chronic conditions only, and the rates are therefore multiplied by *clinic_TBsymp_adjust_chronic* in those individuals.

For people without TB symptoms caused by TB or chronic conditions, we model a rate of seeking treatment for TB symptoms caused by transient conditions equal to:

*prob_transient_comm_hiv0/1/2 * clinic_TBsymp_HIV0/1/2 * clinic_TBsymp_adjust_chronic * clinic_TBsymp_adjust_transient*

allowing for a lower rate of seeking treatment for TB symptoms caused by transient conditions compared to TB symptoms caused by TB and/or chronic conditions.

###### 1.1.9.4 Screening and diagnosis

Supplementary material 2 shows the simulated active screening pathway simulated in the model in the baseline scenario. In brief, we assume that people on ART have a probability of receiving an Xpert Ultra if it has been at least a year since they last received an Xpert Ultra through active case finding, or at least 6 months if they report TB symptoms. Individuals reporting TB symptoms who are not able to produce sputum are referred to a doctor, with an approximately 24.6% chance that they will be diagnosed with TB. The same screening process is followed when people first start ART. In line with expert opinion (see Section 1.1.2), we assume that there is a low probability of active screening for people not on ART, and therefore do not simulate active screening in that group.

We assume that there is an additional probability of diagnosis and starting treatment on each clinic visit made by people seeking care for TB symptoms, equal to *passive_diagnosis_prob*.

###### *1.1.9.5 Calibration targets*

National level data from South Africa reported a mean of 1.7 primary healthcare visits per year in 2022 for all ages[13], and there are some data to indicate that the mean rate of clinic visiting may not vary greatly between 0-14 year olds and 15+ year olds[14]. We therefore calibrated the model so that the overall mean rate of clinic visiting per year in 2022 was 1.5 – 1.9.

We calibrated the model to a 0.66 – 4.63 times higher prevalence of TB in clinic attendees relative to in people in the general population in 2022, informed by data from clinic-based and community-based TB surveys conducted in KwaZulu-Natal, South Africa[8,15]. The upper bound was increased from the crude estimate of 2.87 to account for the much lower proportion of people able to provide a sputum sample in the clinic-based survey compared to in the community-based survey. The lower bound was decreased from the crude estimate of 1.04 to account for the fact that only people who were screen positive were eligible to provide a sputum sample in the community survey.

Data on the prevalence of TB symptoms in clinic attendees in South Africa are limited, and estimates are highly variable. In one clinic exit survey, 64% of people reported symptoms, however only 19% of eligible people participated, and it is plausible that participation was higher among people with symptoms[10]. In a second clinic exit survey, 50% of people reported symptoms, however the authors reported that "*Among those screened for eligibility to the study, the proportion of participants reporting TB symptoms (50%) seems very high and we could not verify if study staff at times pre-screened participants*"[11]. In a third survey, only 6.4% of people reported symptoms[15]. In line with expert opinion (see Section 1.1.2), we calibrated the model so that a maximum of 30% of simulated clinic attendees reported TB symptoms. We did not set a lower bound for the prevalence of TB symptoms in clinic attendees, as the model structure ensured that it could not be lower than the prevalence in the general population (by HIV/ART status).

##### 1.1.10 HIV/ART

Three HIV states were simulated in the model: HIV-, HIV+ART-, and HIV+ART+.

People created in the model at age 15 years were all HIV-. From the introduction of HIV in the model in 1995, HIV- people became HIV+ART- at a rate that varied by sex and over time. HIV incidence had the value *hiv_inc_early_m/f_annual* from 1995 to 2000, decreased/increased linearly from *hiv_inc_early_m/f_annual* to *hiv_inc_mid_m/f_annual* between 2000 and 2020, and decreased/increased linearly from *hiv_inc_mid_m/f_annual* to *hiv_inc_mid_m/f_annual* * *hiv_inc_m/f_late_reduction* between 2023 and 2040.

ART was introduced in the model in 2010. From the introduction of ART, HIV+ART- people became HIV+ART+ at a rate that varied by sex. To capture changes in estimated ART coverage over time, the values of the ART start rates in the model were changed in 2017, from *ART_start_rate_early_m/f_annual* to *ART_start_rate_late_m/f_annual*. People starting ART were actively screened for TB, as described in Section 1.1.9.4.

From 2010, all HIV+ART- people starting TB treatment were made HIV+ART+.

HIV mortality was simulated as a constant rate of (non-TB) HIV-related mortality for all HIV+ART- people (*HIV1_mortality_rate*), and all HIV+ART+ people (*HIV2_mortality_rate*). This was estimated from UNAIDS estimates[16] of HIV-related mortality in adults in 2021 (excluding estimated numbers of deaths due to TB), divided by the estimated number of adults living with HIV, and assuming that mortality rates were 52% lower for HIV+ people on ART than HIV+ people not on ART[17].

###### *1.1.10.1 HIV and TB*

We assume that PLWHIV who are on ART have 1.47 – 10.8 times the rate of developing TB following infection compared to HIV negative people[18,19] (*increased_develop_tb_rate_HIV2_vs_HIV0*). We assume that PLWHIV who are not on ART can have 1.89 – 4.76 times the rate of developing TB following infection compared to PLWHIV who are on ART (*increased_develop_tb_rate_HIV1_vs_HIV2*), based on the findings of a systematic review, with widened bounds to reflect variation in CD4 counts[20].

There are few or no data available to inform how HIV and ART affect the rate of transitioning between subclinical and clinical TB, or mortality rates from untreated clinical TB. We therefore assumed that rates of developing TB symptoms caused by TB and TB mortality rates from untreated clinical TB must be higher for people who are HIV+ART- than for people who are HIV+ART+, and higher for people who are HIV+ART+ than for people who are HIV-. Similarly, we assumed that rates of losing TB symptoms caused by TB must be lower for people who are HIV+ART- than for people who are HIV+ART+, and lower for people who are HIV+ART+ than for people who are HIV-.

###### *1.1.10.2 Calibration targets*

The model was calibrated to the estimated male and female adult HIV prevalence in 2000 and 2023, and to the estimated male and female adult ART coverage in 2016 and 2023[21]. The width of the confidence intervals around the estimated prevalence of HIV and ART coverage in South Africa were reduced greatly between the UNAIDS estimates published in 2023 and 2024. It is likely that the reduction in confidence interval widths for 2000 and 2016 reflect reduced uncertainty for more recent years following the 2022 South Africa HIV prevalence survey[22], and assumptions made in generating UNAIDS prevalence estimates, rather than a genuine reduction in uncertainty around the estimates for earlier years. For that reason, we calibrated the model to the wider 2023-generated uncertainty intervals for HIV prevalence in 2000 and ART coverage in 2016.

The model was also calibrated to the predicted male and female adult HIV prevalence in 2030 from the Thembisa model version 4.7[23], with widened uncertainty ranges. Finally, the model was calibrated to ratios of ART in 2023 vs 2019 by sex greater than one, which ensured that any otherwise fitting runs where ART coverage goes down in the future were excluded.

See Section 1.1.11.9 for details of the calibration targets for HIV-related TB.

##### 1.1.11 Tuberculosis

###### *1.1.11.1 Disease states*

Each individual in the model was in one of five main TB states (uninfected, latent, subclinical TB (no TB symptoms caused by TB), clinical TB (TB symptoms caused by TB), on treatment), with the latent infection state subdivided by time since infection (Figure S2).

Figure S2 Simulated TB states. Red outlines indicate infectious states.

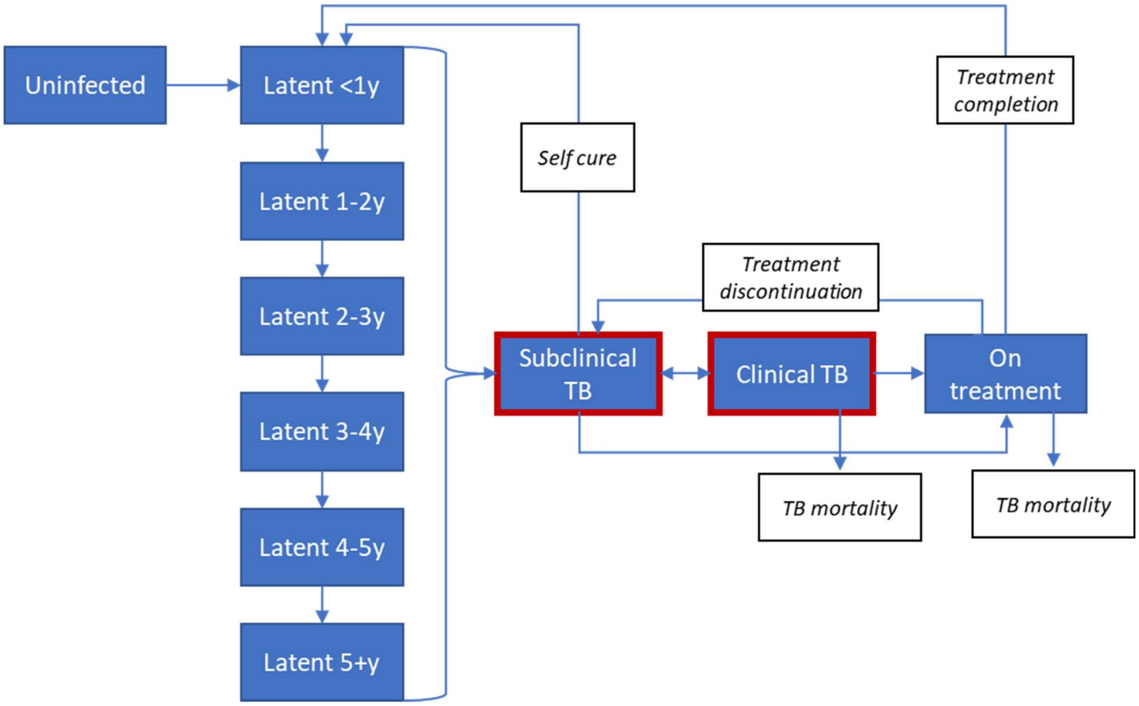

###### 1.1.11.2 Disease progression

The rate of developing tuberculosis disease following infection depended on an individual's time since infection with *Mtb* and their HIV/ART status. The rate was highest in the first year, falling each year over the subsequent five years, and then lowest from five years following infection. The rates in each year were higher for HIV+ART+ people than for HIV- people by a factor of *increased_develop_tb_rate_HIV2_vs_HIV0*, and for HIV+ART- people compared to HIV+ART+ people by a factor of *increased_develop_tb_rate_HIV1_vs_HIV2*. The rates were also higher for people with chronic conditions, by a factor of *increased_TB_development_chronic*, and for men than women by a factor of *increased_TB_development_male*. As an example, rates of developing TB in the first year following infection were set equal to:

*develop_tb_y1_rate_HIV0 * rate_adjust_HIV * rate_adjust_sex * rate_adjust_chronic*

where

- *rate_adjust_HIV* =
    - 1 if HIV-
    - *increased_develop_tb_rate_HIV2_vs_HIV0* if HIV+ART+
    - *increased_develop_tb_rate_HIV2_vs_HIV0 * increased_develop_tb_rate_HIV1_vs_HIV2* if HIV+ART-

- *rate_adjust_sex* =
  - 1 / (*prop_male* + (1 - *prop_male*) / *increased_TB_development_male*) if male
  - 1 / (*prop_male* * *increased_TB_development_male* + 1 - *prop_male*) if female
- *rate_adjust_chronic* =
  - 1 / ((prop_with_symp_hiv0/*1/2* * *proportion_symp_chronic_hiv0/12*) + (1 - (prop_with_symp_hiv0/1/2 * *proportion_symp_chronic_hiv0/12*)) / *increased_TB_development_chronic*) for people with a chronic condition
  - 1 / ((prop_with_symp_hiv0/1/2 * *proportion_symp_chronic_hiv0/12*) * *increased_TB_development_chronic* + 1 - (*prop_with_symp_hiv0/1/2* * *proportion_symp_chronic_hiv0/12*) for people without a chronic condition

Rates for subsequent years following infection were calculated in the same way.

The rate of developing disease also depended on the model year, being reduced by a factor of *decreased_tb_rates_late* in *change_TB_parameters_year* (see Section 1.1.11.8).

All simulated individuals who developed TB first developed subclinical TB (i.e. bacteriologically positive TB with no TB symptoms caused by TB, with or without TB symptoms caused by chronic conditions). From subclinical TB, people could progress to clinical TB (i.e. develop TB symptoms caused by TB), be diagnosed with TB and move to the treatment compartment (see Section 1.1.9), or self-cure.

Individuals with subclinical TB and no chronic conditions progress to clinical TB at a rate of:

- *rate_sub_to_clin_hiv1* if HIV+ART-
- *rate_sub_to_clin_hiv1* * *rate_sub_to_clin_hiv2_vs_hiv1* if HIV+ART+
- *rate_sub_to_clin_hiv1* * *rate_sub_to_clin_hiv2_vs_hiv1* * *rate_sub_to_clin _hiv0_vs_hiv2* if HIV-

These rates are increased by a factor of *relative_rate_sub_to_clin_chronic* for individuals with chronic conditions.

Individuals with subclinical TB and no chronic conditions self-cure at a rate of:

- *self_cure_rate_HIV0* if HIV-
- *self_cure_rate_HIV0* * *self_cure_rate_HIV2_vs_HIV0* if HIV+ART+
- *self_cure_rate_HIV0* * *self_cure_rate_HIV2_vs_HIV0* * *self_cure_rate_HIV1_vs_HIV2* if HIV+ART-

These rates are decreased by a factor of *relative_self_cure_rate_chronic* for individuals with chronic conditions.

Upon self-cure, individuals re-entered the first latent stage, resetting their time since infection back to zero. Individuals who progressed to disease again within 12 months of self-cure were not counted as having incident TB.

From clinical TB, people could regress to the subclinical TB stage, be diagnosed with TB and move to the treatment compartment (see Section 1.1.9), or die of TB.

Individuals with clinical TB and no chronic conditions regress to subclinical TB at a rate of:

- *rate_clin_to_sub_hiv0* if HIV-
- *rate_clin_to_sub_hiv0* * *rate_clin_to_sub_hiv2_vs_hiv0* if HIV+ART+
- *rate_clin_to_sub_hiv0* * *rate_clin_to_sub_hiv2_vs_hiv0* * *rate_clin_to_sub_hiv1_vs_hiv2* if HIV+ART-

These rates are decreased by a factor of *relative_rate_sub_to_clin_chronic* for individuals with chronic conditions.

Few data are available on the mortality rates of untreated clinical TB, particularly for PLWHIV. Using data from historical cohorts, Ragonnet *et al* estimated that the mortality rate of smear+ TB was 0.389 per year (95% credible interval 0.335 – 0.449)[24], with Richards *et al* arguing that the data used and estimated rate may better reflect mortality from clinical TB than smear+ TB[25]. These rates, estimated using data from populations before widespread availability of chemotherapy, are likely to be overestimates of the rates in populations where chemotherapy is available however. This is because the availability of treatment is likely to reduce the average severity of disease in the untreated population, with people with more severe disease having higher rates of starting treatment.

Calibrating the model to data and WHO estimates constrains the TB mortality rates in the calibrated parameter sets, and we therefore use wide, uninformative plausible ranges for the mortality rates.

Individuals with clinical TB and no chronic conditions die of TB at a rate of:

- *TB_mortality_rate_HIV1* if HIV+ART-
- *TB_mortality_rate_HIV1* * *TB_mortality_rate_HIV2_vs_HIV1* if HIV+ART+
- *TB_mortality_rate_HIV1* * *TB_mortality_rate_HIV2_vs_HIV1* * *TB_mortality_rate_HIV0_vs_HIV2* if HIV-

These rates are increased by a factor of *relative_TB_mortality_rate_chronic* for individuals with chronic conditions.

In the community-based intervention scenarios, individuals can also move from the subclinical and clinical TB compartments to the treatment compartment through community active case finding. Details are given in Section 1.1.13.1.

###### 1.1.11.3 Mycobacterium tuberculosis transmission

*Mtb* transmission occurred on a monthly time step in the model. For each susceptible or latent individual in the model, the probability of infection each month was calculated as:

$$1 - \prod_{c=0}^{1} \prod_{h=0}^{2} (1 - transmission\_prob \times w_c \times v_h \times reinfection\_relative\_risk)^{N_{ch}})$$

Where:

- $c$ indicates the symptom status of individuals with TB (0 = no symptoms caused by TB or chronic conditions, 1 = symptoms caused by chronic conditions only, 2 = symptoms caused by TB), $h$ indicates their HIV/ART class (0 = HIV negative, 1 = HIV positive, not on ART, 2 = HIV positive, on ART), and $N_{ch}$ indicates the number of people with TB with a particular symptom state and HIV/ART status.
- $w_c$ =
  - *reduced_transmission_sub* when c = 0
  - *(transmission_multiplier_chronic * 0.587 + reduced_transmission_sub * (1 – 0.587))* when c = 1
  - 1 when c = 2
- $v_h$ =
  - 1 when h = 0
  - *reduced_transmission_HIV1_vs_HIV2 * reduced_transmission_HIV2_vs_HIV0* when h = 1
  - *reduced_transmission_HIV2_vs_HIV0* when h = 2
- *reinfection_relative_risk* =
  - 1 if the individual was uninfected
  - *reinfection_relative_risk_HIV0* if they were HIV- and latently infected
  - *reinfection_relative_risk_HIV1* if they were HIV+ART- and latently infected
  - *reinfection_relative_risk_HIV2* if they were HIV+ART+ and latently infected.

###### 1.1.11.4 Test sensitivities and specificities

###### 1.1.11.4.1 Symptom screen

We assume that a symptom screen has a sensitivity of 100% for people with TB symptoms caused by TB and/or by chronic conditions, and for people visiting clinics seeking care for TB symptoms caused by transient conditions. We assume that all other people have a probability of reporting TB symptoms when asked (i.e. due to transient conditions) which depends on their HIV/ART status and whether they are a clinic attendee or a community screening participant. For screening conducted in the community, the probability of reporting symptoms due to transient conditions, *prob_transient_comm_hiv0/1/2* is calculated as:

- If HIV-negative: (*prop_with_symp_hiv0* - *prop_with_symp_hiv0* * *proportion_symp_chronic_hiv0*)/(1 - *prop_with_symp_hiv0* * *proportion_symp_chronic_hiv0*)
- If HIV-positive: (*prop_with_symp_hiv1/2* - *prop_with_symp_hiv1/2* * *proportion_symp_chronic_hiv12*)/(1 - *prop_with_symp_hiv1/2* * *proportion_symp_chronic_hiv12*)

For screening conducted in clinic, the probability of reporting symptoms, *prob_transient_clinic_hiv0/1/2* is calculated as:

- If not on ART: (*prob_transient_comm_hiv0/1* * *clinic_other_symp_hiv01* / (*prob_transient_comm_hiv0/1* * *clinic_other_symp_hiv01* + (1 - *prob_transient_comm_hiv0/1*)))
- If on ART: (*prob_transient_comm_hiv2* * *clinic_other_symp_hiv2* / (*prob_transient_comm_hiv2* * *clinic_other_symp_hiv2* + (1 - *prob_transient_comm_hiv2*)))
  - where *clinic_other_symp_HIV2* = (1 + (*clinic_other_symp_HIV01* - 1) * *clinic_other_HIV2_vs_HIV01*)

###### 1.1.11.4.2 Ability to produce sputum

Limited data are available on the proportion of people who are able to produce sputum samples in typical routine screening conditions in clinics, however it is likely to be substantially lower than the proportions achieved by prevalence surveys and research studies. One study in Botswana found that, before an intervention, only 44.1% of people starting ART were able to provide at least one sputum sample[26]. A study of clinic attendees in KwaZulu-Natal, South Africa found that only 50.4% were able to produce sputum (while this was a research study, the sputum collection procedures were in line with those used routinely in the clinics)[15]. The majority of people in these studies were not found to have TB, and we therefore assumed that 44.1 – 50.4% of people without TB are able to produce sputum (*produce_sputum_prob_tb*). A community prevalence survey in the same setting in

South Africa found little difference in the proportion of people able to produce sputum by sex, HIV/ART status or by reported TB symptoms[8], and we therefore assumed that the proportion of people able to produce sputum did not vary by these characteristics.

The proportion of people able to produce sputum in routine clinic conditions may be higher for people with TB. Following an intervention, the proportion of people able to produce sputum in the study in Botswana increased from 44.1% to 58.3%, and the overall yield of TB increased from 9.7% to 12.1%[26]. The prevalence of TB in people able to produce a sputum sample varied little between before and after the intervention. Using these data, we estimated that the proportion of people with TB who are able to produce sputum in routine clinic settings (*produce_sputum_prob_notb*) is at most 79.9% (assuming that all TB was detected following the intervention), and at least 48.4% (assuming that the prevalence of TB in people unable to produce sputum following the intervention was the same as in people able to produce sputum following the intervention). In line with people without TB, we assumed that the ability of people with TB to produce sputum does not vary by sex, HIV/ART status, or symptoms.

We assumed that the proportion of people with TB who are able to produce sputum could not be lower than the proportion of people without TB who are able to produce sputum.

###### 1.1.11.4.3 Xpert Ultra

Two Cochrane systematic reviews of the sensitivity and specificity of Xpert have been conducted, finding the sensitivity of Xpert Ultra to be 90.9% (95% CI 86.2% – 94.7%) in adults with presumptive pulmonary tuberculosis[27] and 69% (95% CI 57% – 80%) in people living with HIV "irrespective of signs or symptoms"[28]. We therefore simulated a range for the sensitivity of Xpert Ultra in people seeking care for TB symptoms (passive case finding) of 86.2% – 94.7%. The second review found no studies of screening in the general population, however the estimated sensitivity varied little between people living with HIV and non-hospitalised people in high risk groups, and we therefore assumed that the sensitivity was the same for HIV- people as for HIV+ people, and simulated a range for the sensitivity of Xpert Ultra used for active case finding in clinics or the community of 57% – 80%.

Systematic reviews have estimated that the specificity of Xpert Ultra is 95.6% (93.0% to 97.4%) in people with presumptive TB[27] and 98% (97% to 99%) in PLWHIV irrespective of signs and symptoms[28], with the latter estimate coming from only a single conference abstract. We simulated a wide plausible range of 93.0% - 99.0% for the specificity of Xpert Ultra in all populations, encompassing the ranges from both reviews.

###### 1.1.11.4.4 Xpert Ultra, saliva samples

A recent systematic review summarising studies estimating the sensitivity and specificity of different nucleic acid amplification tests (including Xpert Ultra) using different oral samples[29]. Only two studies estimated sensitivity for Xpert using saliva samples, finding sensitivities of 90% (95% CI 81% – 95%) using Xpert Ultra[30] and 38.5% (22.4% - 57.5%) using Xpert MTB/RIF[31]. All participants were sputum smear positive in the former study however, and 87% in the latter, and in the latter study participants were people being investigated for presumptive TB. These estimates may therefore be overestimates of the sensitivity when used for passive case finding in people unable to produce sputum. We therefore used the estimate from the systematic review for the sensitivity of Xpert MTB/RIF or Xpert Ultra using any oral specimen type, of 30% (95% CI 8% – 51%).

We assumed that Xpert Ultra using saliva samples has the same specificity as Xpert Ultra using sputum samples, giving a range of 93% - 99%, in line with the overall estimated specificity of nucleic acid amplification tests using oral samples[29]

###### 1.1.11.4.5 Chest X-ray

We assume that chest X-ray has a sensitivity of 94% (95% CI: 92% – 96%) and a specificity of 89% (95% CI: 85% – 92%), in line with WHO estimates of the sensitivity and specificity of chest X-ray (any abnormality) for screening for TB in the general population[32].

##### *1.1.11.5 Initial loss to follow up*

We assume that 11 – 18% of people are lost to follow up between diagnosis and starting treatment[33]. We assume that 50% of people diagnosed using Xpert Ultra in clinics or in community screening approaches and lost to follow up will be identified and start treatment at their next clinic visit, provided it occurs with two years of their diagnosis. We assume that the other 50% would need to be diagnosed again, e.g. because they attend a different clinic. We assume that people diagnosed following referral for X-ray or a doctor would need to be diagnosed again if lost to follow up, as we assume that in the majority of cases their local clinic would not be aware of their diagnosis.

##### *1.1.11.6 Treatment*

###### 1.1.11.6.1 People with TB (true-positives)

Treatment was assumed to last for 6 months in the model. Individuals successfully finishing treatment re-entered the latent stage with an effective time since infection of 6 months, reflecting the high rates of disease recurrence following treatment[34,35].

Individuals receiving TB treatment died of TB at a rate of *TB_mortality_rate_treatment_hiv0/12* and discontinued treatment at a rate of *TB_treatment_dropout_rate_hiv0/12*. Upon discontinuation, they returned to the subclinical TB compartment. Values of *TB_mortality_rate_treatment_hiv0/12* and *TB_treatment_dropout_rate_hiv0/12* were chosen to allow the model to match data on TB treatment outcomes for HIV+ and HIV- people in South Africa in 2020[36].

###### 1.1.11.6.2 People without TB (false-positives)

Individuals starting treatment who did not have TB are assumed to have the same rates of treatment discontinuation as people with TB. They are assumed to have no risk of becoming (re)infected with TB or developing disease whilst receiving treatment. After ending treatment (through successfully finishing or discontinuing), it is assumed that they are not at risk of developing TB without (re)infection.

##### *1.1.11.7 Prevalence of infection in 15-year olds*

An empirical study has estimated that the prevalence of *Mtb* infection in 12 – 17 year olds in KwaZulu-Natal province, South Africa in 2018 was 20.6%[37]. Upon being created at the age of 15 years, people in the model therefore set their state to latent with probability *prop_infect_15* = 0.206. The remaining people were assumed to be uninfected.

In calculating rates of progression to active disease in individuals with *Mtb* infections at the point of their creation at age 15 in the model, we assigned them a time of infection, *time_of_infection*, from a uniform distribution covering the 15 years before their creation. Their rate of disease progression following creation was then calculated using the same method as was used for people infected at ages ≥15 years. Progression to disease that occurred prior to the age of 15 was not included in the model.

##### *1.1.11.8 Changes in TB parameters over time*

To reflect secular trends not captured by other time varying parameters in the model (for instance, improvements in treatment coverage, and improvements in nutrition and housing), a change was modelled between *tb_rates_change_start_year* and *tb_rates_change_end_year*. Starting from *tb_rates_change_start_year*, the values of the simulated rate of *Mtb* transmission (*transmission_prob*) and the simulated rates of progression to TB disease following infection were reduced linearly, reaching values *relative_reduction_rates* times their original values in *tb_rates_change_end_year*.

##### *1.1.11.9 Calibration targets*

The model was calibrated to WHO estimates/data[36] for South Africa of:

- Estimated adult TB incidence and HIV+ TB incidence in 2005 and 2023. WHO estimates of overall incidence by year and incidence by age and HIV status in 2023 were used to estimate adult incidence, assuming that the ratio of TB incidence in adults compared to children was the same in 2005 as in 2023, and that the relative increase in incidence in HIV positive adults compared to HIV negative adults is the same as the relative increase in incidence in HIV positive children compared to HIV negative children.

- Estimated TB mortality and HIV+ TB mortality in 2005 and 2023. WHO estimates of overall (adult and child) mortality were adjusted to give estimated mortality in adults only, assuming that the proportion of people (overall and HIV positive) who develop TB who die of the disease is the same for adults as for children
- TB notification rates in adults in 2018 and 2023. We fitted the model to calibration targets ranging from the reported rates -10% to +10% in 2023, and -10% to +30% in 2018. A higher upper bound was used for 2018, as we did not explicitly model the scale up of active case finding in people on ART in clinics, and therefore numbers of false positive diagnoses in the model in 2018 may be too high.
- The ratio of number of people starting TB treatment to estimated TB incidence in 2023
- The proportion of adults starting TB treatment in 2023 who were living with HIV. Plausible bounds were calculated assuming that all adults who were not tested were or were not HIV positive.
- The proportion of TB deaths that occur in people who are on TB treatment in 2022, by HIV status. Calculated from TB treatment outcome data and estimated TB mortality numbers in 2022. Ranges were widened by ±30% for HIV- people, due to the unrealistically low uncertainty around HIV- TB mortality in WHO estimates.

The model was also calibrated to data from the 2017-2019 South Africa TB prevalence survey[3], fitting model output in 2018 to estimates of:

- The prevalence of bacteriologically positive TB in adult men and women
- The estimated prevalence of TB in HIV- and HIV+ adults
- The proportion of HIV- and HIV+ adults with prevalent bacteriologically positive TB who report TB symptoms
- The proportion of all HIV+ adults who report TB symptoms

Published estimates of TB prevalence in both HIV+ and HIV- adults (1734 and 900 per 100,000 respectively) are higher than the overall published estimate (852 per 100,000), due to a low TB prevalence in people with unknown HIV status. We therefore decreased the lower bounds estimated prevalence in HIV- and HIV+ adults, by multiplying them by the ratio of the adjusted overall TB prevalence in the population to the adjusted prevalence of TB in people with known HIV status.

The prevalence of TB symptoms in all participants and participants with bacteriologically positive TB in the South Africa national TB prevalence survey broken down by HIV status are shown in Table S1. The prevalence of reported symptoms was lower in people with unknown HIV status than in participants with known HIV status, both in all participants and in participants with bacteriologically positive TB. We therefore adjusted the lower bounds for symptom prevalence in HIV+ and HIV- people, assuming that all participants with unknown HIV status were HIV+ or HIV- respectively. The ranges may also be overestimates of the true proportions of people with TB symptoms due to selection bias, as only 66% of eligible people participated in the prevalence survey, and it is likely that participation was higher among people with symptoms. A community TB prevalence survey conducted in Kwa-Zulu Natal also found a much lower prevalence of TB symptoms: 8.5% among all participants, and 18.2% among participants with bacteriologically positive TB[8,15]. We therefore reduced the lower bounds by a further 40%. The reduction in the lower bounds for the estimates for people with bacteriologically positive TB also allows for the potential underestimation of the proportion who are symptom-screen negative resulting from only people with a positive chest X-ray or reporting symptoms being tested[7].

Table S1 Prevalence of reported symptoms by HIV status, overall and in people with bacteriologically positive TB, in the South Africa TB prevalence survey, and the ranges used to calibrate the model

| | HIV status | Prevalence of TB symptoms | | |
|---|---|---|---|---|
| | | Crude (95% CI) | Range adjusted for unknown HIV status | Final range used to calibrate model |
| All participants | HIV+ | 22.2% (21.0 – 23.4%) | 15.0 – 23.4% | 9.0 – 23.4% |
| | HIV- | 14.2% (13.7 – 14.6%) | 13.2 – 14.6% | 7.9 – 14.6%* |
| | HIV unknown | 12.0% (11.3 – 12.7%) | | |
| Participants with bacteriologically positive TB | HIV+ | 56.4% (42.3 – 69.7%) | 35.0 – 69.7% | 18.2 – 69.7% |
| | HIV- | 39.0% (30.7 – 47.7%) | 30.0 – 47.7% | 21.5% – 47.7% |
| | HIV unknown | 32.6% (19.1 – 48.5%) | | |

*The prevalence of TB symptoms in HIV- people in the overall population is used as an input parameter, not a calibration target. The prevalence of TB symptoms in HIV+ people in the model is a calibration target, as it is a weighted average of the prevalence in HIV+ people who are and are not on ART.

##### 1.1.12 Infection prevalence in child household contacts

Data were available from the AHCoS study on the prevalence of positive IGRA tests in child household contacts of people diagnosed with TB, and children living in households where no adults had been diagnosed with TB during the children's lifetimes ('controls')[38]. The mean age of children in the study/control households was 9 years.

The community force of infection each month over each the preceding nine years is tracked in the model in the form of a list *FOI_list* with 9 × 12 elements. The proportion of control children infected in 2023, *prop_infected_control*, is estimated in the model as:

$$1 - \prod_{y \in FOI\_list} 1 - child\_transmission\_multiplier \times FOI_y$$

Where *child_transmission_multiplier* is a parameter to account for the fact that 1) children may be exposed to a lower or higher force of infection from community exposure than adults, and 2) the FOI may be different in the study community than in South Africa as a whole.

People with bacteriologically positive TB are assumed to expose their child household contacts to an additional household force of infection. This is tracked each month over a period of up to nine years, using a list (*FOI_hh_list*) for each person with TB. Each month, the risk of household transmission is calculated as:

$$1 - (1 - transmission\_prob \times w_c \times v_h)^{infectiousness \times hh\_transmission\_multipler}$$

Where:

- *c* indicates their symptom status (0 = no symptoms caused by TB or chronic conditions, 1 = symptoms caused by chronic conditions only, 2 = symptoms caused by TB) and *h* indicates their HIV/ART class (0 = HIV negative, 1 = HIV positive, not on ART, 2 = HIV positive, on ART).
- $w_c$ =
  - *reduced_transmission_sub* when c = 0
  - *transmission_multiplier_chronic * 0.587* + *reduced_transmission_sub * (1 – 0.587)* when c = 1
  - 1 when c = 2
- $v_h$ =
  - 1 when h = 0
  - *reduced_transmission_HIV1* when h = 1
  - *reduced_transmission_HIV2* when h = 2

- infectiousness is an individual-level parameter, sampled at the start of the TB episode from a gamma distribution with mean 1 and dispersion parameter (*infectiousness_var*) k = 0.15[39,40].
- *hh_transmission_multiplier* reflects the increased exposure to the person with TB experienced by household members

When someone is diagnosed with TB in the model, the mean proportion of their child household contacts infected is calculated as:

$$1 - \prod_{y \in FOI\_list} 1 - child\_transmission\_multiplier \times FOI_y \prod_{y \in FOI\_hh\_list} 1 - FOI\_hh_y$$

The model was calibrated to the data from the AHCoS study on:

- The proportion of children infected in control households
- The proportion of children infected in child household contacts of HIV- people diagnosed with TB
- The relative risk of infection in child household contacts of people diagnosed with TB who are HIV+ART+ vs HIV-
- The relative risk of infection in child household contacts of people diagnosed with TB who are HIV+ART- vs HIV-
- The relative risk of infection in child household contacts of HIV+ART+ people diagnosed with TB who had symptom-screen negative vs symptom-screen positive TB at the time of diagnosis

##### 1.1.13 Interventions

All intervention approaches were simulated from the start of 2026, and run for a total of 10 years.

###### *1.1.13.1 Community-based*

Three community-based active case finding intervention approaches were simulated. We assumed that the interventions would be targeted at areas thought to have a high prevalence of TB. No data are available on how TB prevalence varies between communities across South Africa. We therefore used district level notification data from 2022[13] to provide a rough indication of how much higher prevalence may be in areas targeted for screening (compared to the country as a whole), and how this may vary with the proportion of the population targeted (Figure S3). Based on this relationship, we simulated scenarios covering 10%, 20%, 40%, 60%, and 80% of the adult population of South

Africa, targeted at areas with true prevalences of TB 2.27, 1.91, 1.56, 1.35, and 1.20 times higher respectively than the prevalence in the country as a whole.

Figure S3 Relationship between the proportion of the population of South Africa, and the notification rate in that group relative to the notification rate in South Africa as a whole. The leftmost point indicates the proportion of the population living in the district with the highest notification rate in 2022 and the notification rate in that district compared to the notification rate in South Africa as a whole. The second point shows data for the two highest notification rate districts, the third the three highest, etc.

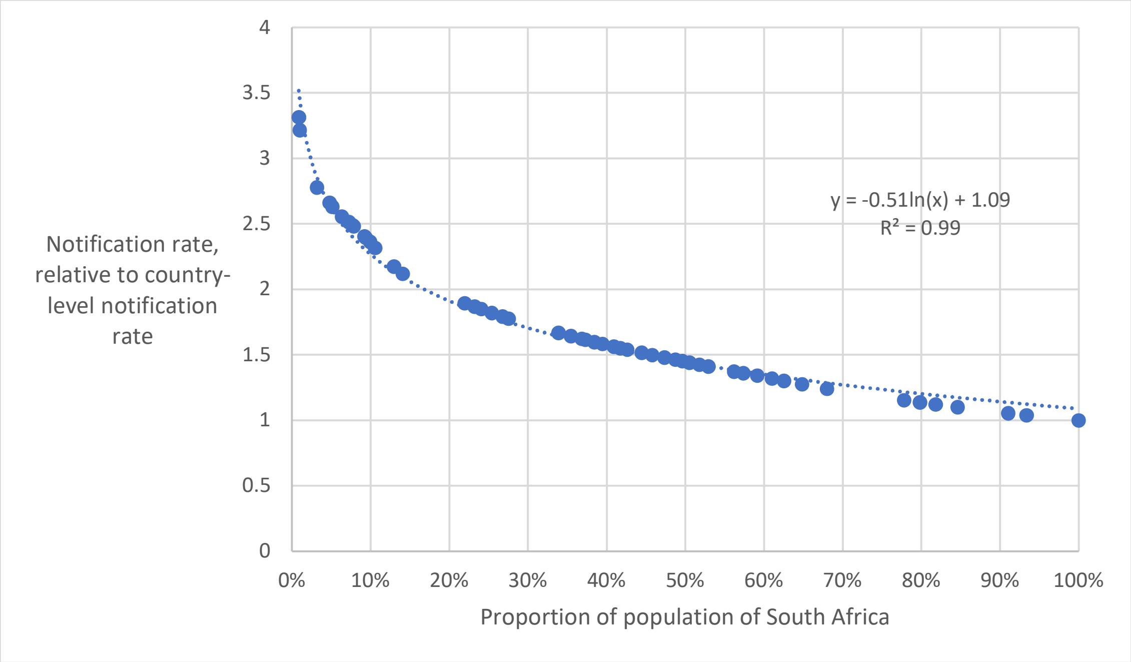

Each simulated individual was assigned a screening month at random at birth. Each month from the start of the intervention, people with the corresponding assigned screening month would be screened with probability:

- (*comm_int_cov* / (*TB_prevalence* + (1 - *TB_prevalence*) / *comm_RR_TB*))
  If they have TB and are not on TB treatment
- (*comm_int_cov* / (*comm_RR_TB* * *TB_prevalence* + 1 - *TB_prevalence*))
  If they do not have TB and are not on TB treatment

Where *TB_prevalence* is the prevalence of untreated TB in the model at the start of that month, *comm_int_cov* is the proportion of the population reached each year by the intervention, and *comm_RR_TB* is the true prevalence of TB in people screened.

Details of the simulated screening algorithms for the community-based approaches are shown in Supplementary material 2.

###### *1.1.13.2 Clinic-based*

Three clinic-based active case finding intervention approaches were simulated. These all included modifications to improve the active-screening pathway for HIV+ people on ART, and included the addition of active screening in some or all HIV- people and HIV+ people who are not on ART. The probability of diagnosis through passive case finding for people seeking care for TB symptoms remained the same as baseline, and was assumed to occur after any screening through active case finding had occurred. Full details of the simulated screening algorithms for the clinic-based approaches are shown in Supplementary material 2.

###### *1.1.13.2.1* Intensified targeted universal testing and treatment (TUTT)

TUTT is an approach where screening with Xpert Ultra is offered to all clinic attendees in three key risk groups: people living with HIV (PLWHIV), people reporting recent TB (within prior two years) and people reporting recent close contact with a person with TB (within past year)[41].

We considered it plausible that most PLWHIV who are not on ART either do not know their status, or would be reluctant to disclose it to healthcare workers as part of TB screening. We therefore assumed in the model that screening was limited to PLWHIV who are on ART only, unless they were also part of one of the other risk groups. We assumed that 95% of people on ART would report that they were HIV positive, with the remaining 5% not receiving screening. We tracked the date at which people last started TB treatment, with people who started treatment within the past two years (and who were no longer on treatment) eligible for testing. We assumed that 100% of people with recent TB would report their recent TB and receive screening.

Contacts of people with TB were not explicitly modelled, and were therefore selected at random from people visiting a clinic who did not meet the criteria for recent TB, with the probability of selection varying over time and with their true TB status. In line with the findings from a trial of the TUTT intervention, we assumed that the true prevalence of TB in recent close contacts is 40% lower than the true prevalence in people with recent TB[42]. Eight times as many people reported recent contact as reported recent TB in the trial, which we considered implausible in a scaled-up intervention. We therefore assumed that twice as many people would report being a recent contact as would report having recent prior TB.

We modelled a minimum time between receiving an Xpert through active screening of six months.

We assumed that people not on ART would be tested for HIV as part of the screening process, and would start ART if they tested positive.

###### 1.1.13.2.2 Intensified TUTT allowing saliva samples

This approach is the same as the intensified TUTT approach described above, except that the pathway for people unable to produce a sputum sample was different. We assumed that people unable to produce a sputum sample would produce a saliva sample, which would be tested with Xpert Ultra, with a reduced sensitivity as described in Section 1.1.11.4.4.

###### *1.1.13.2.3* Clinic-based symptom screening

The probability of symptom screening occurring for people on ART who had an Xpert Ultra attempted within the past year was increased from *symptom_screen_prob* (with baseline values of 0.5 – 0.7) to 0.75. The probability of symptom screening occurring for people not on ART was increased from 0 to 0.75. The proportion of symptomatic people on ART who have received an Xpert Ultra 6 months to 1 year ago and who are unable to produce sputum who see a doctor is increased from 65% to 75%.

##### 1.1.14 Calibration

The model was calibrated using history matching and emulation, using the R package *hmer*[43]. As we were modelling an entire country, the model was treated as deterministic, with results for each parameter set averaged over 200 runs. Calibration was performed iteratively as the model was developed, with the ranges used for parameters that are not informed by data or expert opinion (i.e. those where the only constraints are those imposed by calibrating the model to data) modified each time the model was recalibrated to ensure that a) ranges were wide enough to contain all possible full fitting points and b) ranges were not unnecessarily wide, as this would increase the number of waves required for calibration and increase the risk of full-fitting space being incorrectly ruled out.

A number of input parameters were transformed, to allow the distribution of the values of other input parameters or calibration targets in the fitting points to better reflect the desired distributions.

The model used the parameter *transmission_prob* as the baseline rate of transmission between each infectious and susceptible person (see Section 1.1.11.3), with *transmission_prob* multiplied by *reduced_transmission_sub* for infectious people with subclinical TB and no chronic condition. The value of *transmission_prob* was calculated from the input parameter *baseline_infection_rate* as *transmission_prob* = 1 / *baseline_infection_rate*. This transformation was used because, when all else is equal, there is a non-linear relationship between the values of *transmission_prob* and the values of *reduced_transmission_sub* that give rise to any particular average rate of transmission between each infectious and susceptible person (Figure S4a). As *hmer* treats the distributions of all input parameters as uniform, this would result in the proportion of the multi-dimensional fully fitting parameter space that contains smaller values of *reduced_transmission_sub* being considerably larger than the proportion that contains larger values, and in smaller values of *reduced_transmission_sub* being greatly overrepresented in the final full fitting parameter sets. Using the transformed variable results in a linear relationship between the values of *transmission_prob* and the values of *reduced_transmission_sub* that give rise to any particular average rate of transmission between each infectious and susceptible person (Figure S4b), and prevents this source of bias.

Figure S4 Relationship between the values of a) *transmission_prob* and *reduced_transmission_sub* and b) *baseline_infection_rate* and *reduced_transmission_sub* that, all else being equal, give rise to any particular average rate of transmission between each infectious and susceptible person

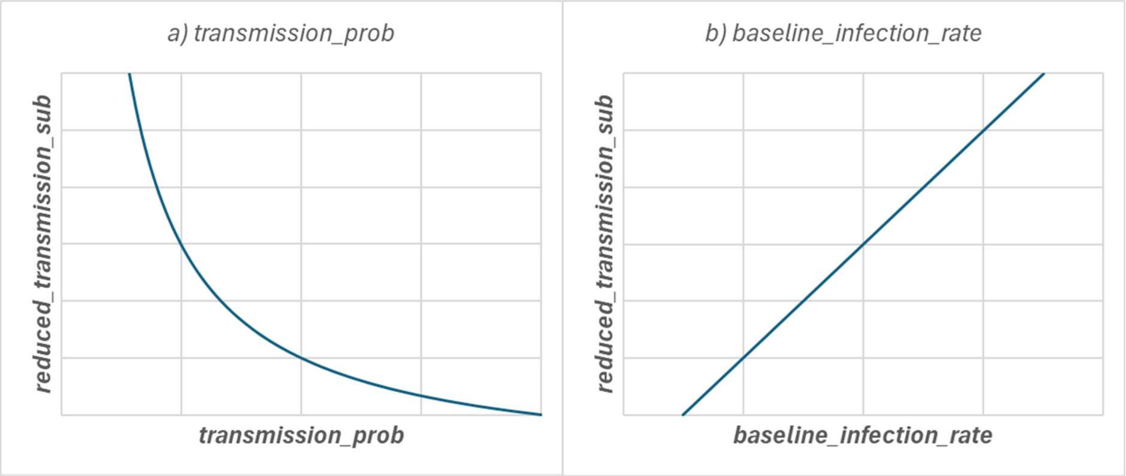

The parameters controlling the rates of starting ART in men and women in different time periods were also transformed. This was because the relationship between the ART start rates and the level of ART coverage was non-linear, with small changes in the values of the rates having a larger impact on ART coverage at the bottom of the ranges of the rates than at the top. This resulted in ART coverage being concentrated at the tops of the ranges in initial calibration attempts. The relationships between the start rates and ART coverage were determined using the fitting points from those early calibration attempts and used to identify transformations that would result in improved distributions of ART coverage in the final calibration. The transformations used were:

ART_start_rate_early_m_annual = 0.054 * exp(1.068 * ART_start_rate_early_m_annual_input)

ART_start_rate_early_f_annual = 0.062 * exp(1.61 * ART_start_rate_early_f_annual_input)

ART_start_rate_late_m_annual = 0.021 * exp(2.86 * ART_start_rate_late_m_annual_input)

ART_start_rate_late_f_annual = 0.0193 * exp(3.44 * ART_start_rate_late_f_annual_input) – 0.019

A total of 64 input parameters were varied during final calibration (Table S3), with the model calibrated to 42 calibration targets (Table S5). A total of 10 waves of calibration were run, with the first full fitting points found from wave 4. The yield of full fitting points was sufficiently high in wave 10 (18% full fitting), so we generated new points from wave 10 in sets of 2000 until a minimum of 1000 full fitting points were found. This gave us a total of 1041 full fitting points that were used in the analysis.

The model was run at 10 × the number of input parameters = 640 parameter sets to provide the training data for the emulators each wave. A further 5 × the number of input parameters = 320 parameter sets were used to validate and adjust the emulators, using the *hmer* package function *diagnostic_pass*. Finally, the emulators were validated using a further 9 hand fitted points near the edges of the full fitting space (i.e. at the lower or upper ends of the ranges for one or more input parameter, and/or at the lower or upper ends of the ranges for one or more calibration target). If any of the emulators ruled out any of these full fitting points, then their uncertainties (sigmas) were iteratively increased by 5% until the point was no longer ruled out.

##### 1.1.15 Input parameters

###### *1.1.15.1 Fixed input parameters*

Table S2 Descriptions and values of input parameters that were not varied

| **Parameter name** | **Description** | **Value** | **Source** |
|---|---|---|---|
| **Demography** | | | |
| min_age | Minimum age in model | 15 | Majority of *Mtb* transmission from people aged 15+, and interventions are focused on screening adults |
| max_age | Maximum age in model | 80 | <1.3% of adult population aged >80 in South Africa in 2021 |
| min_background_mort_age_m | Minimum age from which non-TB and non-HIV mortality occurs in men in the model | 40 | Values chosen to give an approximate fit to the age distribution by sex in South Africa in 2021[2] |
| min_background_mort_age_f | Minimum age from which non-TB and non-HIV mortality occurs in women in the model | 45 | |
| background_mort_m | Annual non-HIV and non-TB mortality rate in men above min_background_mort_age_m | 0.0458 | |
| background_mort_f | Annual non-HIV and non-TB mortality rate in women above min_background_mort_age_f | 0.0355 | |
| prop_male | Proportion of newly created people who are male | 0.476 | Demographic estimates[2] |
| **HIV and ART** | | | |

| | | | |
|---|---|---|---|
| hiv_intro_year | Year HIV first introduced into the model | 1995 | Chosen to allow the model to fit to trends in HIV prevalence and ART coverage, and in TB incidence |
| art_intro_year | Year ART first introduced into the model | 2010 | |
| HIV1_mortality_rate_annual | Annual non-TB HIV mortality rate in HIV+ people not on ART | 0.00225 | See Section 1.1.10 |
| HIV2_mortality_rate_annual | Annual non-TB HIV mortality rate in HIV+ people on ART | 0.00108 | |
| hiv_inc_change_year1_start | Year first change in HIV incidence starts | 2000 | Chosen to allow the model to fit to trends in HIV prevalence and ART coverage |
| hiv_inc_change_year1_end | Year first change in HIV incidence ends | 2020 | |
| hiv_inc_change_year2_start | Year second change in HIV incidence starts | 2023 | |
| hiv_inc_change_year2_end | Year second change in HIV incidence ends | 2040 | |
| ART_start_rate_change_year | Year change in ART start rates occurs | 2017 | |
| abs_increased_prop_symp_chronic_hiv | Absolute increase in the proportion of TB symptoms that are due to chronic conditions in people who are HIV+ | 0.1 | Expert opinion, see Section 1.1.7.2 |
| **Clinic visiting** | | | |
| clinic_visit_alpha_m_hiv01 | Parameters controlling the distributions for the individual-level rates at which men and women who are on or not on ART visit clinics, for reasons other than seeking care for TB symptoms | 0.149 | See section 1.1.9.2.1.1 |
| clinic_visit_lambda_m_hiv01 | | 0.186 | |
| clinic_visit_alpha_m_hiv2 | | 1.75 | |
| clinic_visit_lambda_m_hiv2 | | 0.444 | |
| clinic_visit_alpha_f_hiv01 | | 0.368 | |
| clinic_visit_lambda_f_hiv01 | | 0.198 | |
| clinic_visit_alpha_f_hiv2 | | 1.93 | |

| | | | |
|---|---|---|---|
| clinic_visit_lambda_f_hiv2 | | 0.416 | |
| **TB diagnosis and treatment** | | | |
| TB_mortality_rate_treatment_hiv0 | Rate of dying of TB on TB treatment if HIV- negative, per month | 0.0131 | Data on TB treatment outcomes for HIV+ and HIV- people in South Africa in 2020[36] |
| TB_treatment_dropout_rate_hiv0 | Rate of stopping TB treatment if HIV- negative, per month | 0.0263 | |
| TB_mortality_rate_treatment_hiv12 | Rate of dying of TB on TB treatment if HIV+ negative, per month | 0.0169 | |
| TB_treatment_dropout_rate_hiv12 | Rate of stopping TB treatment if HIV+ negative, per month | 0.0180 | |
| **Mtb transmission and TB development** | | | |
| reinfection_relative_risk_HIV0 | Relative risk of reinfection compared to initial infection, in HIV-, HIV+HIV-, and HIV+ART+ people | 0.21 | Andrews *et al* 2012[44] |
| reinfection_relative_risk_HIV1 | | 0.75 | Dowdy and Chaisson, 2009[45] |
| reinfection_relative_risk_HIV2 | | 0.48 | Intermediate between protection in HIV- and HIV+ART- |
| develop_tb_y1_rate_HIV0_annual | Annual rate of developing TB in HIV- people <1 year, 1-2 years, 2-3 years, 3-4 years, 4-5 years, and >5 years following last infection | 0.0866 | Kasaie 2014[46] |
| develop_tb_y2_rate_HIV0_annual | | 0.0355 | |
| develop_tb_y3_rate_HIV0_annual | | 0.0112 | |
| develop_tb_y4_rate_HIV0_annual | | 0.0074 | |
| develop_tb_y5_rate_HIV0_annual | | 0.0024 | |
| develop_tb_reactivation_rate_HIV0_annual | | 0.0005 | |
| prop_infect_15 | Proportion of people with *Mtb* infection at their creation at age 15 | 0.206 | Mzembe 2020[37] |

| | | | |
|---|---|---|---|
| infectiousness_var | Individual-level infectiousness is sampled from a gamma distribution with mean 1 and dispersion parameter k = infectiousness_var | 0.15 | McCreesh and White 2018[39] and Smith *et al* 2023[40] |
| infection_seed_proportion | Proportion of people initially seeded with Mtb infection | 0.7 | NA |
| tb_seed_proportion | Proportion of people initially seeded with TB disease | 0.001 | NA |

##### *1.1.15.2 Input parameters varied during calibration*

Table S3 Descriptions and ranges of input parameters that were varied during calibration

| **Parameter name** | **Description** | **Range permitted for fitting points** | **Range (median) in fitted parameter sets** | **Type/source** |
|---|---|---|---|---|
| **HIV/ART** | | | | |
| hiv_inc_early_f_annual | Parameters controlling HIV incidence in men and women in different time periods (see Section 1.1.10) | 0 – 0.05 | 0.019 – 0.034 (0.025) | Constrained during calibration |
| hiv_inc_early_m_annual | | 0 – 0.05 | 0.013 – 0.025 (0.019) | |
| hiv_inc_mid_f_annual | | 0 – 0.04 | 0.0042 – 0.011 (0.0072) | |
| hiv_inc_mid_m_annual | | 0 – 0.03 | 0.00069 – 0.0063 (0.0038) | |
| hiv_inc_f_late_reduction | | 0.2 – 1 | 0.205 – 0.98 (0.508) | |

| | | | | |
|---|---|---|---|---|
| hiv_inc_m_late_reduction | | 0.2 – 1 | 0.408 – 0.999 (0.665) | |
| art_intro_year | Year ART is introduced into the model | 2005 – 2010 | 2005.0 – 2010.0 (2007.06) | Chosen to allow the model to fit to data on ART coverage |
| ART_start_rate_early_m_annual_input | Parameters controlling ART start rates (see Section 1.1.13.2.3 for details) | 0 – 1 | 0.014 – 0.77 (0.44) | Constrained during calibration |
| ART_start_rate_early_f_annual_input | | 0 – 1 | 0.131 – 0.878 (0.53) | |
| ART_start_rate_late_m_annual_input | | 0 – 1 | 0.32 – 0.71 (0.53) | |
| ART_start_rate_late_f_annual_input | | 0 – 1 | 0.51 – 0.83 (0.69) | |
| ***Mycobacterium tuberculosis*** **transmission** | | | | |
| baseline_infection_rate | The baseline rate of transmission between each infectious and susceptible person in the model is a function of 1 / baseline_infection_rate. See sections Mycobacterium tuberculosis transmission1.1.11.3 and 1.1.13.2.3 for details | 3000 – 30000 | 8260.7 – 15075 (11450) | Constrained during calibration |
| reduced_transmission_sub | Rate of transmission from people with subclinical TB and no chronic condition, relative to people with clinical TB | 0.2 – 1.3 | 0.53 – 1.2 (0.83) | Expert opinion |
| transmission_multiplier_chronic | Parameter controlling rate of transmission from people subclinical TB and a chronic condition (see Section 1.1.7.2) | 1.1 – 1.5 | 1.1 – 1.4 (1.3) | Expert opinion |
| reduced_transmission_HIV1_vs_HIV2 | Rate of *Mtb* transmission from HIV+ART- people relative to HIV+ART+ people | 0 – 1 | 0.31 – 0.93 (0.62) | Assume that HIV+ART- people cannot be more infectious than HIV+ART+ people |

| | | | | |
|---|---|---|---|---|
| reduced_transmission_HIV2_vs_HIV0 | Rate of *Mtb* transmission from HIV+ART+ people relative to HIV- people | 0 – 1 | 0.29 – 0.98 (0.68) | Assume that HIV+ART+ people cannot be more infectious than HIV- people |
| hh_trans_rate_multiplier | Parameter controlling rate of transmission to child household contacts | 0 – 2000 | 97 – 433 (232) | Constrained during calibration |
| child_transmission_multiplier | Parameter controlling rate of community transmission to children | 0 – 0.4 | 0.097 – 0.32 (0.21) | Constrained during calibration |
| **Progression to disease** | | | | |
| increased_develop_tb_rate_HIV2_vs_HIV0 | Increased rate of developing TB following infection in HIV+ART+ people relative to HIV- people | 1.47 – 10.8 | 1.66 – 4.25 (2.88) | Gupta *et al* 2012[18] and Gatechompol *et al* 2022[19] |
| increased_develop_tb_rate_HIV1_vs_HIV2 | Increased rate of developing TB following infection in HIV+ART- people relative to HIV+ART+ people | 1.89 – 4.76 | 2.48 – 4.6 (3.47) | Suthar *at al* 2015[20] |
| increased_TB_development_male | Increased rate of developing TB following infection in men compared to women | 1 – 4 | 1.14 – 2.87 (1.88) | Constrained during calibration |
| increased_TB_development_chronic | Increased rate of developing TB following infection in people with chronic conditions that cause TB symptoms | 1.3 – 1.9 | 1.33 – 1.83 (1.59) | Expert opinion |
| tb_rates_change_start_year | Years between which rates of *Mtb* transmission and TB disease development reduce, and magnitude of reduction (see Section 1.1.11.8) | 2000 – 2007 | 2001.9 – 2006.9 (2004.8) | Ranges chosen to allow model to fit to trends in TB incidence over time |
| tb_rates_change_end_year | | 2007 – 2015 | 2008.7 – 2015.0 (2012.4) | |
| relative_reduction_rates | | 0.70 – 1.00 | 0.86 – 0.98 (0.91) | |
| **Disease progression and regression** | | | | |
| self_cure_rate_HIV0 | Rate of self cure per year for HIV- people with asymptomatic TB | 0 – 0.8 | 0.068 – 0.28 (0.17) | Constrained during calibration |

| | | | | |
|---|---|---|---|---|
| self_cure_rate_HIV1_vs_HIV2 | Rate of self cure for HIV+ART- people with asymptomatic TB, relative to HIV+ART+ people | 0 – 1 | 0.006 – 0.92 (0.46) | Assumed that rate of self cure must be lower in HIV+ART- people than HIV+ART+ people |
| self_cure_rate_HIV2_vs_HIV0 | Rate of self cure for HIV+ART+ people with asymptomatic TB, relative to HIV- people | 0 – 1 | 0.063 – 0.96 (0.54) | Assumed that rate of self cure must be lower in HIV+ART+ people than HIV- people |
| relative_self_cure_rate_chronic | Rate of self cure in people with chronic conditions that cause TB symptoms, relative to people with no chronic conditions that cause TB symptoms | 0.53 – 0.77 | 0.54 – 0.76 (0.65) | Expert opinion |
| TB_mortality_rate_HIV0_vs_HIV2 | TB mortality rate in HIV- people relative to HIV+ART+ people | 0 – 1 | 0.24 – 0.86 (0.55) | Assumed that TB mortality rate must be lower in HIV- people than HIV+ART+ people |
| TB_mortality_rate_HIV1 | TB mortality rate per year in HIV+ART- people | 0.3 – 4 | 0.97 – 2.7 (1.7) | Constrained during calibration |
| TB_mortality_rate_HIV2_vs_HIV1 | TB mortality rate in HIV+ART+ people relative to HIV+ART- people | 0 – 1 | 0.17 – 0.69 (0.41) | Assumed that TB mortality rate must be lower in HIV+ART+ people than HIV+ART- people |
| relative_TB_mortality_rate_chronic | TB mortality rate in people with chronic conditions that cause TB symptoms, relative to people with no chronic conditions that cause TB symptoms | 1.3 – 1.9 | 1.3 – 1.8 (1.5) | Expert opinion |
| rate_sub_to_clin_hiv0_vs_hiv2 | Rate of developing TB symptoms caused by TB in HIV- people relative to HIV+ART+ people | 0 – 1 | 0.27 – 0.8 (0.53) | Assume that rate must be lower in HIV- people relative to HIV+ART+ people |
| rate_sub_to_clin_hiv1 | Rate of developing TB symptoms caused by TB, per year, in HIV+ART- people | 0 – 15 | 4.8 – 13.1 (8.0) | Wide plausible range, as few data available (see Section 1.1.7.1) |
| rate_sub_to_clin_hiv2_vs_hiv1 | Rate of developing TB symptoms caused by TB in HIV+ART+ people relative to HIV+ART- people | 0 – 1 | 0.37 – 0.97 (0.6) | Assume that rate must be lower in HIV+ART+ people relative to HIV+ART- people |
| relative_rate_sub_to_clin_chronic | Rate of developing TB symptoms caused by TB in people with chronic conditions that cause TB symptoms relative to people without chronic conditions that cause TB symptoms | 1.3 – 1.9 | 1.3 – 1.9 (1.6) | Expert opinion |

| | | | | |
|---|---|---|---|---|
| rate_clin_to_sub_hiv0 | Rate of losing TB symptoms caused by TB, per year, in HIV- people | 0 – 12 | 2.95 – 11.7 (7.84) | Wide plausible range, as few data available (see Section 1.1.7.1) |
| rate_clin_to_sub_hiv2_vs_hiv0 | Rate of losing TB symptoms caused by TB in HIV+ART+ people relative to HIV- people | 0 – 1 | 0.32 – 0.97 (0.65) | Assume that rate must be lower in HIV+ART+ people relative to HIV- people |
| rate_clin_to_sub_hiv1_vs_hiv2 | Rate of losing TB symptoms caused by TB in HIV+ART- people relative to HIV+ART+ people | 0 – 1 | 0.059 – 0.95 (0.53) | Assume that rate must be lower in HIV+ART- people relative to HIV+ART+ people |
| relative_rate_clin_to_sub_chronic | Rate of losing TB symptoms caused by TB in people with chronic conditions that cause TB symptoms relative to people without chronic conditions that cause TB symptoms | 0.526 – 0.769 | 0.54 – 0.74 (0.63) | Expert opinion |
| **Symptoms and clinic visiting** | | | | |
| prop_with_symp_hiv0 | Proportion of HIV- people reporting TB symptoms | 0.079 – 0.146 | 0.08 – 0.12 (0.098) | South Africa TB prevalence survey[3], adjusted as described in Section 1.1.11.9 |
| prop_with_symp_input_hiv1 | Parameter controlling the proportion of HIV+ART- people reporting symptoms (see Section 1.1.9.2.2) | 0 – 1 | 0.015 – 0.26 (0.1) | Constrained during calibration |
| prop_with_symp_input_hiv2 | Parameter controlling the proportion of HIV+ART+ people reporting symptoms (see Section 1.1.9.2.2) | 0 – 1 | 0.028 – 0.65 (0.27) | Constrained during calibration |
| proportion_symp_chronic_hiv0 | Proportion of TB symptoms in people without TB that result from chronic conditions (opposed to transient conditions) | 0.3 – 0.7 | 0.30 – 0.54 (0.42) | Expert opinion |
| clinic_TBsymp_HIV0 | Rate of seeking care for TB symptoms caused by TB in HIV- people | 0 – 6 | 2.2 – 4.6 (3.2) | Constrained during calibration |
| clinic_TBsymp_HIV1 | Rate of seeking care for TB symptoms caused by TB in HIV+ART- people | 0 – 6 | 1.4 – 4.2 (2.4) | Only partially constrained during calibration, as most calibration targets not broken down by ART status and people on ART can also start treatment through active case finding. Maximum rate of 6 per year therefore set |
| clinic_TBsymp_HIV2 | Rate of seeking care for TB symptoms caused by TB in HIV+ART+ people | 0 – 6 | 0.45 – 5.6 (2.4) | |

| | | | | |
|---|---|---|---|---|
| clinic_TBsymp_adjust_chronic | Rate of seeking care for TB symptoms in people whose TB symptoms are caused by chronic conditions only, compared to people with TB symptoms caused by TB | 0.5 – 2 | 0.51 – 1.5 (0.93) | Expert opinion |
| clinic_TBsymp_adjust_transient | Rate of seeking care for TB symptoms in people whose TB symptoms are caused by transient conditions only, compared to people with TB symptoms caused by chronic conditions | 0 – 1 | 0.01 – 0.97 (0.4) | Considered plausible that the rate of seeking care for TB symptoms caused by transient conditions would be lower than the rate of seeking care for TB symptoms caused by chronic conditions |
| clinic_visit_adjust | Parameter scaling the rates of clinic visiting for reasons other than seeking care for TB symptoms | 0.5 – 1.5 | 0.88 – 1.1 (1.0) | Constrained during calibration |
| clinic_other_symp_HIV01 | Increased rate of clinic visiting for reasons other than seeking care for TB symptoms, in people not on ART with TB symptoms | 1 – 3 | 1.0 – 2.1 (1.5) | Constrained during calibration |
| clinic_other_tb_HIV01 | Increased rate of clinic visiting for reasons other than seeking care for TB symptoms, in people not on ART with TB | 1 – 15 | 1.1 – 4.9 (3.1) | Constrained during calibration |
| clinic_other_HIV2_vs_HIV01 | Reduction in increased rates of clinic visiting for reasons other than seeking care for TB symptoms, for people on ART compare to people not on ART | 0 – 1 | 0.048 – 0.77 (0.41) | Considered plausible that it would be lower in people on ART, due to the higher rate of clinic visiting for routine HIV care |
| **Diagnosis** | | | | |
| passive_diagnosis_prob | Probability of diagnosis through passive case finding for people seeking care for TB symptoms | 0.25 – 0.75 | 0.36 – 0.71 (0.49) | McCarthy *et al* 2018[47] and Foster *et al* 2015[48] |
| produce_sputum_prob_tb | Probability that a person with TB will be able to produce sputum | 0.48 – 0.80 | 0.56 – 0.75 (0.64) | See Section 1.1.11.4.2 |
| xpert_sens_passive | Sensitivity of Xpert Ultra in people seeking care for TB symptoms (passive case finding) | 0.862 – 0.947 | 0.58 – 0.8 (0.69) | Zifodya 2021[27] |

| | | | | |
|---|---|---|---|---|
| xpert_sens_active | Sensitivity of Xpert Ultra used for active case finding | 0.57 – 0.80 | 0.86 – 0.94 (0.9) | Shapiro 2021[28] |
| symptom_screen_prob | Probability of symptom screening occurring during active case finding (see Section 1.1.9.4) | 0.5 – 0.7 | 0.51 – 0.7 (0.61) | Expert opinion |
| xray_sens | Sensitivity of chest X-ray | 0.92 – 0.96 | 0.92 – 0.95 (0.94) | World Health Organization 2021[32] |
| iltfu | Proportion of people lost to follow up between diagnosis and starting treatment | 0.11 – 0.18 | 0.11 – 0.17 (0.13) | Mwansa-Kambafwile 2020[33] |
| produce_sputum_prob_notb | Probability that a person without TB will be able to produce sputum | 0.44 – 0.50 | 0.44 – 0.5 (0.47) | Mathebula 2020[26] |
| xpert_spec | Specificity of Xpert Ultra | 0.93 – 0.99 | 0.966 – 0.99 (0.983) | Zifodya 2021[27] and Shapiro 2021[28] |
| xray_spec | Specificity of chest X-ray | 0.85 – 0.92 | 0.85 – 0.92 (0.88) | World Health Organization 2021[32] |

###### *1.1.15.3 Input parameters varied in future projections*

Table S4 Descriptions and ranges of input parameters that were varied during the intervention runs

| **Parameter name** | **Description** | **Range and distribution** | **Source** |
|---|---|---|---|
| xpert_saliva_sens | Sensitivity of Xpert Ultra with a saliva sample | Uniform distribution 0.08 – 0.51 | Wang 2023[29] |

##### 1.1.16 Calibration targets table

Table S5 Calibration targets, ranges, and fitting values

| Target description | Range | Range (median) in fitting points | Source |
|---|---|---|---|
| **HIV prevalence and ART coverage** | | | |
| Male adult HIV prevalence in 2000 | 0.061 – 0.13 | 0.061 – 0.11 (0.082) | UNAIDS estimates[21] |
| Female adult HIV prevalence in 2000 | 0.087 – 0.17 | 0.087 – 0.15 (0.12) | |
| Male adult HIV prevalence in 2023 | 0.11 – 0.13 | 0.11 – 0.13 (0.12) | |
| Female adult HIV prevalence in 2023 | 0.19 – 0.23 | 0.19 – 0.23 (0.21) | |
| Male adult HIV prevalence in 2030 | 0.077 – 0.14 | 0.085 – 0.12 (0.1) | Estimates from the Thembisa model[23], with widened ranges (see Section 1.1.10.2) |
| Female adult HIV prevalence in 2030 | 0.14 – 0.26 | 0.17 – 0.23 (0.2) | |
| Male adult ART coverage in 2016 | 0.37 – 0.67 | 0.41 – 0.65 (0.54) | UNAIDS estimates[21] |
| Female adult ART coverage in 2016 | 0.46 – 0.82 | 0.47 – 0.81 (0.68) | |
| Male adult ART coverage in 2023 | 0.65 – 0.77 | 0.66 – 0.77 (0.72) | |
| Female adult ART coverage in 2023 | 0.75 – 0.88 | 0.77 – 0.88 (0.84) | |
| Ratio of ART coverage in 2019 vs 2023 in adult men | 0 – 1 | 0.8 – 0.97 (0.89) | Target used to exclude otherwise fitting runs where ART coverage declines following 2023 |
| Ratio of ART coverage in 2019 vs 2023 in adult women | 0 – 1 | 0.83 – 0.99 (0.92) | Target used to exclude otherwise fitting runs where ART coverage declines following 2023 |

| TB incidence, mortality, and notifications | | | |
|---|---|---|---|
| TB incidence/100,000 in 2005 | 598 – 3423 | 946 – 1990 (1330) | WHO data and estimates[36] |
| HIV+ TB incidence/100,000 in 2005 | 385 – 2450 | 574 – 1330 (889) | |
| TB incidence/100,000 in 2023 | 358 – 841 | 411 – 598 (508) | |
| HIV+ TB incidence/100,000 in 2023 | 190 – 449 | 190 – 315 (245) | |
| TB mortality/100,000 in 2005 | 379 – 934 | 417 – 893 (628) | |
| HIV+ TB mortality/100,000 in 2005 | 275 – 814 | 348 – 801 (548) | |
| TB mortality/100,000 in 2023 | 65 – 187 | 100 – 187 (155) | |
| HIV+ TB mortality/100,000 in 2023 | 19 – 139 | 65.5 – 136 (102) | |
| TB notifications/100,000 in 2018 | 448 – 647 | 449 – 615 (511) | |
| TB notifications/100,000 in 2023 | 379 – 464 | 381 – 464 (445) | |
| Ratio of TB notifications to incidence in 2023 | 0.54 – 1.30 | 0.73 – 1.1 (0.88) | |
| Proportion of people starting TB treatment in 2023 who are HIV+ | 0.45 – 0.61 | 0.47 – 0.61 (0.58) | |
| Proportion of HIV+ people starting TB treatment in 2023 who are on ART | 0.80 – 0.95 | 0.8 – 0.89 (0.83) | |
| Proportion of TB mortality in HIV- people in 2022 that occurs in people on TB treatment | 0.17 – 0.35 | 0.17 – 0.35 (0.25) | |

| | | | |
|---|---|---|---|
| Proportion of TB mortality in HIV+ people in 2022 that occurs in people on TB treatment | 0.14 – 0.91 | 0.14 – 0.25 (0.15) | |
| **Data from 2018 South African TB prevalence survey** | | | |
| TB prevalence/100,000 in men 2018 | 835 – 1352 | 835 – 1350 (1018) | South Africa TB prevalence survey[3] (see Section 1.1.11.9 for details) |
| TB prevalence/100,000 in women 2018 | 494 – 855 | 495 – 842 (565) | |
| TB prevalence/100,000 in HIV+ people | 996 – 2249 | 997 – 1918 (1213) | |
| TB prevalence/100,000 in HIV- people | 565 – 1108 | 565 – 1007 (684.8) | |
| Prevalence of self-reported TB symptoms in HIV- people with TB | 0.22 – 0.48 | 0.22 – 0.38 (0.27) | |
| Prevalence of self-reported TB symptoms in HIV+ people with TB | 0.18 – 0.70 | 0.39 – 0.68 (0.55) | |
| Proportion of HIV+ people reporting TB symptoms | 0.090 – 0.23 | 0.090 – 0.23 (0.14) | |
| **Clinic visiting** | | | |
| Mean number of clinic visits per adult in 2022 | 1.50 – 1.90 | 1.51 – 1.87 (1.68) | See Section 1.1.9.5 |
| Prevalence of symptoms in clinic attendees in 2022 | 0 – 0.30 | 0.16 – 0.30 (0.24) | |

| | | | |
|---|---|---|---|
| Increased prevalence of TB in clinic attendees relative to the general population in 2022 | 0.66 – 4.63 | 1.81 – 4.63 (3.42) | |
| **Household contact study data** | | | |
| Prevalence of infection in children in control households | 0.046 – 0.13 | 0.060 – 0.13 (0.10) | See Section 1.1.12 |
| Prevalence of infection in child household contacts of people diagnosed with TB who are HIV- | 0.19 – 0.31 | 0.19 – 0.31 (0.24) | |
| Relative prevalence of infection in child household contacts of people diagnosed with TB who are HIV+ART+ compared to HIV- | 0.50 – 0.99 | 0.50 – 0.79 (0.61) | |
| Relative prevalence of infection in child household contacts of people diagnosed with TB who are HIV+ART- compared to HIV- | 0.41 – 0.87 | 0.41 – 0.73 (0.52) | |
| Relative prevalence of infection in child household contacts of HIV+ART+ people diagnosed with symptom-screen negative TB | 0.35 – 2.69 | 0.76 – 0.98 (0.92) | |

| |
|---|
| compared to symptom-screen positive TB |

#### 1.2 Costing methods

We conducted primary data collection from Feb 7, 2024 to Feb 15, 2024 at six public health facilities (district hospitals, primary care clinics), including a district hospital laboratory. We observed workflows and interviewed TB nurses, district TB coordinators/managers, hospital managers, and laboratory managers to map TB service processes for cost parameterisation. We developed process maps for TB-related services at each level of the health system to enable parameterization of cost models to reflect a range of potential diagnostic algorithms. We estimated costs for all direct and ancillary services which could be included in a diagnostic algorithm; services and their cost estimates are listed in Table S6.

For each service we identified all resources used to produce the service, including capital (building, equipment and furniture, vehicles, and training) and recurrent (staff, supplies, maintenance, utilities, fuel, food and other recurrent) inputs. Resource quantities were obtained from facility and government documentation and in-person interviews. In the case of services not currently in operation at-scale in South Africa, we identified the potential resources involved using in-person interviews and data from pilot projects. Commodity resource prices were estimated using the National Health Laboratory Services (NHLS) state price list [49]. Staff salaries were estimated using the average salary across all grades per cadre (1), and using public service pay scales for non-health care workers [50]. Prices for other resources, as well as overheads and other fixed costs, were sourced from published and grey literature. We assumed a relative standard error of 20% for costs collected as primary data.

We supplemented primary data collection with comprehensive searches of the literature to identify suitable unit cost estimates for TB-related services, with a prioritization on the study having taken place within the last 5 years and the study having taken place in South Africa. We adapted recent unit cost estimates for symptom screening and sputum collection [51], lab-based Xpert MTB/RIF Ultra [52], mobile chest X-ray units, ward-based outreach teams [53], TB treatment [54], and HIV treatment [55], to our setting. Overhead costs for all health facility-based services were adapted from a study in Georgia to reflect the South African setting [56].

All costs from secondary data were classified as tradeable, non-tradeable, and staff costs. Tradeable goods included things that can easily be traded in the international market, such as supplies. To adjust these we converted the price to USD using the exchange rate for the year of study, and then inflated to 2023 USD using the US Consumer Price Index [57] [58]. Non-tradeable goods cannot be traded in international markets and are generally consumed in the country where they are produced; these include for example buildings and utilities. Non-tradeable and staff costs were adjusted by first inflating the price to 2023 Rand using the South Africa Consumer Price Index [58], and then converting to 2023 USD using the average official exchange rate for the year 2023 [59]. Where the 'reference' prices used were not from South Africa (e.g. overhead costs), we applied an adjustment factor to staff and non-tradeable costs representing the relative GDP per capita expressed in current international dollars, converted by purchasing power parities [60].

Costs were parameterized using a mechanistic cost function approach, which accounted for variations in unit costs at different scales of implementation. We disaggregated costs into fixed programme costs (including the costs of running Ward-Based Outreach Teams, trainings, and transport of lab samples), fixed facility- and lab-level costs (including building, utilities, management, and overhead costs), and variable costs (including supplies, staff time and other costs) [61]. Fixed costs assumed no variation with scale and were set according to the national number of health facilities, labs, and Ward-Based Outreach Teams. We assumed a total of 4206 public clinics, 71 public hospitals, and 162 district labs [62], and a total of 7800 Ward-Based Outreach Teams in South Africa [63]. We assumed 80% of each of these would participate in active case finding activities regardless of the number of people screened/tested. We assumed that costs of TB treatment and ART were fully variable, as we were unable to find recent estimates reported by cost ingredient. Lifetime costs of ART were estimated according to total life expectancy by age.[16]

In all scenarios, we assumed that symptom screening occurring at the health facility would take place in a private room, conducted by an enrolled nurse. Similarly, we assumed that TUTT eligibility screening was conducted by a nurse in a private space. We assumed mobile chest radiography results were interpreted by a radiographer, and assumed some community mobilization would occur in each location to recruit participants. We assumed WBOTs worked in areas within 5 kilometres of a health facility, and that all 'high prevalence' targeting would take place within these areas. For the community-level approaches, we assumed some additional training would be required for WBOTs, and assumed a small cost for cool bags where sputum samples were collected at the community level. We assumed that sputum samples for all people who could produce sputum would be sent to a district laboratory for testing with Xpert Ultra. We assumed sputum samples collected by WBOTs would not require any extra investment for delivery to the district lab, as they would likely be delivered to local health facilities by WBOTs on foot, and would then included in existing shipments from the health facility to the district lab. We assumed each mobile radiography unit would have a maximum annual throughput of 12,000 tests [64]. We assumed that the national number of mobile radiography units would scale with the proportion of the population that was targeted, in increments of five.

Table S6 Costs of TB-related services (2023 USD)

| Service | Fixed equipment costs | Fixed staff costs | Fixed overhead costs | Fixed transport costs | Variable consumables costs | Variable other costs | Variable staff costs | Total variable costs | Total fixed costs | Fixed unit definition |
|---|---|---|---|---|---|---|---|---|---|---|
| Symptom Screening - Static Sites | 2.15 | 60.43 | 253.30 | - | 0.00 | - | 0.45 | 0.46 | 315.88 | per clinic per year (n = 4206) |
| LTFU - Phone Call - Static Sites | 48.21 | 249.38 | 436.89 | - | - | 0.34 | 0.57 | 0.90 | 734.48 | per clinic per year (n = 4206) |
| Sputum collection attempt - Static Sites | 58.22 | 1,349.19 | 3,264.35 | - | 1.90 | - | 1.13 | 3.04 | 4,671.76 | per clinic per year (n = 4206) |
| Doctor Visit - Static Site | 3,652.69 | 737.54 | 1,411.40 | - | 0.07 | - | 9.24 | 9.31 | 5,801.63 | per clinic per year (n = 4206) |
| Results Notification - SMS | - | - | - | - | - | 0.37 | - | 0.37 | - | per hospital per year (n = 71) |
| Chest X-ray - Static Sites | 293.79 | 581.15 | 1,127.26 | - | 0.13 | - | 8.36 | 8.50 | 2,002.20 | per hospital per year (n = 71) |
| GXP Sample transport - District Laboratory | - | - | - | 2,993.39 | 0.26 | - | - | 0.26 | 2,993.39 | per lab per year (n = 162) |
| Full haemogram | 434.05 | 561.99 | 1,227.51 | - | 3.52 | - | 0.68 | 4.20 | 2,223.55 | per lab per year (n = 162) |
| GXP - District Laboratory | 25,788.30 | - | 2,284.49 | - | 8.31 | - | 10.37 | 18.68 | 28,072.80 | per lab per year (n = 162) |
| Chest X-ray - Mobile | 8,077.08 | 15,335.61 | 803.35 | 17,772.75 | 0.13 | - | 3.31 | 3.45 | 41,988.80 | per unit per year (n = 162) |
| Fixed costs - WBDOT pair | - | - | 242.90 | - | - | - | - | - | 242.90 | per team per year (n = 7800) |
| Symptom screen - community | - | - | - | - | 0.03 | - | 1.19 | 1.23 | - | per team per year (n = 7800) |
| Sputum collection attempt - community | 135.16 | 2,840.16 | - | - | 1.25 | - | 3.35 | 4.60 | 2,975.32 | per team per year (n = 7800) |
| ART treatment (per patient per year) | - | - | - | - | - | - | - | 241.77 | - | |
| Outpatient treatment (DS-TB) - adults | - | - | - | - | - | - | - | 250.49 | - | |
| Symptom screen - community - travel | - | - | - | - | 0.21 | - | 7.14 | 7.34 | - | |

Table S7 Costs of TB-related services (2023 ZAR)

| Service | Fixed equipment costs | Fixed staff costs | Fixed overhead costs | Fixed transport costs | Variable consumables costs | Variable other costs | Variable staff costs | Total variable costs | Total fixed costs | Fixed unit definition |
|---|---|---|---|---|---|---|---|---|---|---|
| Symptom Screening - Static Sites | 39.62 | 1,115.00 | 4,673.52 | - | 0.04 | - | 8.37 | 8.41 | 5,828.14 | per clinic per year (n = 4206) |
| LTFU - Phone Call - Static Sites | 889.44 | 4,601.11 | 8,060.75 | - | - | 6.21 | 10.46 | 16.67 | 13,551.30 | per clinic per year (n = 4206) |
| Sputum collection attempt - Static Sites | 1,074.15 | 24,892.88 | 60,228.06 | - | 35.08 | - | 20.92 | 56.00 | 86,195.09 | per clinic per year (n = 4206) |
| Doctor Visit - Static Site | 67,393.08 | 13,607.83 | 26,040.60 | - | 1.32 | - | 170.40 | 171.72 | 107,041.51 | per clinic per year (n = 4206) |
| Results Notification - SMS | - | - | - | - | - | 6.76 | - | 6.76 | - | per hospital per year (n = 71) |
| Chest X-ray - Static Sites | 5,420.57 | 10,722.32 | 20,798.25 | - | 2.43 | - | 154.32 | 156.74 | 36,941.14 | per hospital per year (n = 71) |
| GXP Sample transport - District Laboratory | - | - | - | 55,228.78 | 4.87 | - | - | 4.87 | 55,228.78 | per lab per year (n = 162) |
| Full haemogram | 8,008.39 | 10,368.77 | 22,647.82 | - | 64.92 | - | 12.48 | 77.40 | 41,024.98 | per lab per year (n = 162) |
| GXP - District Laboratory | 475,800.48 | - | 42,149.47 | - | 153.32 | - | 191.32 | 344.65 | 517,949.95 | per lab per year (n = 162) |
| Chest X-ray - Mobile | 149,024.17 | 282,945.71 | 14,822.04 | 327,911.61 | 2.43 | - | 61.14 | 63.57 | 774,703.54 | per unit per year (n = 162) |
| Fixed costs - WBDOT pair | - | - | 4,481.64 | - | - | - | - | - | 4,481.64 | per team per year (n = 7800) |
| Symptom screen - community | - | - | - | - | 0.63 | - | 21.98 | 22.61 | - | per team per year (n = 7800) |
| Sputum collection attempt - community | 2,493.75 | 52,401.71 | - | - | 23.11 | - | 61.81 | 84.92 | 54,895.46 | per team per year (n = 7800) |
| ART treatment (per patient per year) | - | - | - | - | - | - | - | - | - | |
| Outpatient treatment (DS-TB) - adults | - | - | - | - | - | - | - | - | - | |
| Symptom screen - community - travel | - | - | - | - | 3.79 | - | 131.67 | 135.46 | - | |

#### 1.3 CEA Outcomes

We used disability-adjusted life years (DALYs) as our primary outcome measure; DALYs were estimated as the sum of years of life lost (YLLs) and years of life lived with disability (YLDs), and were parameterized based on time spent in different health states in the model. We estimated YLLs using the weighted mean life expectancy at age at death by HIV status.[16,19,20] We estimated YLDs using disability weights for each health state, sourced from the most recent Global Burden of Disease study,[21] with expert-informed mapping for states not explicitly defined (e.g., false-positive TB treatment or chronic respiratory symptoms). All disability weights are specified in Table S8. We assumed that TB-related symptoms would subside after the first two months of treatment on average. Combined disability weights used a multiplicative approach.[22] YLDs were parameterized following a beta distribution in the model. All outcomes were discounted at 5% in the base case.[18]

Table S8 Disability weights used in model parameterization

| Intervention | Mean | SE | Source and Notes |
|---|---|---|---|
| Full health (no TB) | 0.00 | | |
| Active TB & HIV | 0.41 | 0.07 | [65] |
| Active TB, no HIV | 0.33 | 0.06 | [65] |
| HIV, no active TB | 0.27 | 0.05 | [65] |
| HIV on ART, no active TB | 0.08 | 0.02 | [65] |
| Post-TB | 0.03 | 0.00 | [66] |
| TB treatment due to a false-positive TB diagnosis | 0.05 | 0.01 | [65] mapped to health state "Generic uncomplicated disease: worry and daily medication" |
| HIV on ART, active TB | 0.33 | 0.06 | [65] |
| Treated TB | 0.14 | 0.01 | [65] Assumed 2 months equivalent to health state "Active TB"; 4 months equivalent to health state "Generic uncomplicated disease: worry and daily medication". |
| Chronic respiratory disease | 0.02 | 0.01 | [65] mapped to health state "COPD and other chronic respiratory problems, mild" |

### 2 Supplementary Results

#### 2.1 Model calibration

##### 2.1.1 Fit to data

We identified a total of 1041 full fitting points using history matching and model emulation. See Table S3 for the range and median values across the 1041 points of each input parameter varied during calibration, and Table S5 for the range and median values of each calibration target in the 1041 points.

##### 2.1.2 TB mortality rates in people with chronic conditions

The median rate of TB mortality for people with chronic conditions that cause TB symptoms compared to people without chronic conditions that cause TB symptoms in 2024 in the fitting parameter sets was 2.71 (95% range 2.17 – 3.43) in HIV- people, 1.82 (1.48 – 2.23) in HIV+ people who are not ART, and 2.29 (1.82 – 2.88) in HIV+ people who are on ART. These ratios are in line with the plausible ratio we elicited of 1.7 – 3.0 (see Section 1.1.7.2).

#### 2.2 Additional results figures

Figure S5 Number of Xperts/testing coverage as part of active case finding in 2026, by HIV/ART status and scenario

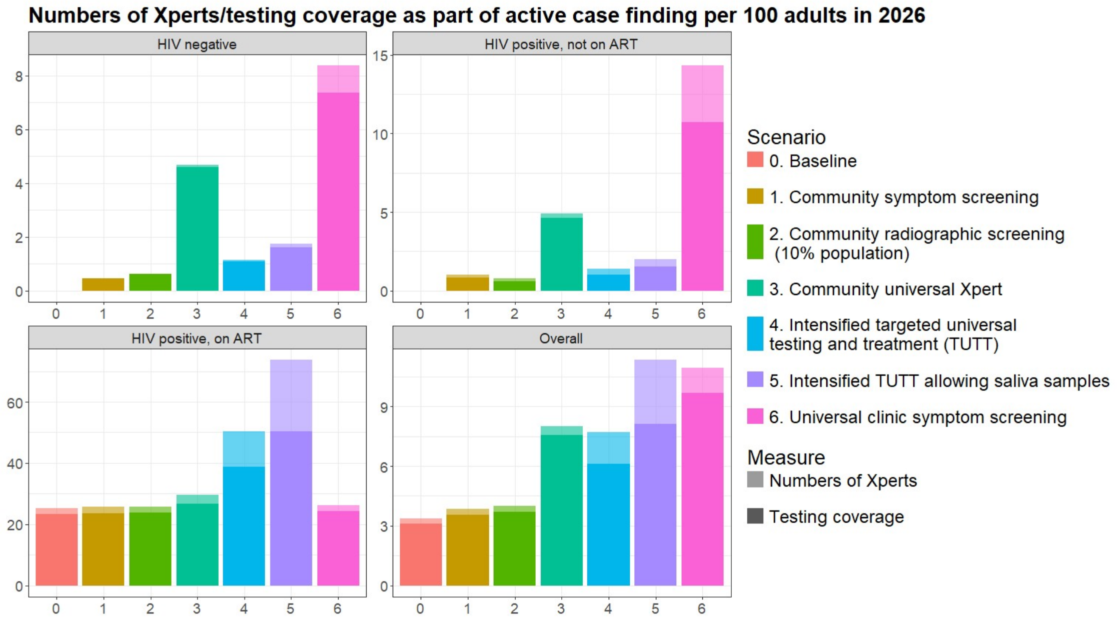

The full height of the bars shows the number of Xperts conducted. The height of the darker bars shows the proportion of the population who received a test. The difference in height between the two bars indicates that

some individuals received more than one Xpert as part of active case finding in 2026. The height of the bars are zero for HIV negative people and HIV positive people who are not on ART in the baseline scenario, as we assume that active case finding is only being conducted for people on ART in the baseline scenario

Figure S6 Notification rate of true positive TB in 2026, by HIV/ART status and scenario

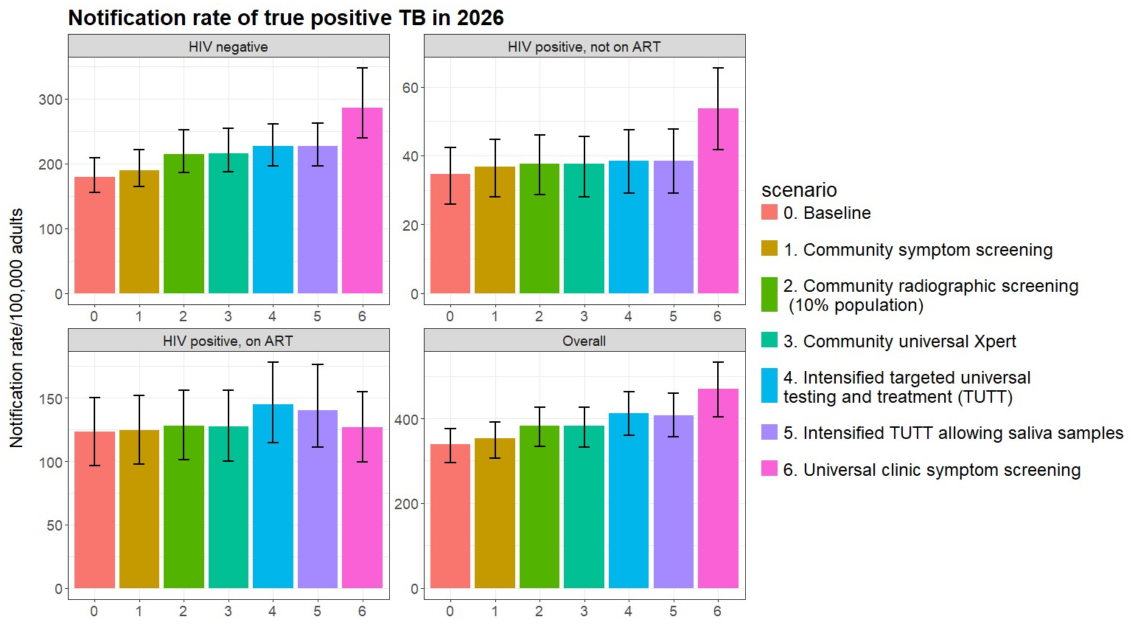

Figure S7 Mean TB episode duration in 2026, by HIV/ART status and scenario

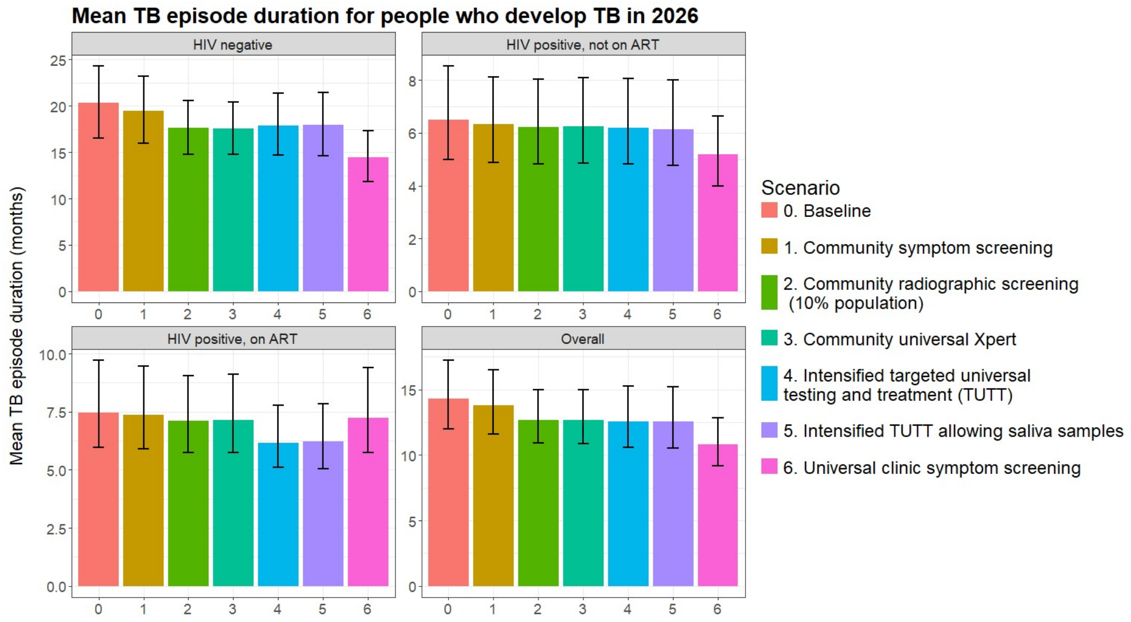

Figure S8 Yield of true positive TB per Xpert test conducted as part of active case finding in 2026, by HIV/ART status and scenario.

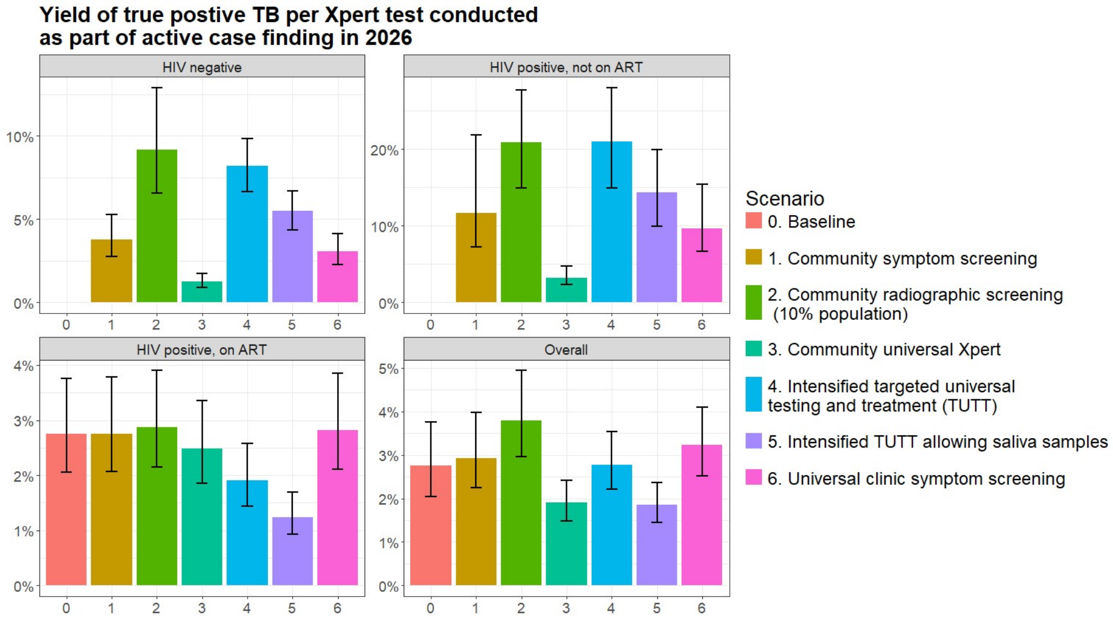

#### 2.3 Partial rank correlation coefficients

Figure S9 Partial rank correlation coefficients plots, for each intervention approach

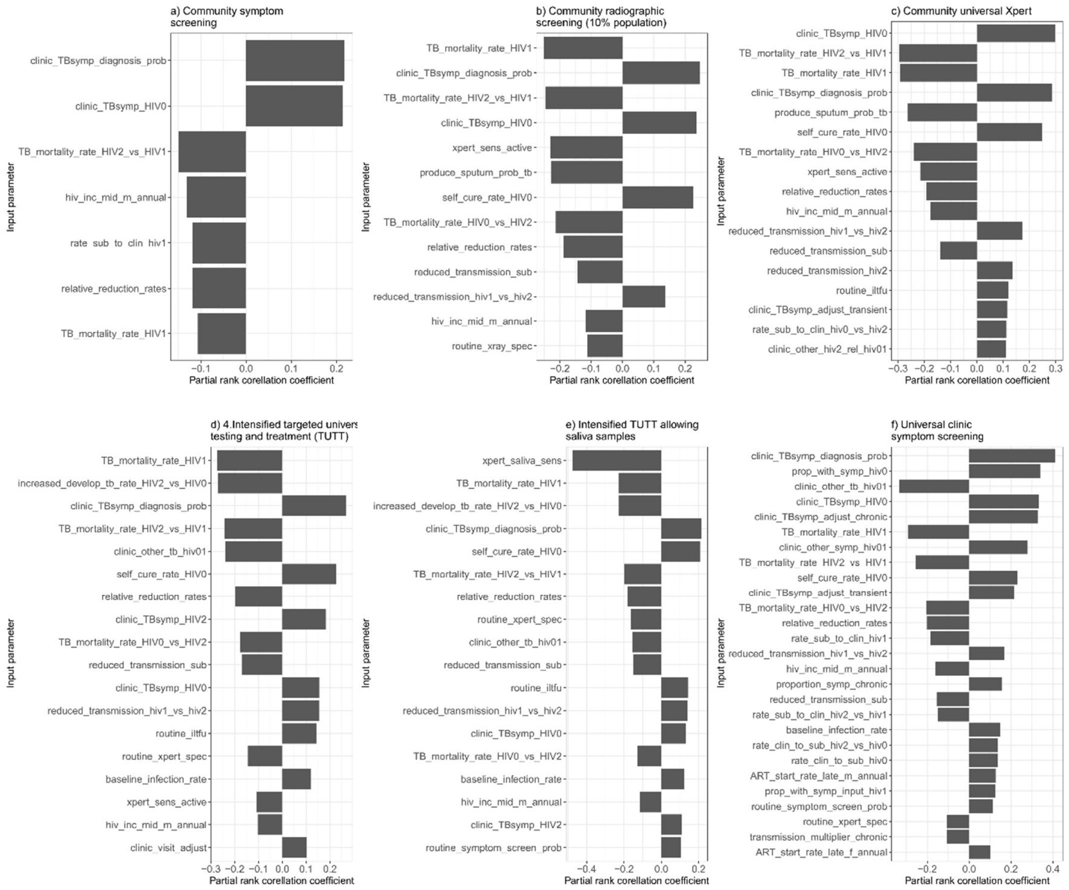

The plots show the strength of the relationship between the values of the input parameters that were varied in the mathematical model runs and the incremental cost effectiveness ratios. Input parameters where the absolute value of the point estimate of the partial rank correlation coefficient was less than 0.1 are not shown.

#### 2.4 Additional cost-effectiveness results

Figure S10 Mean incremental costs and DALYs averted

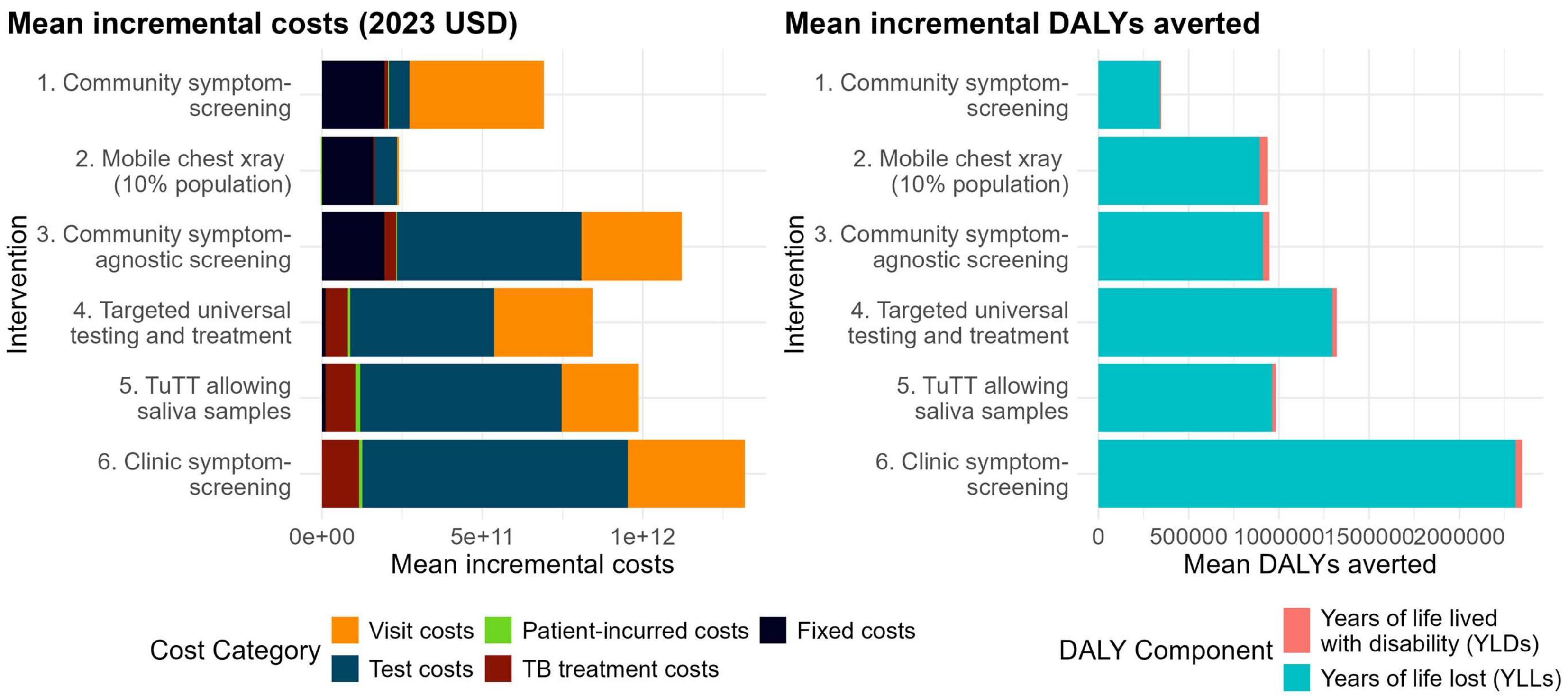

Table S 9 Mean total and incremental costs and DALYs (South African Rand)

| Variable | Baseline | 1. Community symptom screening | 2. Community radiographic screening | 3. Community universal Xpert | 4. Intensified TUTT | 5. Intensified TUTT allowing saliva samples | 6. Clinic symptom screening |
|---|---|---|---|---|---|---|---|
| Mean annual incidence of infectious TB in 2026-2035 | 438 | 423 | 391 | 389 | 376 | 385 | 328 |
| Mean reduction in the incidence of infectious TB in 2026-3035, relative to baseline | | 3.4% | 10.7% | 11.2% | 14.2% | 12.1% | 25.1% |
| Total cost (2023 ZAR) | R299.7B | R311.2B | R306.7B | R319.8B | R315.2B | R317.5B | R324.6B |
| *Total provider- incurred cost* | R267.7B | R279.2B | R274.8B | R287.8B | R283.2B | R285.4B | R292.4B |
| *Total patient- incurred cost* | R32.0B | R32.0B | R31.9B | R32.0B | R32.1B | R32.1B | R32.2B |
| Total DALYs | 137.6M | 137.3M | 136.7M | 136.7M | 136.3M | 136.3M | 135.3M |
| *Years of life lost* | 152.3M | 151.9M | 151.1M | 151.1M | 150.6M | 150.7M | 149.3M |
| *Years of life lived with disability* | 19.1M | 19.1M | 19.1M | 19.1M | 19.1M | 19.1M | 19.1M |
| Mean incremental cost (2023 ZAR) | | R11.5B | R7.1B | R20.1B | R15.5B | R17.8B | R24.9B |
| Mean incremental DALYs averted | | 0.3M | 0.9M | 0.9M | 1.3M | 1.3M | 2.4M |
| ICER (2023 ZAR) | | R35,055 | R7,708 | R21,851 | R12,102 | R14,375 | R10,863 |

Table S10 Base intervention approaches proportion cost-effective by threshold

| Approach | Mean ICER | 95% Confidence Interval | Proportion of runs where the approach is cost-effective WTP = $618 / DALY averted | WTP = $985 / DALY averted | WTP = $3,1279 / DALY averted |
|---|---|---|---|---|---|
| 1. Community symptom screening | $1,903 | $1,134 - $3,389 | 0% | 0% | 96% |
| 2. Community radiographic screening | $418 | $287 - $613 | 98% | 100% | 100% |
| 3. Community universal Xpert | $1,182 | $829 - $1,779 | 0% | 20% | 100% |
| 4. Intensified TUTT | $655 | $428 - $994 | 45% | 97% | 100% |
| 5. Intensified TUTT allowing saliva samples | $1,003 | $602 - $1,601 | 3% | 52% | 100% |
| 6. Clinic symptom screening | $586 | $388 - $870 | 65% | 99% | 100% |

Table S11 Full expansion path analysis results

| Approach | Mean total costs | Mean total DALYs | Mean incremental costs | Mean incremental DALYs averted | Mean ICER |
|---|---|---|---|---|---|
| 0. Baseline | 16,243,176,136 | 137,628,110 | - | - | - |
| 1. Community symptom screening | 16,865,500,413 | 137,279,860 | 622,324,278 | 348,250 | 1,899.98 |
| 2. Community radiographic screening | 16,625,578,806 | 136,687,806 | 382,402,671 | 940,304 | 417.75 |
| 3. Community universal Xpert | 17,333,952,359 | 136,680,056 | 1,090,776,223 | 948,054 | 1,184.33 |
| 4. Intensified targeted universal testing and treatment (TUTT) | 17,084,446,546 | 136,304,837 | 841,270,410 | 1,323,273 | 655.92 |
| 5. Intensified TUTT allowing saliva samples | 17,208,981,554 | 136,341,852 | 965,805,418 | 1,286,258 | 779.13 |
| 6. Universal clinic symptom screening | 17,594,105,352 | 135,275,995 | 1,350,929,216 | 2,352,115 | 588.78 |
| 7. Community radiographic screening (20% population) | 16,999,538,699 | 136,129,165 | 756,362,563 | 1,498,945 | 516.47 |
| 8. Community radiographic screening (40% population) | 17,741,359,118 | 135,393,593 | 1,498,182,983 | 2,234,518 | 685.26 |

| | | | | | |
|---|---|---|---|---|---|
| 9. Community radiographic screening (60% population) | 18,479,312,567 | 134,914,034 | 2,236,136,431 | 2,714,076 | 841.27 |
| 10. Community radiographic screening (80% population) | 19,215,796,057 | 134,570,079 | 2,972,619,921 | 3,058,031 | 992.02 |
| 12. Universal clinic symptom screening, differentiated frequencies | 17,724,267,665 | 135,113,269 | 1,481,091,529 | 2,514,841 | 603.39 |
| 13. Universal clinic symptom screening, every 2 months | 18,017,252,489 | 134,994,481 | 1,774,076,353 | 2,633,629 | 690.55 |
| 14. TuTT, differentiated frequencies | 17,257,252,308 | 135,891,947 | 1,014,076,172 | 1,736,163 | 603.58 |
| 15. TuTT, every 2 months | 17,611,127,483 | 135,786,882 | 1,367,951,348 | 1,841,229 | 767.23 |
| 16. TuTT with saliva, differentiated frequency | 17,742,644,015 | 135,820,366 | 1,499,467,879 | 1,807,744 | 858.77 |
| 17. TuTTs, every 2 months | 18,146,794,667 | 135,665,769 | 1,903,618,531 | 1,962,341 | 1,003.64 |
| 18. Community universal Xpert + Universal clinic symptom screening | 18,190,120,043 | 134,780,644 | 1,946,943,908 | 2,847,466 | 698.03 |
| 19. Community Community universal Xpert 10% + Intensified TUTT | 17,374,145,422 | 135,530,219 | 1,130,969,287 | 2,097,891 | 551.20 |
| 20. Community universal Xpert 10% + Intensified TUTT with saliva | 17,499,641,866 | 135,563,822 | 1,256,465,731 | 2,064,288 | 623.08 |
| 21. Community universal Xpert 20% + Universal clinic symptom screening | 18,786,265,698 | 134,486,404 | 2,543,089,562 | 3,141,706 | 825.52 |
| 22. Community universal Xpert 20% + Intensified TUTT | 17,981,989,953 | 135,081,714 | 1,738,813,817 | 2,546,396 | 697.43 |
| 23. Community universal Xpert 20% + Intensified TUTT with saliva | 18,111,044,144 | 135,115,255 | 1,867,868,009 | 2,512,855 | 759.72 |
| 24. Community symptom screening 10% + Universal clinic symptom screening | 17,722,386,944 | 135,104,766 | 1,479,210,809 | 2,523,344 | 600.32 |
| 25. Community symptom screening 10% + Intensified TUTT | 16,902,699,649 | 136,012,940 | 659,523,514 | 1,615,170 | 419.06 |
| 26. Community symptom screening 10% + Intensified TUTT with saliva | 17,025,146,881 | 136,053,562 | 781,970,745 | 1,574,548 | 511.13 |
| 27. Community symptom screening 20% + Universal clinic symptom screening | 17,849,651,120 | 134,990,439 | 1,606,474,984 | 2,637,671 | 623.19 |
| 28. Community symptom screening 20% + Intensified TUTT | 17,036,752,942 | 135,818,470 | 793,576,806 | 1,809,640 | 449.25 |
| 29. Community symptomscreening 20% + Intensified TUTT with saliva | 17,159,835,355 | 135,863,776 | 916,659,219 | 1,764,334 | 533.76 |
| 30. Community radiographic screening 10% + Universal clinic symptom screening | 17,967,494,698 | 134,779,728 | 1,724,318,562 | 2,848,382 | 617.87 |
| 31. Community radiographic screening 10% + Intensified TUTT | 17,141,652,398 | 135,528,398 | 898,476,263 | 2,099,712 | 437.28 |

| | | | | | |
|---|---|---|---|---|---|
| 32. Community radiographic screening 10% + Intensified TUTT with saliva | 17,265,204,395 | 135,565,462 | 1,022,028,259 | 2,062,648 | 506.90 |
| 33. Community radiographic screening 20% + Universal clinic symptom screening | 18,336,557,786 | 134,478,926 | 2,093,381,651 | 3,149,184 | 677.48 |
| 34. Community radiographic screening 20% + Intensified TUTT | 17,512,251,696 | 135,081,445 | 1,269,075,560 | 2,546,665 | 508.56 |
| 35. Community radiographic screening 20% + Intensified TUTT with saliva | 17,636,930,366 | 135,119,888 | 1,393,754,230 | 2,508,222 | 567.25 |
| 36. Community radiographic screening 40% + Universal clinic symptom screening | 19,072,624,152 | 134,074,340 | 2,829,448,016 | 3,553,770 | 810.67 |
| 37. Community radiographic screening 40% + Intensified TUTT | 18,250,489,601 | 134,499,899 | 2,007,313,466 | 3,128,211 | 654.20 |
| 38. Community radiographic screening 60% + Universal clinic symptom screening | 19,807,684,814 | 133,798,831 | 3,564,508,679 | 3,829,279 | 947.29 |
| 39. Community radiographic screening 60% + Intensified TUTT | 18,987,178,879 | 134,124,956 | 2,744,002,744 | 3,503,154 | 798.05 |
| 40. Community radiographic screening 80% + Universal clinic symptom screening | 20,542,012,364 | 133,601,116 | 4,298,836,229 | 4,026,994 | 1,086.23 |
| 41. Community radiographic screening 80% + Intensified TUTT | 19,722,220,165 | 133,859,267 | 3,479,044,029 | 3,768,843 | 940.23 |
| 44. Intensified TUTT + Universal clinic symptom screening, every 2 months | 18,639,206,884 | 134,497,055 | 2,396,030,748 | 3,131,055 | 782.18 |
| 45. Intensified TUTT + Universal clinic symptom screening, every 6 months | 18,051,514,691 | 134,878,931 | 1,808,338,555 | 2,749,179 | 672.43 |

Figure S11 Cost-effectiveness acceptability curves for intervention approaches on the expansion pathway

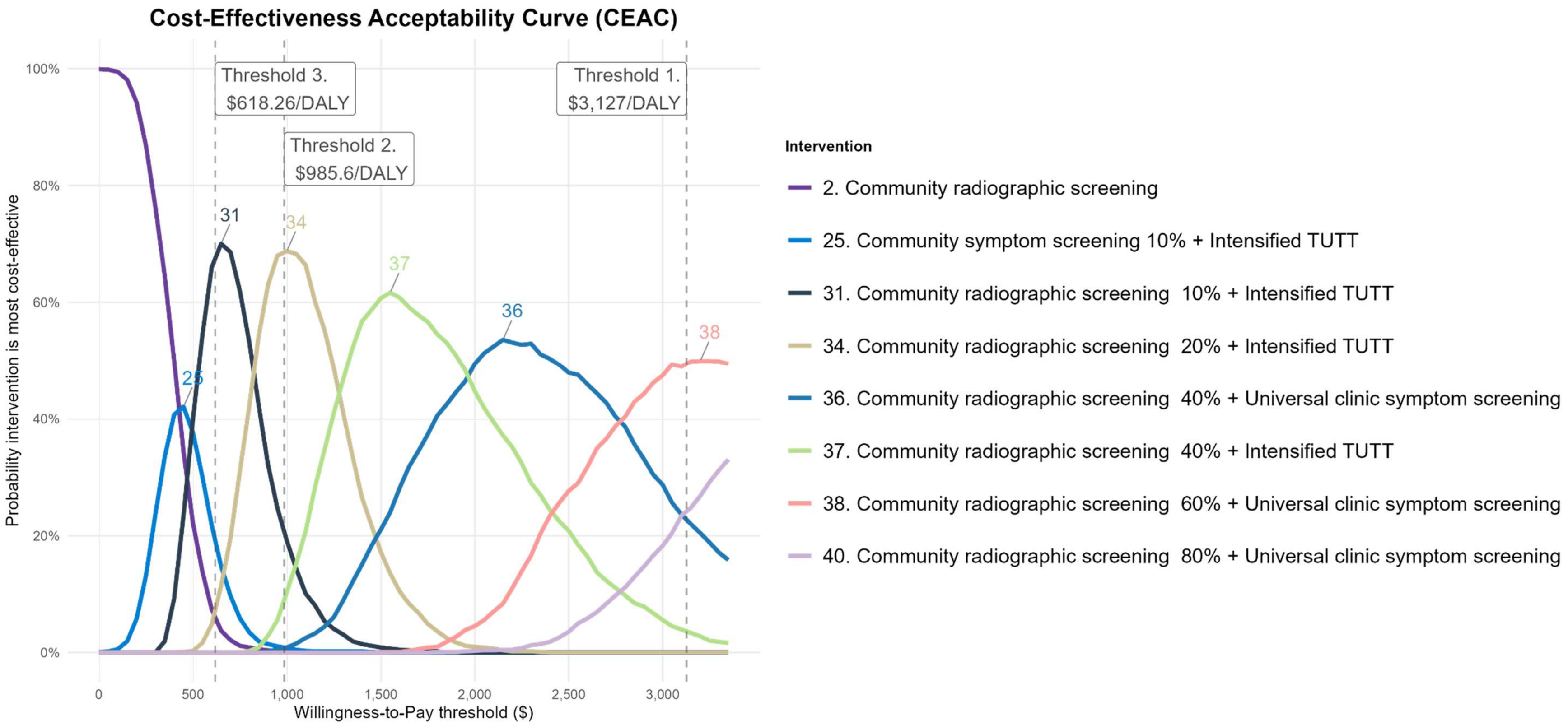

Figure S 12 One-way scenario sensitivity analysis

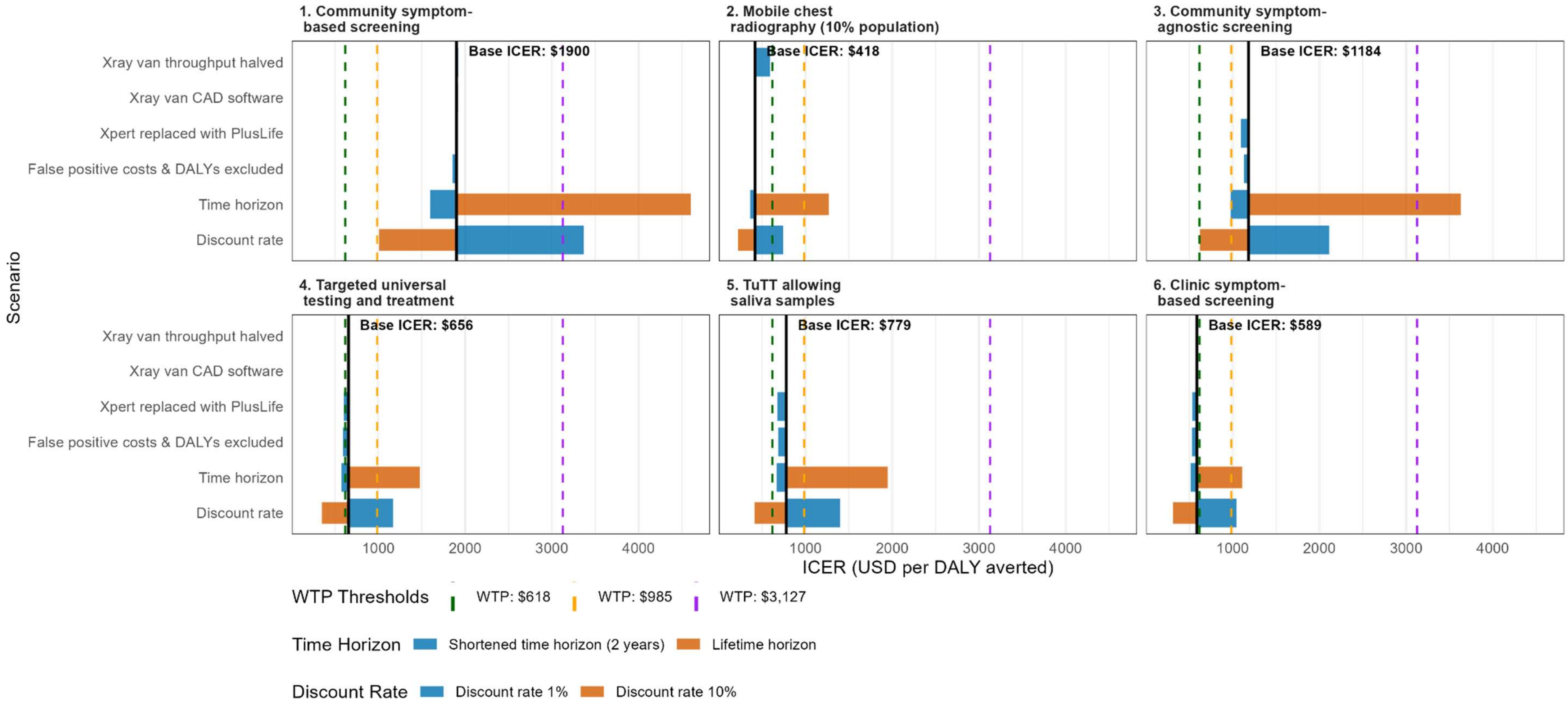

Figure S13 One-way sensitivity analysis expansion pathways

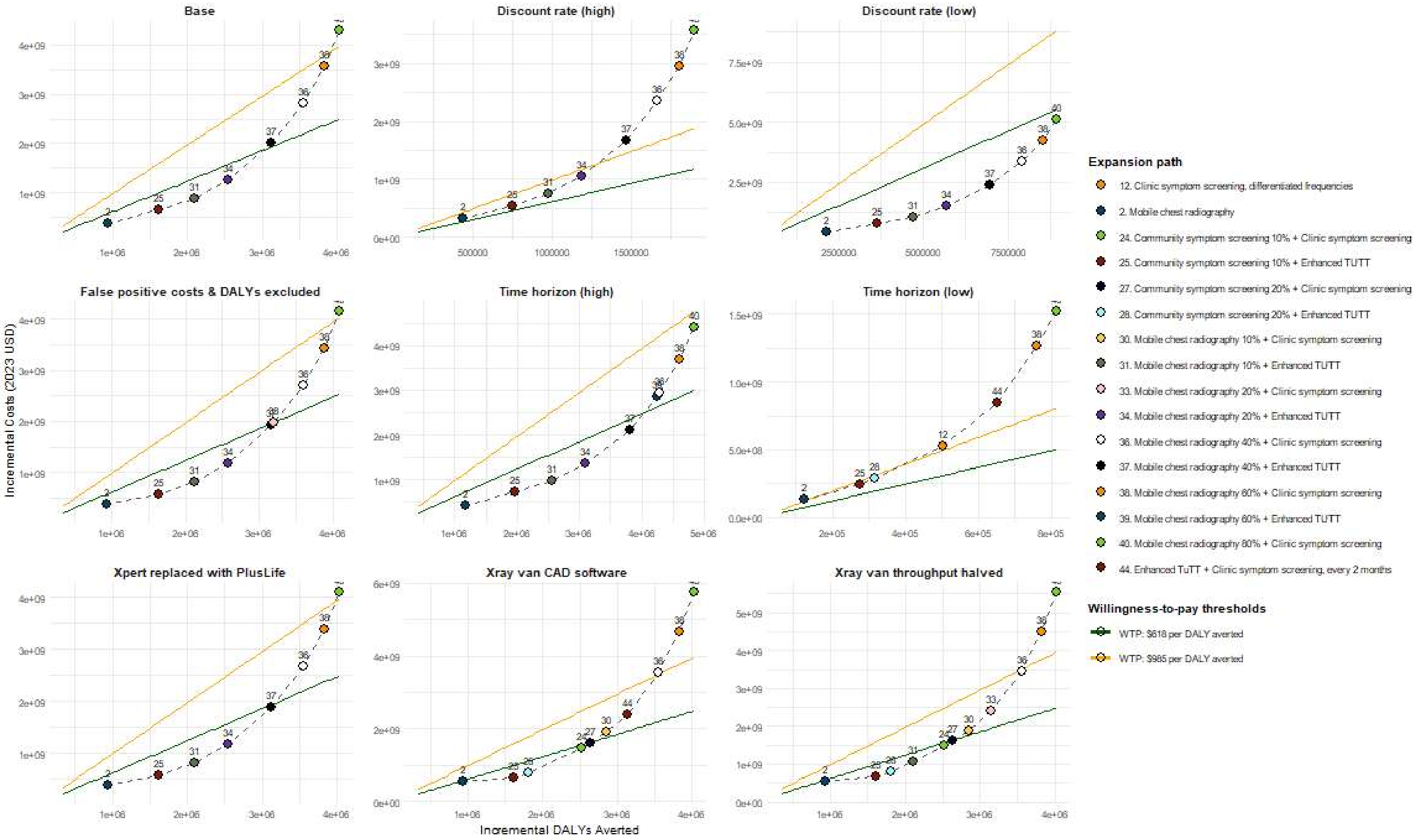

Figure S 14 Community radiographic screening throughput analysis

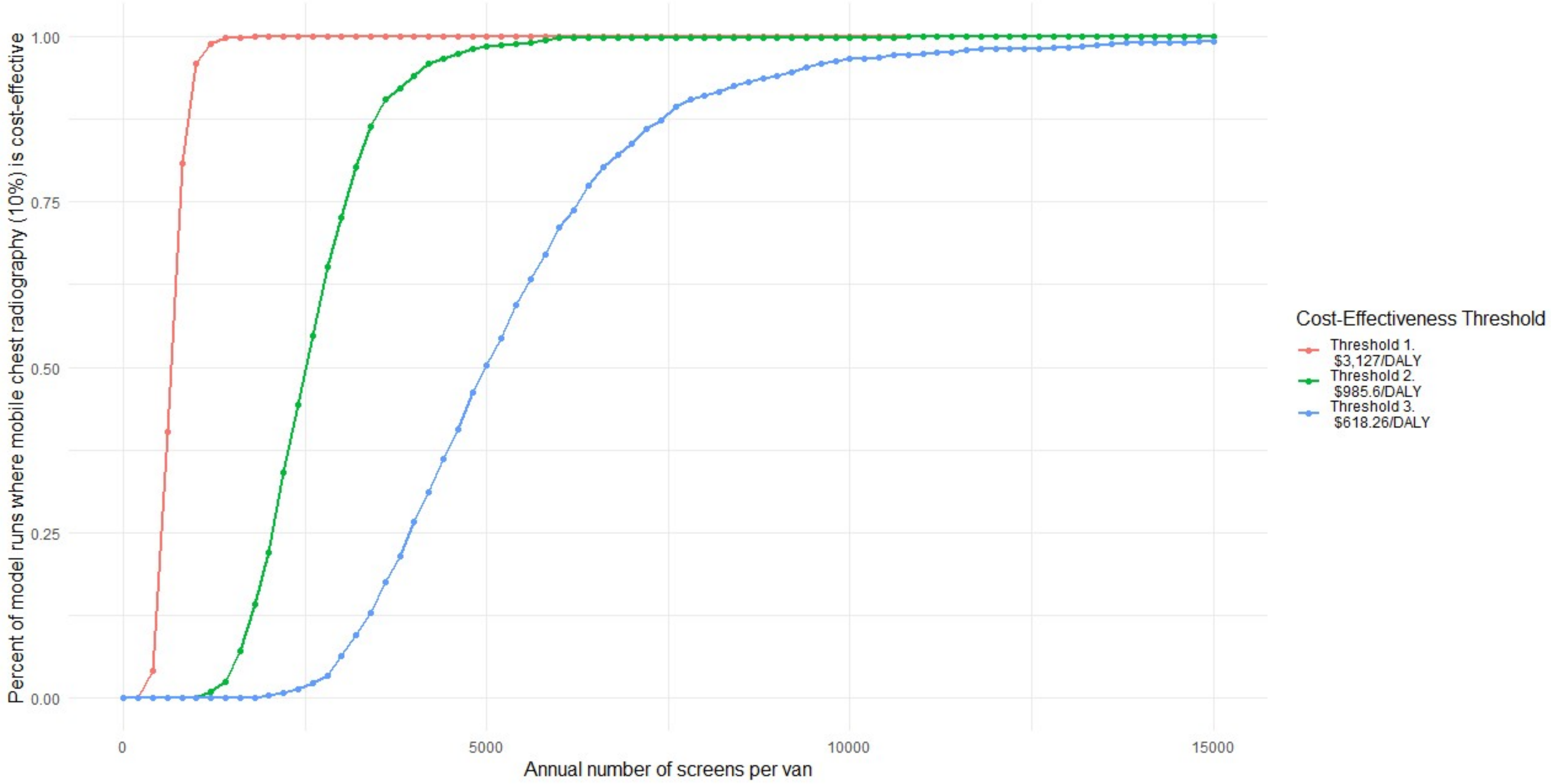

### 3 Additional discussion

We assumed that chest X-ray and Xpert have the same sensitivity in people with subclinical and clinical TB (with Xpert sensitivity higher in people who are passively seeking care in clinics only), and that the ability to produce sputum does not vary with reported symptoms. In practice, sensitivity and the ability to produce sputum are likely to be higher in people with clinical TB. The magnitude of the differences may be small however, due to the lack of clear distinction between people who do or do not report symptoms on a symptom screen[67]. In contrast, there may be more substantial differences in chest X-ray and Xpert sensitivity between people with less and more advanced TB, however it is unclear how such states should be defined or parameterised in models. In addition, the sensitivity and specificity of individual tests are likely to vary when they are used following a screening test[68], something not included in the model due to the lack of data to inform parameterisation. Finally, we used a wide plausible range for the specificity of Xpert Ultra, informed by the estimates of two systematic reviews[27,28], which may underestimate the true specificity of Xpert in community settings[69,70]. Our findings are likely to be robust to this limitation however, as our sensitivity analysis shows that excluding costs and DALYS associated with false positive diagnoses has little impact on the estimated ICERs for any approach (**Error! Reference source not found.**).

The model was calibrated to the prevalence of TB as estimated by the South Africa TB prevalence survey. As the survey only tested people who were screen-positive (on symptom screen and/or chest X-ray), and as a single Xpert and single culture have imperfect sensitivity for bacteriologically positive TB, we are likely to be underestimating the true prevalence of TB in South Africa in the model, and the true cost-effectiveness of the approaches.

The model was also calibrated to WHO estimates of TB incidence and mortality. The 2023 WHO TB report clarified that, in countries that have conducted national TB prevalence surveys, TB incidence estimates are based on "*on an assumed natural history in which most people with asymptomatic TB ultimately develop symptoms*". Incidence estimates for 2022 and 2023 are approximately equal to the sum of notifications and estimated TB deaths – suggesting that the assumed natural history may assume not only that most people who develop TB develop symptoms, but that they go on to either receive treatment or die of TB. We know that in practice TB can resolve without treatment however ('self-cure'), and allow self-cure to occur in the model. This means that, in theory, we could be underestimating TB incidence in the model. In practice, the confidence intervals around WHO incidence estimates are wide, and no fitting points were found near the upper limits of the ranges, and so it is unlikely that increasing the upper limits of the ranges would have had any effect on the results.

We did not model self-clearance of infection, assuming instead that, once infected, people retain a small lifelong risk of reactivation. This means that we may be underestimating the impact and cost-effectiveness of the approaches[71]. The magnitude of this effect will depend on the rate at which risk of reactivation decreases with time since infection, and will be higher for approaches that result in greater reductions in TB incidence. The proportion of incident TB in the model that results from an infection occurring over 10 years ago was 8.0% (95% CI 6.8 – 9.7%) in 2025, 8.4% (95% CI 6.9 – 11.3%) in 2035 in the baseline scenario, and 16.0% (95% CI 12.5 – 22.8%) in 2035 in the clinic symptom screening scenario.

Screening coverage by sex was explicitly accounted for in the model in the clinic-based approaches, as rates of clinic visiting were parameterised by sex. In the community approaches, we assumed that the coverage of screening would be the same in men as for women. As men have a higher prevalence of TB than women in South Africa[3], and coverage of screening tends to be higher in women in practice[8], we may have overestimated the impact of the community screening approaches. This is particularly the case for the universal Xpert and symptom-based community screening approaches, which we assumed would be delivered door-to-door. The community radiographic screening approach would rely on people visiting a screening van, and could therefore be targeted at locations that men tend to visit.

Community case finding is likely to be targeted at areas of the country where it is thought that the prevalence of TB is above that of the country as a whole. We parameterised the relationship between intervention coverage and prevalence in people screened using data on TB notification rates by district in South Africa. Prevalence does not correspond perfectly with notification rates however, potentially leading us to overestimate how successful an approach would be at targeting the highest prevalence areas. On the other hand, targeting could be more successful than we simulate, as it is likely that it would be done at a finer resolution than district level, and force of infection may be higher from people in high prevalence areas than low prevalence areas (e.g. due to more crowded housing). We may therefore be under- or over-estimating how the impact and cost-effectiveness of the community approaches scales with their level of coverage.

We assumed in the community radiographic screening approach that people who were able to produce sputum would only be treated if they tested positive on Xpert Ultra. If some empirical treatment occurred of people with negative Xpert results by signs of TB on chest X-ray, then we may be underestimating the impact of community radiographic screening.

We assumed that, in routine implementation of active case finding, 44.1 – 50.4% of people without TB and 48.4 – 79.9% of people with TB would be able to produce a sputum sample for testing. These are pragmatic estimates, and many studies have achieved higher proportions (e.g. the Vukuzazi community health survey[8]). If a higher proportion of people with TB produced a sputum sample in practice, then the reductions in incidence and DALYs averted would be higher.

### 4 CHEERS 2022 Checklist

| | Item | Guidance for Reporting | Reported in section |
|---|---|---|---|
| **TITLE** | | | |
| Title | 1 | Identify the study as an economic evaluation and specify the interventions being compared. | Title |
| **ABSTRACT** | | | |
| Abstract | 2 | Provide a structured summary that highlights context, key methods, results and alternative analyses. | Abstract |
| **INTRODUCTION** | | | |
| Background and objectives | 3 | Give the context for the study, the study question and its practical relevance for decision making in policy or practice. | Introduction |
| **METHODS** | | | |
| Health economic analysis plan | 4 | Indicate whether a health economic analysis plan was developed and where available. | Methods, subheading: Cost-effectiveness and expansion paths |
| Study population | 5 | Describe characteristics of the study population (such as age range, demographics, socioeconomic, or clinical characteristics). | Methods, subheading: Mathematical model |
| Setting and location | 6 | Provide relevant contextual information that may influence findings. | Methods, subheading: Mathematical model |
| Comparators | 7 | Describe the interventions or strategies being compared and why chosen. | Method, subheading: Baseline and intervention scenarios |
| Perspective | 8 | State the perspective(s) adopted by the study and why chosen. | Methods, subheading: Cost-effectiveness and expansion paths |
| Time horizon | 9 | State the time horizon for the study and why appropriate. | Methods, subheading: Baseline and intervention scenarios |
| Discount rate | 10 | Report the discount rate(s) and reason chosen. | Methods, subheadings: Costing Approach; Outcomes |
| Selection of outcomes | 11 | Describe what outcomes were used as the measure(s) of benefit(s) and harm(s). | Methods, subheading: Outcomes |
| Measurement of outcomes | 12 | Describe how outcomes used to capture benefit(s) and harm(s) were measured. | Methods, subheading: Outcomes |

| | | | |
|---|---|---|---|
| Valuation of outcomes | 13 | Describe the population and methods used to measure and value outcomes. | Methods, subheading: Outcomes |
| Measurement and valuation of resources and costs | 14 | Describe how costs were valued. | Methods, subheading: Costing Approach |
| Currency, price date, and conversion | 15 | Report the dates of the estimated resource quantities and unit costs, plus the currency and year of conversion. | Methods, subheading: Costing Approach. Supplement, Section 1.2 |
| Rationale and description of model | 16 | If modelling is used, describe in detail and why used. Report if the model is publicly available and where it can be accessed. | Methods, subheading: Mathematical model. Supplement, section 1.1 |
| Analytics and assumptions | 17 | Describe any methods for analysing or statistically transforming data, any extrapolation methods, and approaches for validating any model used. | Supplement, Section 1.1.13.2.3 |
| Characterizing heterogeneity | 18 | Describe any methods used for estimating how the results of the study vary for sub-groups. | Methods, subheading: Mathematical model. |
| Characterizing distributional effects | 19 | Describe how impacts are distributed across different individuals or adjustments made to reflect priority populations. | Not done |
| Characterizing uncertainty | 20 | Describe methods to characterize any sources of uncertainty in the analysis. | Methods, subheading: Sensitivity Analysis |
| Approach to engagement with patients and others affected by the study | 21 | Describe any approaches to engage patients or service recipients, the general public, communities, or stakeholders (e.g., clinicians or payers) in the design of the study. | Methods, subheading: Cost-effectiveness and expansion paths |
| **RESULTS** | | | |
| Study parameters | 22 | Report all analytic inputs (e.g., values, ranges, references) including uncertainty or distributional assumptions. | Supplement |
| Summary of main results | 23 | Report the mean values for the main categories of costs and outcomes of interest and summarise them in the most appropriate overall measure. | Results |
| Effect of uncertainty | 24 | Describe how uncertainty about analytic judgments, inputs, or projections affect findings. Report the effect of choice of discount rate and time horizon, if applicable. | Results |
| Effect of engagement with patients and others affected by the study | 25 | Report on any difference patient/service recipient, general public, community, or stakeholder involvement made to the approach or findings of the study | Methods, subheading: Cost-effectiveness and expansion paths |
| **DISCUSSION** | | | |
| Study findings, limitations, generalizability, and current knowledge | 26 | Report key findings, limitations, ethical or equity considerations not captured, and how these could impact patients, policy, or practice. | Discussion |
| **OTHER RELEVANT INFORMATION** | | | |

| | | | |
|---|---|---|---|
| Source of funding | 27 | Describe how the study was funded and any role of the funder in the identification, design, conduct, and reporting of the analysis | Methods, subheading: Role of the funding source |
| Conflicts of interest | 28 | Report authors conflicts of interest according to journal or International Committee of Medical Journal Editors requirements. | Author information Subheading: Conflict of interest statement |
