## Supporting info 2 for "Projected population level impact and cost-effectiveness of clinic and community-based tuberculosis screening approaches"

#### Supporting information 2

#### Baseline and intervention approach flowcharts

### Baseline

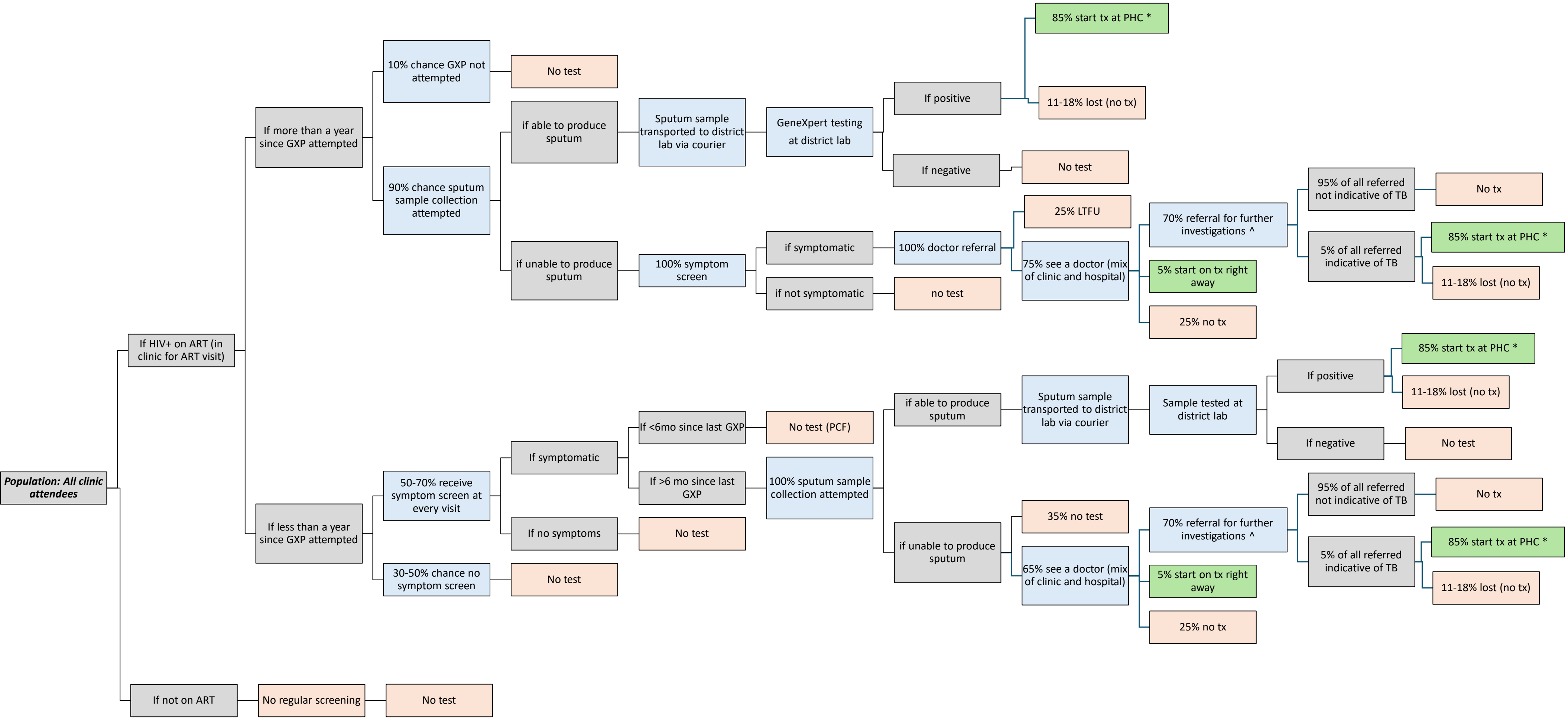


* Linkage to care activities: 50% of people <u>testing positive</u> called by nurse, 40% of people <u>receiving GXP</u> notified of result via automated text message by lab
^ Further investigations: 100% chest xray, 50% sputum attempt, 25% GeneXpert, 10% blood test FBC

### 1. Community symptom screening

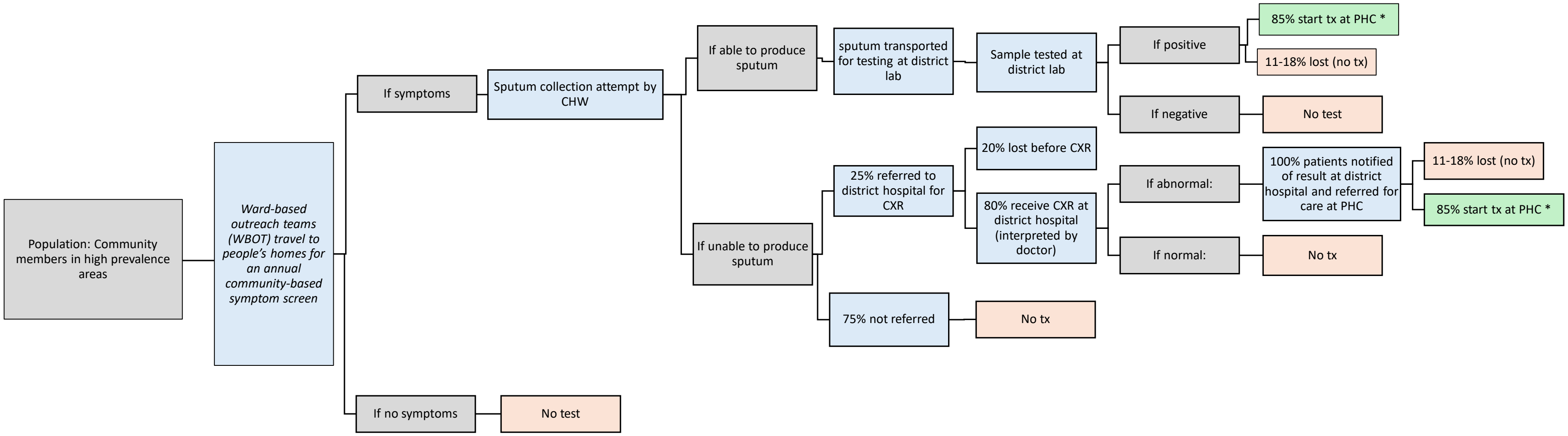


* Linkage to care activities: 50% of people testing positive called by nurse, 40% of people receiving GXP notified of result via automated text message by lab

^ Further investigations: 100% chest xray, 50% sputum attempt, 25% GeneXpert, 10% blood test FBC

### 2. Community radiographic screening

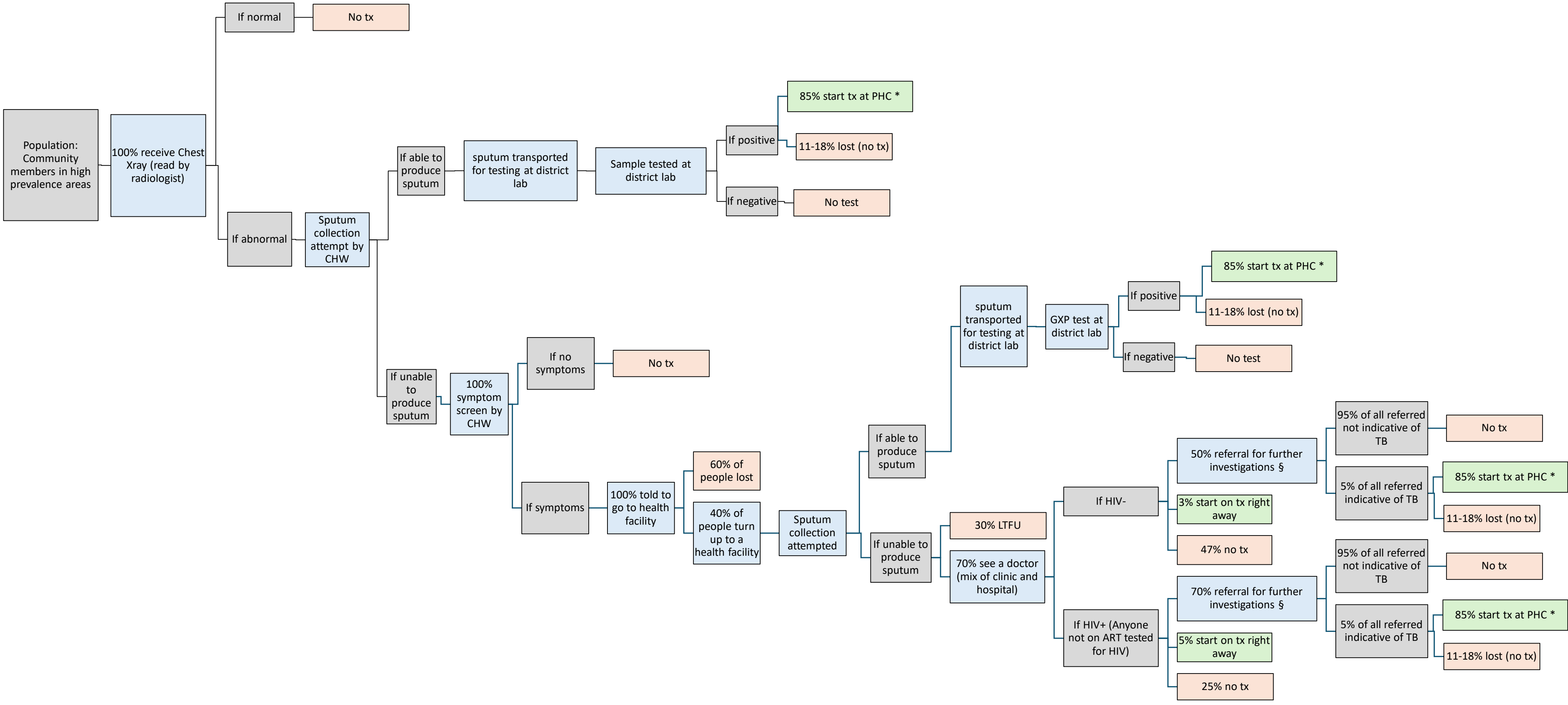


* Linkage to care activities: 50% of people testing positive called by nurse, 40% of people receiving GXP notified of result via automated text message by lab

§ Further investigations: 50% sputum attempt, 25% GeneXpert, 10% blood test FBC

### 3. Community universal Xpert

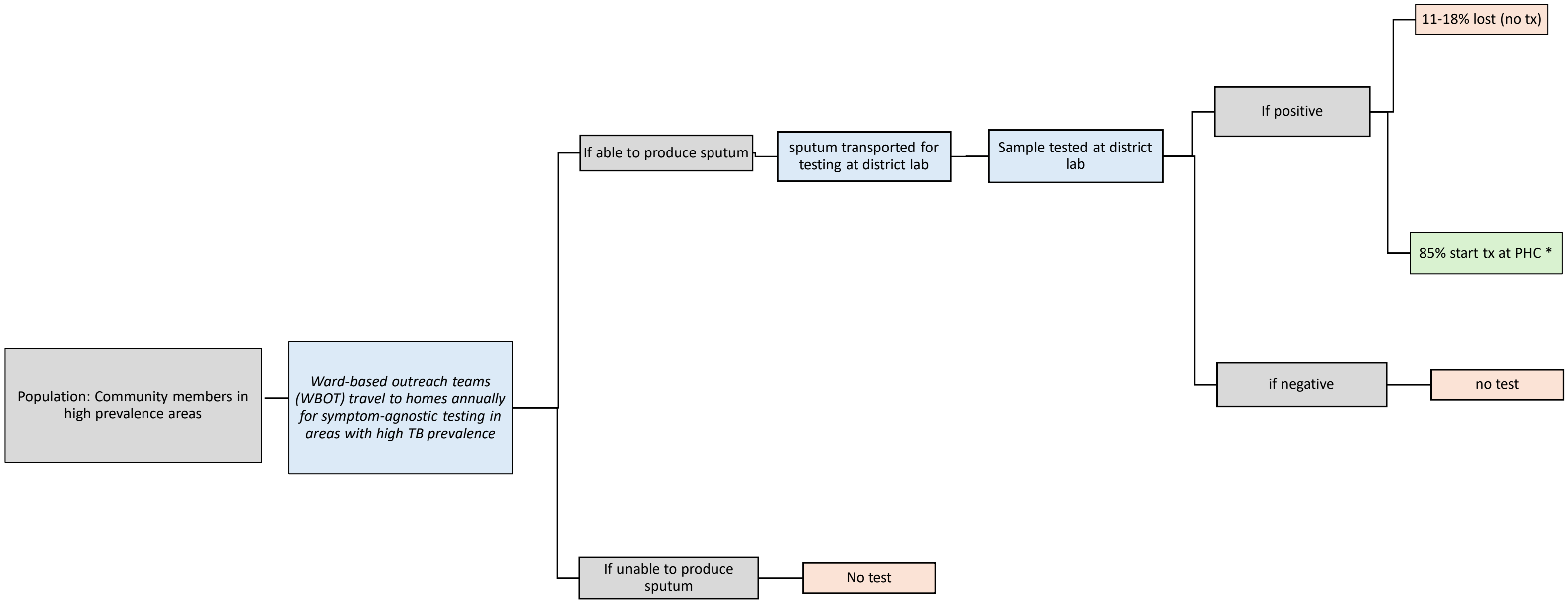


* Linkage to care activities: 50% of people testing positive called by nurse, 40% of people receiving GXP notified of result via automated text message by lab
^ Further investigations: 100% chest xray, 50% sputum attempt, 25% GeneXpert, 10% blood test FBC

### 4. Intensified targeted universal testing and treatment (TUTT)

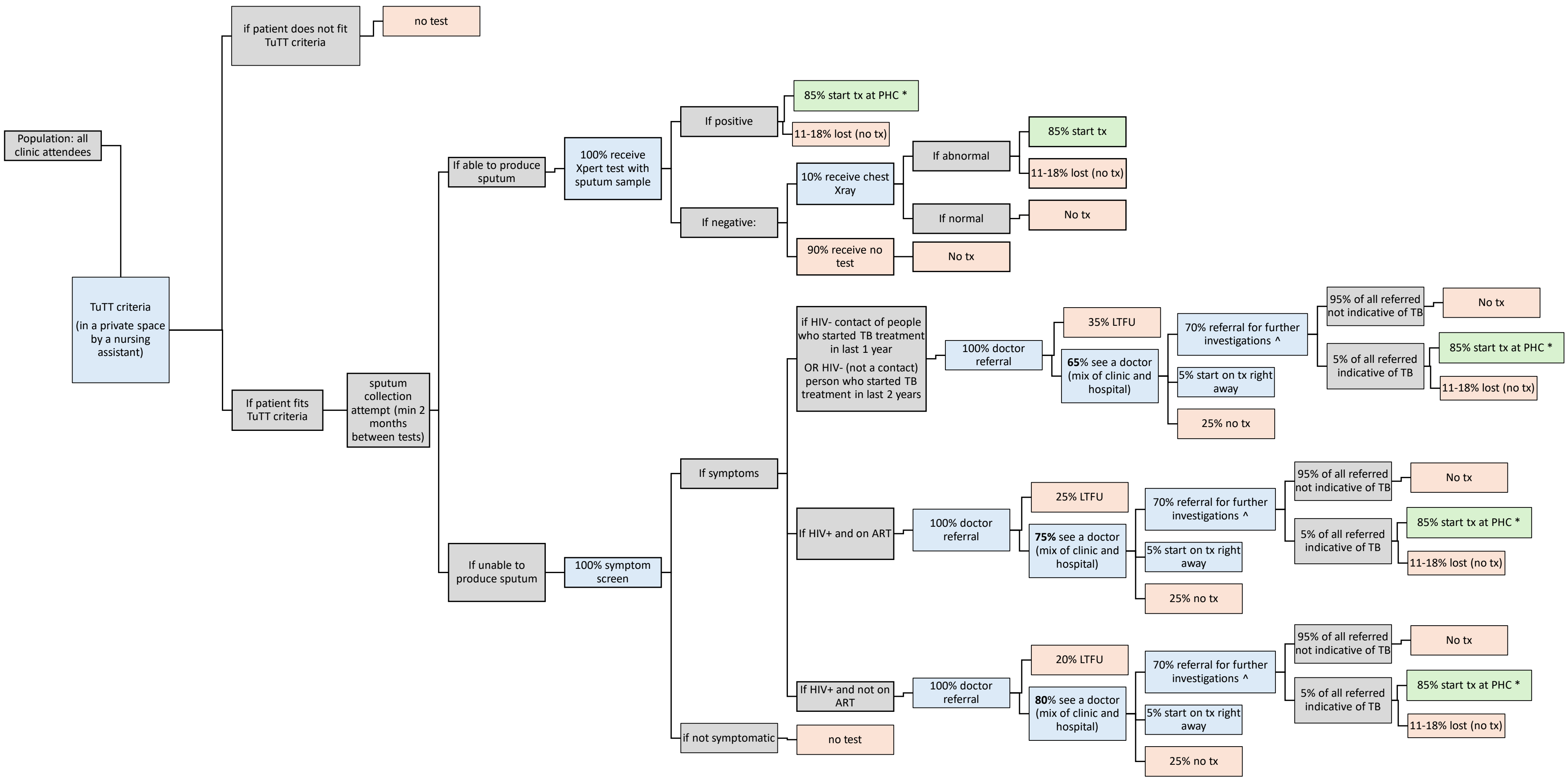


* Linkage to care activities: 50% of people testing positive called by nurse, 40% of people receiving GXP notified of result via automated text message by lab
^ Further investigations: 100% chest xray, 50% sputum attempt, 25% GeneXpert, 10% blood test FBC

### 5. Intensified TUTT allowing saliva samples

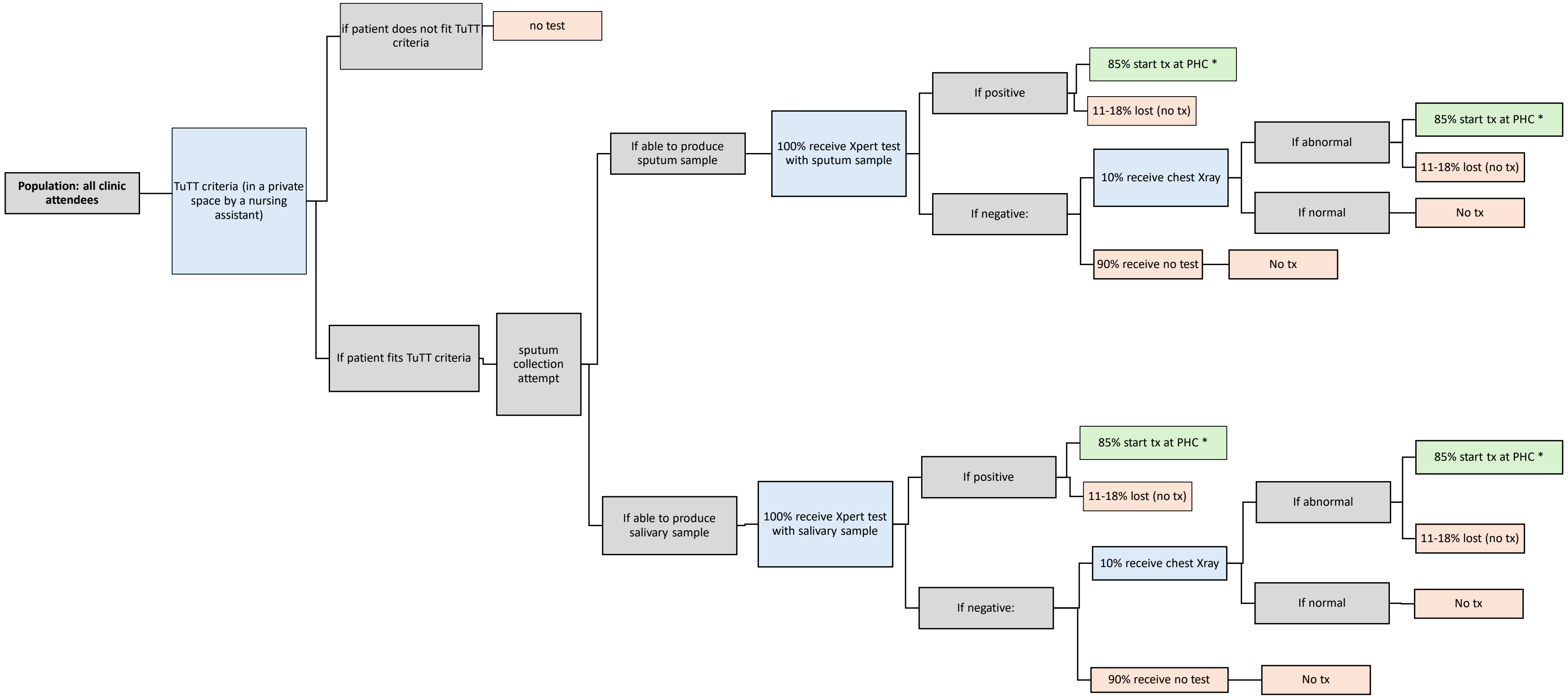


* Linkage to care activities: 50% of people testing positive called by nurse, 40% of people receiving GXP notified of result via automated text message by lab

^ Further investigations: 100% chest xray, 50% sputum attempt, 25% GeneXpert, 10% blood test FBC

### 6. Universal clinic symptom screening

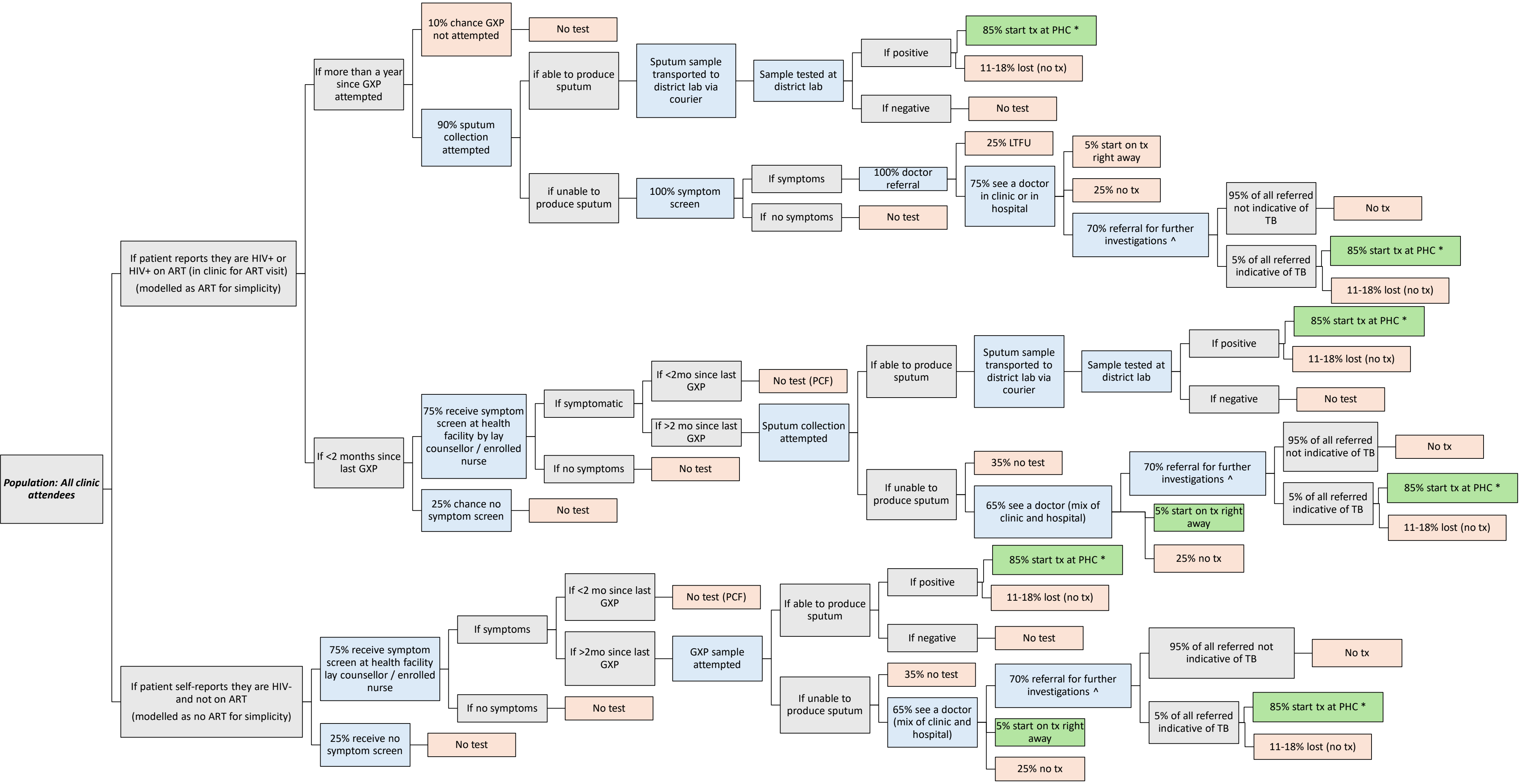


* Linkage to care activities: 50% of people testing positive called by nurse, 40% of people receiving GXP notified of result via automated text message by lab

^ Further investigations: 100% chest xray, 50% sputum attempt, 25% GeneXpert, 10% blood test FBC
